# Diet-wide association study of foods and nutrients with hip fracture risk: a prospective cohort study of 27,318 incident cases among 541,887 postmenopausal women

**DOI:** 10.64898/2026.02.02.26345368

**Authors:** Yang Tao, Timothy J. Key, Gillian K. Reeves, Sarah Floud, Keren Papier, Tammy Y.N. Tong

## Abstract

**Background:** Previous research on diet and hip fracture risk focused on selected foods and nutrients.

**Objective:** To conduct a diet-wide association study of hip fracture risk.

**Methods:** The study population comprised 541,887 postmenopausal women in the Million Women Study. Dietary information was assessed using a validated food frequency questionnaire in 2000-2004, and calibrated using a 24-hour dietary recall from a subset 10 years later. Cox regression was used to estimate hazard ratios (HR) and 95% confidence intervals (CI) for associations between 99 dietary factors and risk of incident hip fracture, ascertained through linkage to hospital admission data.

**Results:** After an average of 19.7 years of follow-up, 27,318 incident hip fractures occurred. In multivariable-adjusted models, 60 dietary factors were associated with hip fracture risk after correction for multiple testing (False Discovery Rate *p*-value <0.05). The five foods most significantly associated with hip fracture risk (based on *p*-value) were chicken (HR and 95% CI = 0.84, 0.80-0.87 per 20 g/day), vegetables (0.88, 0.85-0.91 per 100 g/day), pasta (0.87, 0.83-0.91 per 20 g/day), chips (1.14, 1.10-1.18 per 25 g/day), and fizzy drinks (1.18, 1.12-1.24 per 50 g/day). The five nutrients with the strongest associations were protein (0.79, 0.75-0.84 per 15 g/day), zinc (0.81, 0.77-0.86 per 2 mg/day), carotene (0.91, 0.88-0.94 per 1000 µg/day), fiber (0.89, 0.85-0.92 per 5 g/day), and niacin (0.82, 0.77-0.87 per 10 mg/day). The results were consistent across subgroups of body mass index, smoking, alcohol consumption, health status, physical activity, menopausal hormone use, deprivation status, and in analyses excluding the first five years of follow-up.

**Conclusions:** Many “healthy” foods were associated with lower risk of hip fracture, and “unhealthy” foods with higher risk. Further research is needed to assess whether these associations reflect causal relationships, are indicators of a healthier overall diet, or represent confounding from other lifestyle factors.

## Introduction

Hip fracture is a major public health issue in our ageing society. In 2025, there were nearly 72,000 incident hip fractures among people aged 60 and over in England, Wales, and Northern Ireland (1). Women and older people are high-risk populations, with a female to male ratio of around 3:1 in people aged 50+ years and more than 80% cases occurring in people aged 75 years and older (2). A recent population-based UK study reported that incident hip fractures increased in 2021-2024 compared with 2014-2019 for both men and women, resulting in an excess of 5595 more hip fractures (3). Hip fracture imposes a heavy economic burden by costing £2 billion per year due to hospitalization in the UK according to the 2024 National Hip Fracture Database report (4). Therefore, prevention of hip fractures is of great importance.

Diet is one of the modifiable risk factors for hip fracture. Evidence from clinical trials indicated that some dietary factors may be protective, including calcium and vitamin D supplementation from a recent meta-analysis of six randomized clinical trials (5) and increased dairy intake from a cluster randomized controlled trial (6). Meta-analytic evidence from prospective cohort studies suggested that higher protein intake (7) and adequate vegetables and fruits (8) were associated with a lower risk of hip fracture. However, the existing evidence is inconclusive mainly because it is based on limited numbers of studies, with insufficient cases and potential for confounding in the observational studies (9, 10). Moreover, prospective evidence to date has predominantly focused on calcium (5), vitamin D (5), and dairy (11), leaving many other dietary factors understudied (10). More prospective studies are needed, particularly those including a comprehensive range of dietary factors.

To address the limited and inconclusive evidence on many dietary factors, we conducted a diet-wide association study, systematically investigating the associations of 99 dietary factors with hip fracture risk in a large prospective cohort of postmenopausal women in the UK—the Million Women Study.

## Methods

### Study population

The Million Women Study is a large ongoing population-based prospective cohort study (12, 13). Women were recruited between 1996 and 2001 from 66 breast cancer screening centers across nine regions of England and one in Scotland. Women aged 50-64 were sent a recruitment questionnaire, with their invitation for routine National Health Service (NHS) breast screening. The recruitment questionnaire asked for information about anthropometric, socioeconomic, lifestyle, reproductive, and medical factors. In total 1.3 million women completed the recruitment questionnaire, which represented 25% of women in the targeted age group in the UK (12). All participants were followed up for hospital admissions, cancer registrations, and deaths through electronic linkage to routinely collected NHS data. Follow-up questionnaires were subsequently sent to all participants approximately every three to five years to collect up-to-date information on key factors of interest. The East of England-Cambridge South Research Ethics Committee approved the study, and all women gave written consent. More details are available on the Million Women Study website (14).

### Data collection and variables

#### Assessment of dietary intake

Dietary information was first collected through the follow-up questionnaire administered around 2001 (median year 2001, interquartile range 2000-2003; hereafter the “2001” questionnaire), which was the baseline for the main analysis. Dietary intake was obtained using a validated 130-item food frequency questionnaire (FFQ) (15) with good reproducibility (16). Participants were asked about their diet in a typical week using both quantitative and semi-quantitative questions, reporting how many times they consumed certain foods and food groups and ticking specific foods they ate once a week or more. Nutrient intakes were calculated using standard food portion sizes and the composition of each item (17, 18). Resurvey dietary data were collected, on average, a decade later in a subset of participants (7%) using a web-based 24-hour dietary recall questionnaire, the Oxford WebQ (19), which has been validated against biomarkers of key nutrients (20). In the WebQ assessment, participants were asked to report how many portions they consumed of each food item the previous day for 206 foods and 32 beverages. In total, 99 dietary factors (71 foods and 28 nutrients, **STable 1**) were available in both the baseline and the WebQ assessment and were therefore considered as exposures of interest in our diet-wide analyses.

#### Assessment of covariates

Covariates were based on information collected at the “2001” questionnaire, except for region of residence, deprivation status using the Townsend deprivation index (21), education level, physical activity level, height, and parity, which were based on information collected at recruitment around three years earlier. A Townsend deprivation index score for a small census area was assigned based on participants’ residential postcodes. Physical activity levels were derived from self-reported information on the frequency of strenuous exercise, defined as that which caused sweating or increased heartbeat.

#### Ascertainment of hip fracture

The outcome of interest for this study was incident hip fracture, and any first occurrence of hip fracture as a diagnosis in the hospital admission record was included. The outcomes were coded according to the relevant version of the International Classification of Diseases (ICD-10: S72.0-72.2 and ICD-9 820).

#### Exclusions

For the analyses, we excluded women 1) who had a previous hip fracture, 2) with insufficient dietary information (**SFigure 1**), 3) with unreliable dietary intake (baseline daily energy intake < 500 kcal or >3500 kcal, or self-reported changed diet within the previous five years due to illness) (**SFigure 1**). Moreover, due to the profound effect of menopause on bone mineral density (22), the observational period before age 55 years was excluded for women who were pre/perimenopausal (3%) or had missing information on menopausal status (16%) at baseline. We assumed women would be postmenopausal after age 55 because 96% of women in this cohort with a known age of natural menopause were postmenopausal by this age (23).

**Figure 1.**
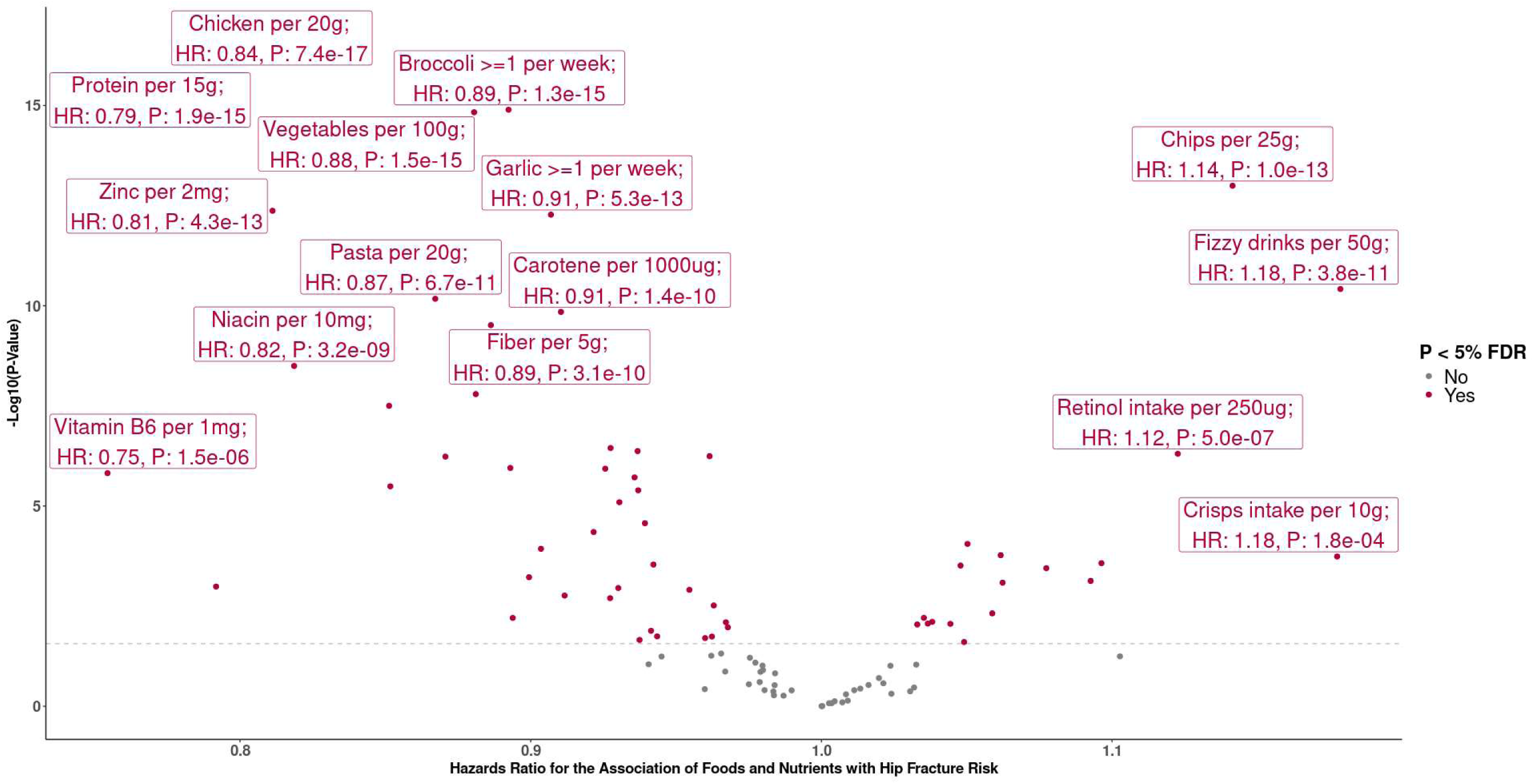
Volcano plot for the associations of foods and nutrients with hip fracture risk. The y-axis shows p-values (two-sided) for the associations between each of the 99 dietary factors with hip fracture risk calculated separately based on the fully adjusted Cox regression models, which were stratified by birth year (≤1921,five-year intervals between 1922 and 1946, >1946), year of completing the baseline dietary questionnaire (per year from 1999 to 2004, and 2005 or later), and 10 regions (9 in England and 1 in Scotland) and adjusted for ethnicity (white/ non-white, missing), Townsend deprivation index (quintiles and missing), education (tertiary, secondary, technical, no qualification, missing), self-rated health (excellent, good, fair, poor, missing), energy intake (500 to <1,000, 1,000 to <1,250, 1,250 to <1,500, 1,500 to <1,750, 1,750 to <2,000, 2,000 to < 2,250, 2,250 to ≤3,500 kcal/day), smoking (never smoker, past smoker, current smoker <15 cigarettes/day, current smoker ≥15 cigarettes/day, missing), alcohol consumption (<1, 1 to <10, 10 to <20, and ≥20g/day), strenuous exercise frequencies (none, <1, 1, 2-3, 4-6, 7 times/week, missing), MHT use (never, past, current, missing), parity (nulliparous, 1-2 children, ≥3 children, missing), age at menopause (<40, 40-44, 45-49, 50-54, ≥55 years, missing), height (quintiles and missing), family history of hip fractures (yes, no, missing), and BMI (<18.5, 18.5 to <20, 20 to <22.5, 22.5 to <25, 25 to <27.5, 27.5 to < 30, 30 to <32.5, ≥32.5 kg/m^2^, missing). For analyses of total energy and alcohol, covariates of energy intake and alcohol consumption were taken out of the model respectively. The x-axis shows the hazard ratios. Dietary factors significantly associated with hip fracture risk after False Discovery Rate (FDR) correction were in pink and those non-significant ones are in grey. The dotted line is the level of FDR-corrected significance threshold.

### Statistical analysis

#### Calibration for dietary data

We used the repeated measure of dietary intake, derived from the 24-hour dietary assessment Oxford WebQ, to calibrate dose response estimates based on the baseline intake categories. We first classified individuals according to self-reported intake of all 99 dietary factors at baseline. For the 64 quantitatively assessed dietary factors, baseline intake was grouped into three to five approximately equal-sized categories, using quintiles wherever possible. For each baseline intake category, we calculated the mean intake using the repeated measure in the subset of women who provided a WebQ assessment, and assigned the mean value to all women in that intake category. We then treated the mean intakes as a continuous variable to estimate hazard ratios (HR) associated with a predefined intake increment which was specific to each dietary factor. The increment was chosen to approximate the difference in mean intake based on the WebQ assessment between participants in the lowest and highest baseline intake categories for each dietary factor (**STable 1)**. Only WebQ data with reliable intake (daily energy ≤3,500 kcal/day; no lower cutoff applied given the single-day recall) and in women with no record of a prior hip fracture was used. If a woman completed more than one WebQ assessment, we only used the first one. The method of calibration aimed to reduce the measurement bias from regression dilution (24), and has been used for previous dietary research in this cohort (25, 26) and in UK Biobank (27). For the 35 foods with binary intake categories, we did not apply this calibration approach, but we checked the consistency of high and low intakes between baseline and the WebQ assessment and used baseline binary groups to calculate risks.

#### Main analyses

We used Cox proportional hazards regression models to estimate HR and 95% confidence intervals (CI) for the associations between each of the 99 dietary factors and hip fracture risk. Attained age was the underlying timeframe. Person-years were calculated from study entry (date of completion of the “2001” questionnaire or when women became postmenopausal) to whichever came first: first recorded admission for hip fracture, loss to follow-up, death, 90 years old, or the last date of follow-up available through electronic health records (31^st^ December 2023). Participants aged 90 and over were censored because more comorbidities may obscure the associations between diet and hip fracture risk.

All models were stratified by birth year (≤1921, five-year intervals between 1922 and 1946, >1946), year of completing the baseline dietary questionnaire (per year from 1999 to 2004, and 2005 or later), and ten recruitment regions (9 in England and 1 in Scotland). Model 1 was adjusted for ethnicity (white/non-white), Townsend deprivation index (quintiles), and educational qualifications (tertiary, secondary, technical, no qualification). Model 2 was further adjusted for self-rated health (excellent, good, fair, poor), energy intake (500 to <1,000, 1,000 to <1,250, 1,250 to <1,500, 1,500 to <1,750, 1,750 to <2,000, 2,000 to < 2,250, 2,250 to 3,500 kcal/day), smoking (never smoker, past smoker, current smoker <15 cigarettes/day, and current smoker ≥15 cigarettes/day), alcohol consumption (<1, 1 to <10, 10 to <20, and ≥20 g/day), strenuous exercise frequency (none, <1, 1, 2-3, 4-6, 7 times/week), menopausal hormone therapy (MHT) use (never, past, and current), parity (nulliparous, 1-2 children, ≥3 children), age at menopause (<40, 40-44, 45-49, 50-54, ≥55 years), height (quintiles), and family history of hip fracture (yes, no). Model 3 was further adjusted for BMI (<18.5, 18.5 to <20, 20 to <22.5, 22.5 to <25, 25 to <27.5, 27.5 to < 30, 30 to <32.5, ≥32.5 kg/m^2^). The proportion of women with missing data was less than 5% for all covariates except for age at menopause which had 13% missing data, and such women were assigned to a “missing” category in the analyses. We used the Benjamini-Hochberg false discovery rate (FDR) method with alpha=0.05 to adjust for multiple testing (28). To assess the extent to which associations of dietary factors with hip fracture risk were likely to be independent, Spearman correlations were used to assess the correlations among baseline dietary factors with quantitative information that were FDR-significant.

#### Sensitivity analyses

Stratified analyses, aimed at examining the potential for residual confounding and evidence of effect modification, were conducted for BMI (<20, 20-24.9, ≥25 kg/m^2^), smoking status (never smoker, ever smoker), frequency of strenuous exercise (rarely or <once/week, 1-6 times/week, daily), MHT use (never, ever), and deprivation status (least deprived, most deprived). For alcohol consumption, since the non-drinkers could be a composite of never-drinkers and people who abstain due to illness, we conducted stratified analyses in mild/moderate drinkers (1 to <10g/day) and in regular drinkers (≥10g/day). For baseline health status, stratified analyses were conducted in two groups: (1) individuals with excellent or good health and no bone-related comorbidities, and (2) individuals with fair or poor health or those with bone-related comorbidities, including self-reported baseline cardiovascular diseases, diabetes, cancer, gallstones, inflammatory bowel diseases, thyroid disease, osteoarthritis, and rheumatoid arthritis which were likely to affect bone health or nutrient absorption (29). We also repeated the analyses in the “healthiest” subgroup based on a combination of characteristics, which was defined as women who had normal BMI (20-24.9 kg/m^2^), had never smoked, were mild/moderate drinkers (alcohol consumption 1 to <10g/day), and had never used MHT, and thus were the least likely to be influenced by these potential confounders. To explore the possible impact of reverse causation, we excluded the first five years of follow-up in additional analyses.

All analyses were conducted with STATA version 18.0 (StataCorporation, Inc., College Station, TX, USA), and figures were created with “gglot2” package in R version 4.3.2.

## Results

After excluding participants with previous hip fracture and unreliable dietary information at baseline, 541,887 women were included in the analyses (**SFigure 1**). During an average of 19.7 (SD 4.6) years of follow-up, 27,318 women had a hip fracture. The baseline characteristics are shown in **Table 1** and **STable 2**. Compared with women without hip fracture, those with incident hip fracture were older and more deprived, and self-reported being less educated, more likely to smoke, drink less alcohol, engage in less strenuous exercise, be taller with lower BMI, have fewer offspring, experience earlier menopause, be less likely to report MHT use, and more frequently report a family history of hip fracture and fair or poor baseline health.

**Table 1.**
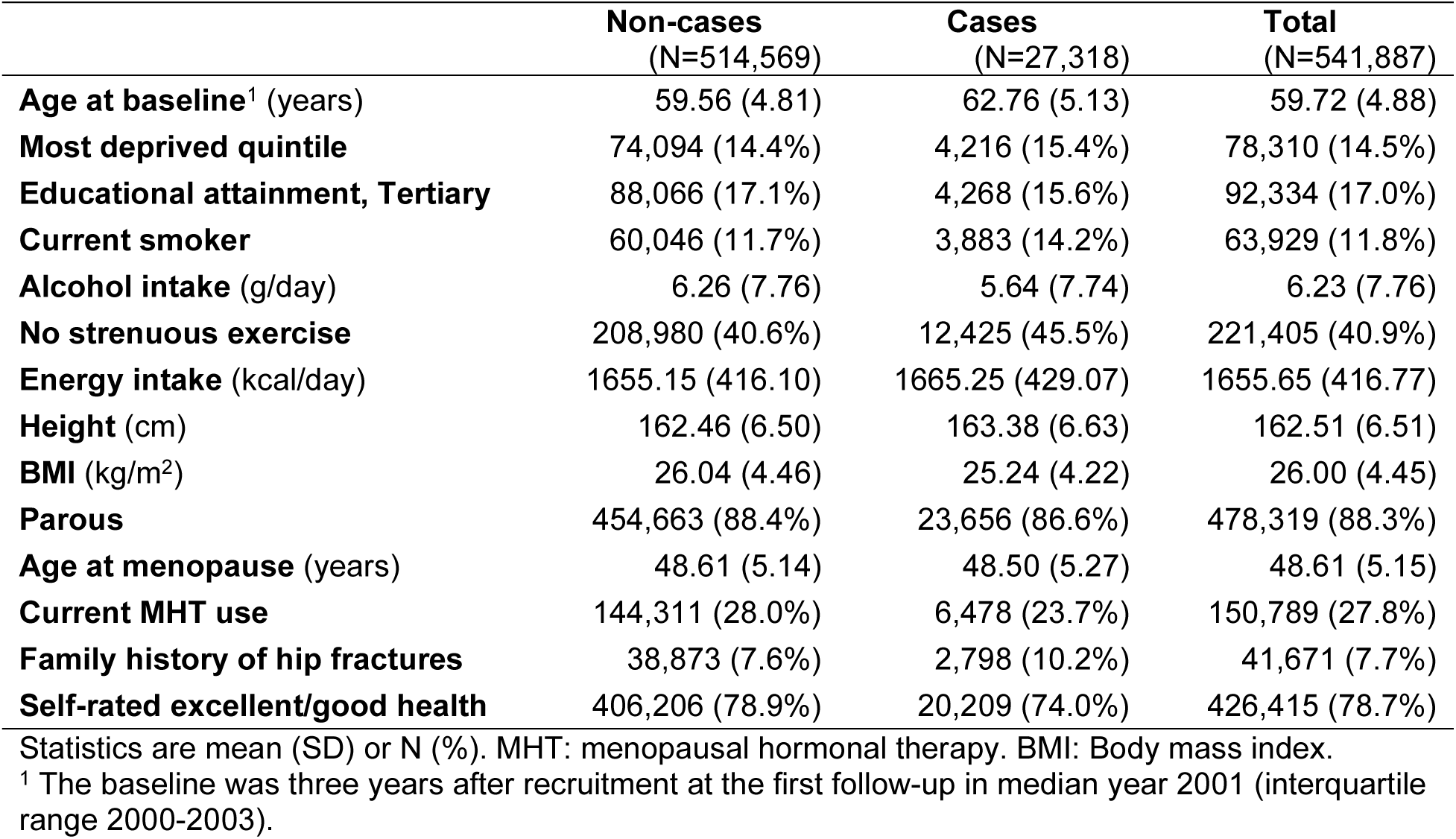
Baseline characteristics of participants with and without hip fractures.

|  | **Non-cases**<br>(N=514,569) | **Cases**<br>(N=27,318) | **Total**<br>(N=541,887) |
| --- | --- | --- | --- |
| **Age at baseline<sup>1</sup>** (years) | 59.56 (4.81) | 62.76 (5.13) | 59.72 (4.88) |
| **Most deprived quintile** | 74,094 (14.4%) | 4,216 (15.4%) | 78,310 (14.5%) |
| **Educational attainment, Tertiary** | 88,066 (17.1%) | 4,268 (15.6%) | 92,334 (17.0%) |
| **Current smoker** | 60,046 (11.7%) | 3,883 (14.2%) | 63,929 (11.8%) |
| **Alcohol intake** (g/day) | 6.26 (7.76) | 5.64 (7.74) | 6.23 (7.76) |
| **No strenuous exercise** | 208,980 (40.6%) | 12,425 (45.5%) | 221,405 (40.9%) |
| **Energy intake** (kcal/day) | 1655.15 (416.10) | 1665.25 (429.07) | 1655.65 (416.77) |
| **Height** (cm) | 162.46 (6.50) | 163.38 (6.63) | 162.51 (6.51) |
| **BMI** (kg/m<sup>2</sup>) | 26.04 (4.46) | 25.24 (4.22) | 26.00 (4.45) |
| **Parous** | 454,663 (88.4%) | 23,656 (86.6%) | 478,319 (88.3%) |
| **Age at menopause** (years) | 48.61 (5.14) | 48.50 (5.27) | 48.61 (5.15) |
| **Current MHT use** | 144,311 (28.0%) | 6,478 (23.7%) | 150,789 (27.8%) |
| **Family history of hip fractures** | 38,873 (7.6%) | 2,798 (10.2%) | 41,671 (7.7%) |
| **Self-rated excellent/good health** | 406,206 (78.9%) | 20,209 (74.0%) | 426,415 (78.7%) |

Statistics are mean (SD) or N (%). MHT: menopausal hormonal therapy. BMI: Body mass index.
<sup>1</sup> The baseline was three years after recruitment at the first follow-up in median year 2001 (interquartile range 2000-2003).

For foods with multiple intake categories, women consumed more fresh fruits and vegetables, cheese, meat, chips, and cereal, but reported lower intake of dried fruit, grains, sweets, sauces, and all drinks (tea, coffee, fruit juice, fizzy drinks, and squash drinks) at resurvey compared with baseline. Overall, the differences between the highest and the lowest fifths of dietary factors were substantially narrower at resurvey (**STable 1**). For the 35 foods with binary categories, the classification of high and low categories was consistent between baseline and resurvey data. Average nutrient intakes at baseline and resurvey were in general similar, although women tended to report slightly higher intakes of all nutrients at the resurvey, except for free sugars.

Based on the fully adjusted model, the associations of 60 out of 99 dietary factors were significant after FDR correction for multiple testing (**Figure 1**, **Table 2**, and **STable 3**). Of the 60 dietary factors, 25 foods and 17 nutrients were inversely associated with hip fracture risk, including chicken (HR and 95% CI: 0.84, 0.80-0.87 per 20 g/day), vegetables (0.88, 0.85-0.91 per 100 g/day), and pasta (0.87, 0.83-0.91 per 20 g/day), which were ranked as the top foods based on the lowest *p*-values; and protein (0.79, 0.75-0.84 per 15 g/day), zinc (0.81, 0.77-0.86 per 2 mg/day), carotene (0.91, 0.88-0.94 per 1000 µg/day), fiber (0.89, 0.85-0.92 per 5 g/day), and niacin (0.82, 0.77-0.87 per 10 mg/day) as the top nutrients. Other FDR-significant foods associated with lower risk were specific kinds of vegetables (broccoli, garlic, onion, pepper, courgette, carrot, lettuce, cucumber, mushrooms) and fruits (total fruits excluding juice, apple, banana, avocado, stone fruits, dried fruits), wholegrains, rice, brown bread, cereal, yoghurt, nuts, and oily fish. Other FDR-significant nutrients associated with lower risk were phosphorus, alcohol, selenium, vitamin B_6_, magnesium, potassium, thiamin, vitamin C, vitamin B_12_, riboflavin, calcium, and folate. In contrast, 15 foods were positively associated with hip fracture risk, with chips and fizzy drinks ranked as the top foods (HR and 95% CI 1.14, 1.10-1.18 per 25g/day and 1.18, 1.12-1.24 per 50g/day) based on *p*-values. Other FDR-significant foods associated with higher risk included some vegetables (Brussels sprouts, cauliflower, green peas), stewed fruits, baked beans, squash drinks, juice, white bread, sweet biscuits, eggs, processed meat, crisps, and soup. There were only three FDR-significant nutrients associated with higher risk: retinol (1.12, 1.07-1.17 per 250 µg/day), free sugars (1.10, 1.04-1.15 per 30g/day), and saturated fatty acids (SFA) (1.09, 1.04-1.15 per 30g/day).

**Table 2.**
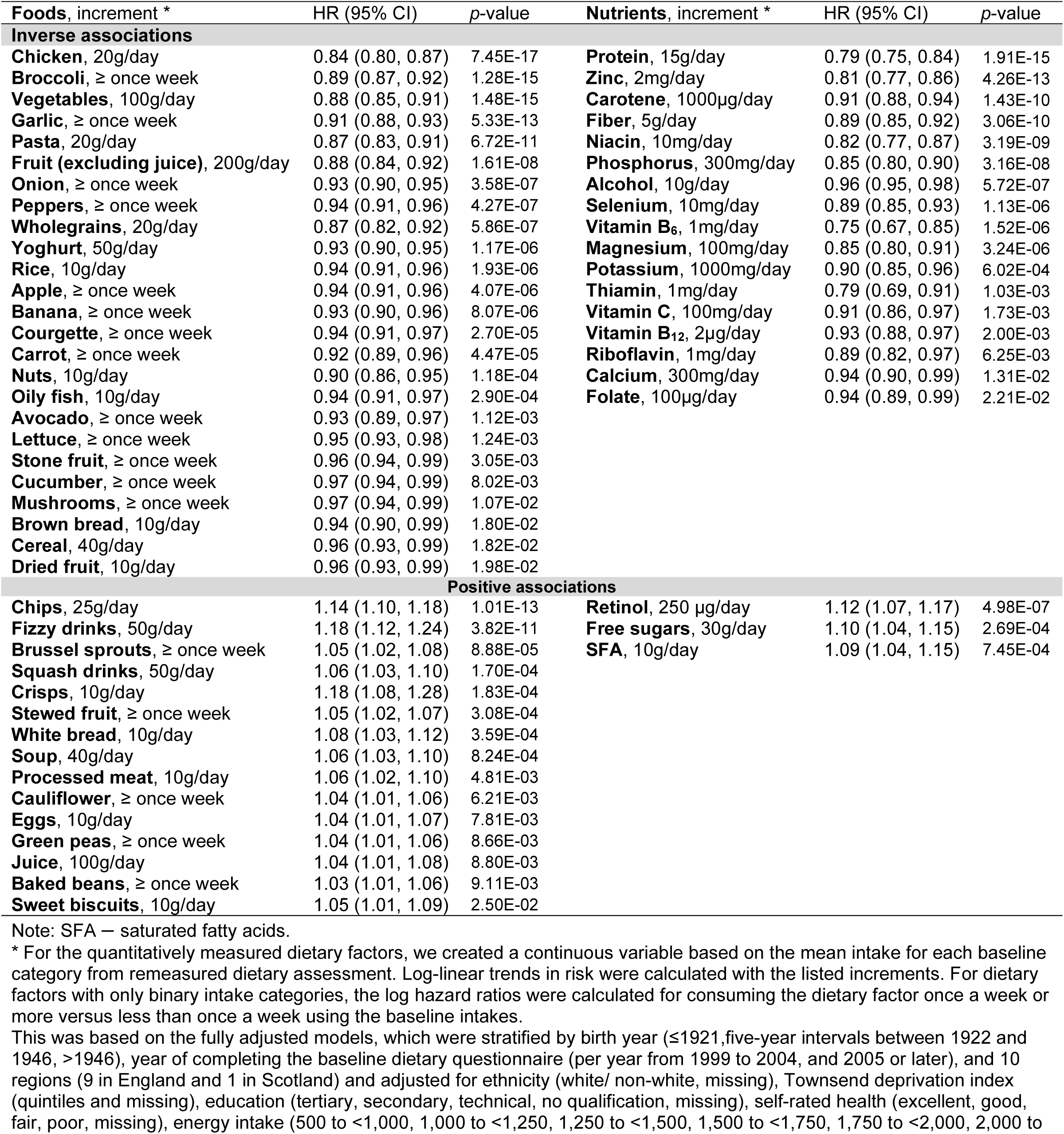
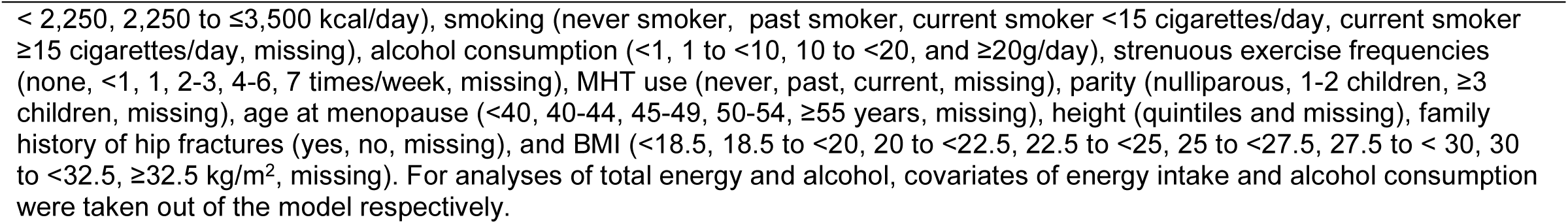
Risk of hip fracture for 60 FDR-significant dietary factors by direction of effects, ranked based on *p*-value (Person-years = 10,652,303, 27,318 incident cases)

| **Foods, increment \*** | **HR (95% CI)** | ***p*-value** | **Nutrients, increment \*** | **HR (95% CI)** | ***p*-value** |
| --- | --- | --- | --- | --- | --- |
| **Inverse associations** |  |  |  |  |  |
| Chicken, 20g/day | 0.84 (0.80, 0.87) | 7.45E-17 | Protein, 15g/day | 0.79 (0.75, 0.84) | 1.91E-15 |
| Broccoli, ≥ once week | 0.89 (0.87, 0.92) | 1.28E-15 | Zinc, 2mg/day | 0.81 (0.77, 0.86) | 4.26E-13 |
| Vegetables, 100g/day | 0.88 (0.85, 0.91) | 1.48E-15 | Carotene, 1000µg/day | 0.91 (0.88, 0.94) | 1.43E-10 |
| Garlic, ≥ once week | 0.91 (0.88, 0.93) | 5.33E-13 | Fiber, 5g/day | 0.89 (0.85, 0.92) | 3.06E-10 |
| Pasta, 20g/day | 0.87 (0.83, 0.91) | 6.72E-11 | Niacin, 10mg/day | 0.82 (0.77, 0.87) | 3.19E-09 |
| Fruit (excluding juice), 200g/day | 0.88 (0.84, 0.92) | 1.61E-08 | Phosphorus, 300mg/day | 0.85 (0.80, 0.90) | 3.16E-08 |
| Onion, ≥ once week | 0.93 (0.90, 0.95) | 3.58E-07 | Alcohol, 10g/day | 0.96 (0.95, 0.98) | 5.72E-07 |
| Peppers, ≥ once week | 0.94 (0.91, 0.96) | 4.27E-07 | Selenium, 10mg/day | 0.89 (0.85, 0.93) | 1.13E-06 |
| Wholegrains, 20g/day | 0.87 (0.82, 0.92) | 5.86E-07 | Vitamin B<sub>6</sub>, 1mg/day | 0.75 (0.67, 0.85) | 1.52E-06 |
| Yoghurt, 50g/day | 0.93 (0.90, 0.95) | 1.17E-06 | Magnesium, 100mg/day | 0.85 (0.80, 0.91) | 3.24E-06 |
| Rice, 10g/day | 0.94 (0.91, 0.96) | 1.93E-06 | Potassium, 1000mg/day | 0.90 (0.85, 0.96) | 6.02E-04 |
| Apple, ≥ once week | 0.94 (0.91, 0.96) | 4.07E-06 | Thiamin, 1mg/day | 0.79 (0.69, 0.91) | 1.03E-03 |
| Banana, ≥ once week | 0.93 (0.90, 0.96) | 8.07E-06 | Vitamin C, 100mg/day | 0.91 (0.86, 0.97) | 1.73E-03 |
| Courgette, ≥ once week | 0.94 (0.91, 0.97) | 2.70E-05 | Vitamin B<sub>12</sub>, 2µg/day | 0.93 (0.88, 0.97) | 2.00E-03 |
| Carrot, ≥ once week | 0.92 (0.89, 0.96) | 4.47E-05 | Riboflavin, 1mg/day | 0.89 (0.82, 0.97) | 6.25E-03 |
| Nuts, 10g/day | 0.90 (0.86, 0.95) | 1.18E-04 | Calcium, 300mg/day | 0.94 (0.90, 0.99) | 1.31E-02 |
| Oily fish, 10g/day | 0.94 (0.91, 0.97) | 2.90E-04 | Folate, 100µg/day | 0.94 (0.89, 0.99) | 2.21E-02 |
| Avocado, ≥ once week | 0.93 (0.89, 0.97) | 1.12E-03 |  |  |  |
| Lettuce, ≥ once week | 0.95 (0.93, 0.98) | 1.24E-03 |  |  |  |
| Stone fruit, ≥ once week | 0.96 (0.94, 0.99) | 3.05E-03 |  |  |  |
| Cucumber, ≥ once week | 0.97 (0.94, 0.99) | 8.02E-03 |  |  |  |
| Mushrooms, ≥ once week | 0.97 (0.94, 0.99) | 1.07E-02 |  |  |  |
| Brown bread, 10g/day | 0.94 (0.90, 0.99) | 1.80E-02 |  |  |  |
| Cereal, 40g/day | 0.96 (0.93, 0.99) | 1.82E-02 |  |  |  |
| Dried fruit, 10g/day | 0.96 (0.93, 0.99) | 1.98E-02 |  |  |  |
| **Positive associations** |  |  |  |  |  |
| Chips, 25g/day | 1.14 (1.10, 1.18) | 1.01E-13 | Retinol, 250 µg/day | 1.12 (1.07, 1.17) | 4.98E-07 |
| Fizzy drinks, 50g/day | 1.18 (1.12, 1.24) | 3.82E-11 | Free sugars, 30g/day | 1.10 (1.04, 1.15) | 2.69E-04 |
| Brussel sprouts, ≥ once week | 1.05 (1.02, 1.08) | 8.88E-05 | SFA, 10g/day | 1.09 (1.04, 1.15) | 7.45E-04 |
| Squash drinks, 50g/day | 1.06 (1.03, 1.10) | 1.70E-04 |  |  |  |
| Crisps, 10g/day | 1.18 (1.08, 1.28) | 1.83E-04 |  |  |  |
| Stewed fruit, ≥ once week | 1.05 (1.02, 1.07) | 3.08E-04 |  |  |  |
| White bread, 10g/day | 1.08 (1.03, 1.12) | 3.59E-04 |  |  |  |
| Soup, 40g/day | 1.06 (1.03, 1.10) | 8.24E-04 |  |  |  |
| Processed meat, 10g/day | 1.06 (1.02, 1.10) | 4.81E-03 |  |  |  |
| Cauliflower, ≥ once week | 1.04 (1.01, 1.06) | 6.21E-03 |  |  |  |
| Eggs, 10g/day | 1.04 (1.01, 1.07) | 7.81E-03 |  |  |  |
| Green peas, ≥ once week | 1.04 (1.01, 1.06) | 8.66E-03 |  |  |  |
| Juice, 100g/day | 1.04 (1.01, 1.08) | 8.80E-03 |  |  |  |
| Baked beans, ≥ once week | 1.03 (1.01, 1.06) | 9.11E-03 |  |  |  |
| Sweet biscuits, 10g/day | 1.05 (1.01, 1.09) | 2.50E-02 |  |  |  |

Note: SFA — saturated fatty acids.
\* For the quantitatively measured dietary factors, we created a continuous variable based on the mean intake for each baseline category from remeasured dietary assessment. Log-linear trends in risk were calculated with the listed increments. For dietary factors with only binary intake categories, the log hazard ratios were calculated for consuming the dietary factor once a week or more versus less than once a week using the baseline intakes.

Among the 60 significant dietary factors, we selected all the foods and nutrients with three or more categories to conduct pair-wise Spearman correlations to evaluate the likelihood of independence of these associations (**SFigure 2**). Taking some of the top five foods and top five nutrients as examples, protein was strongly correlated (defined as r≥0.8) with zinc and phosphorus and moderately-strongly correlated (defined as 0.6≤r<0.8) with vitamin B_6_, thiamin, magnesium, potassium, niacin, selenium, vitamin B_12_, calcium, riboflavin, and folate. Vegetables were strongly correlated with carotene, and moderately-strongly correlated with fiber, vitamin C, folate, potassium, and vitamin B_6_. Chicken was moderately correlated (defined as 0.4≤r<0.6) with protein and niacin. Weak correlations (defined as 0.2≤r<0.4) were found between fizzy drinks and free sugars, chips and white bread, and pasta and rice.

The results of stratified analyses by key covariates for the top five foods and the top five nutrients (based on *p*-values) are shown in **Table 3** and **Table 4**, including BMI, smoking, alcohol consumption, baseline health status, frequency of strenuous exercise, MHT use, and deprivation status. For the 60 dietary factors significantly associated with hip fracture risk, the majority of the results were consistent across subgroups (**Table 3**, **Table 4**, **STable 4** – **STable 10**). However, the inverse association observed for alcohol overall became a positive association in regular drinkers (alcohol consumption ≥10g/day, HR and 95% CI 1.10, 1.05-1.15 per 10g/day, **STable 6**). Also, modest heterogeneity was seen for baked beans by MHT use (**STable 9**).

**Table 3.**
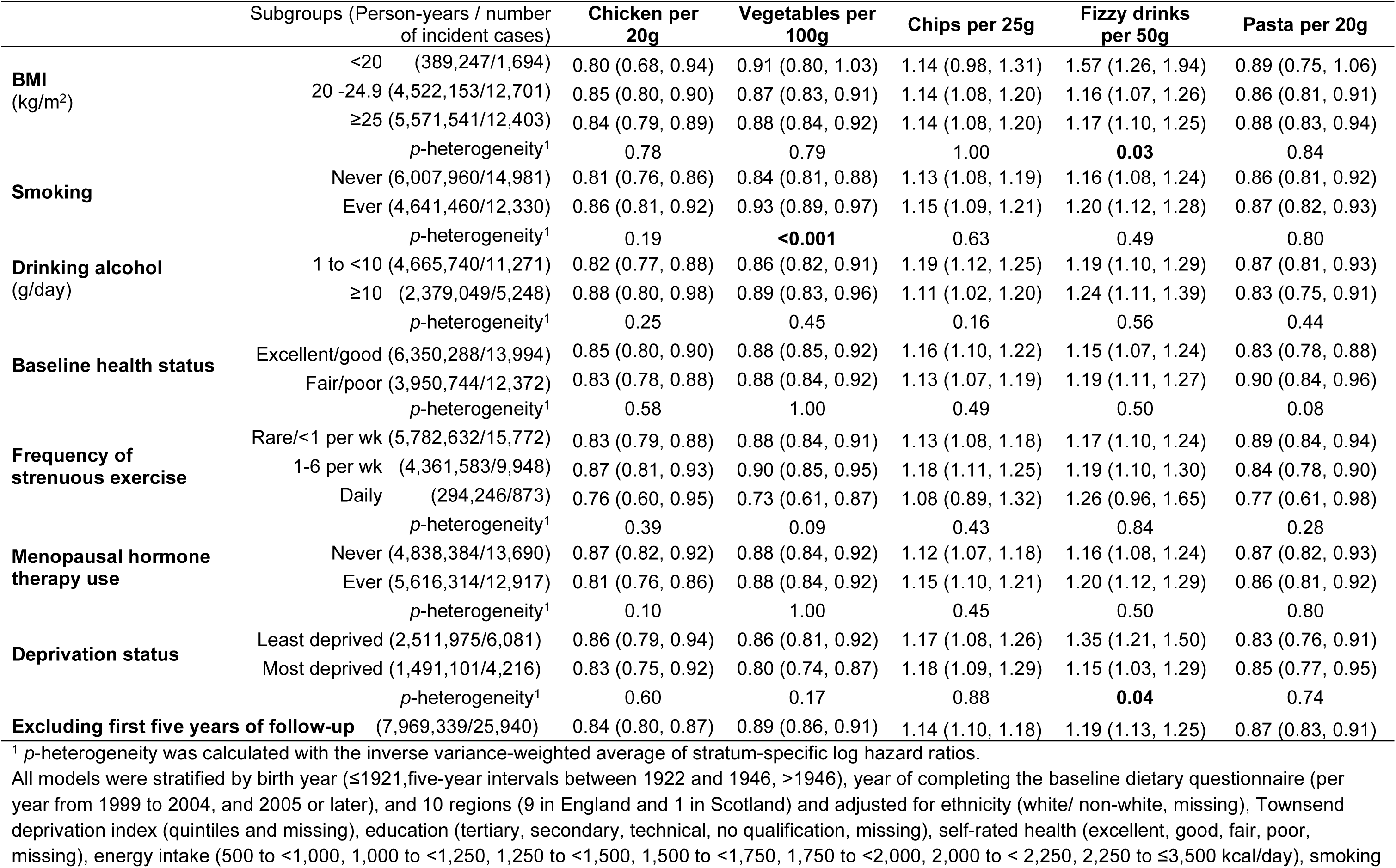
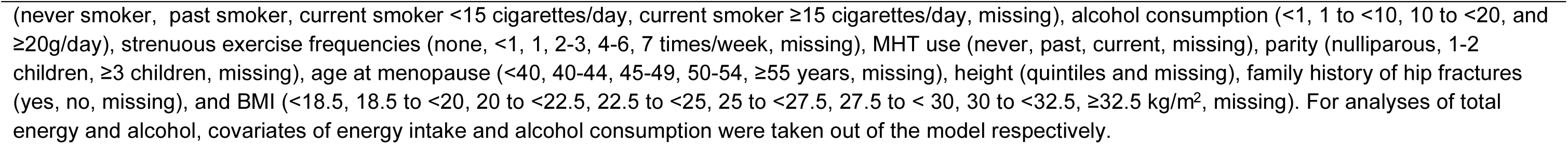
Stratified and sensitivity analyses for associations between hip fracture risk and top 5 foods.

|  | Subgroups (Person-years / number of incident cases) | Chicken per 20g | Vegetables per 100g | Chips per 25g | Fizzy drinks per 50g | Pasta per 20g |
| --- | --- | --- | --- | --- | --- | --- |
| **BMI**<br>(kg/m<sup>2</sup>) | <20 (389,247/1,694) | 0.80 (0.68, 0.94) | 0.91 (0.80, 1.03) | 1.14 (0.98, 1.31) | 1.57 (1.26, 1.94) | 0.89 (0.75, 1.06) |
|  | 20 -24.9 (4,522,153/12,701) | 0.85 (0.80, 0.90) | 0.87 (0.83, 0.91) | 1.14 (1.08, 1.20) | 1.16 (1.07, 1.26) | 0.86 (0.81, 0.91) |
|  | ≥25 (5,571,541/12,403) | 0.84 (0.79, 0.89) | 0.88 (0.84, 0.92) | 1.14 (1.08, 1.20) | 1.17 (1.10, 1.25) | 0.88 (0.83, 0.94) |
|  | *p*-heterogeneity<sup>1</sup> | 0.78 | 0.79 | 1.00 | **0.03** | 0.84 |
| **Smoking** | Never (6,007,960/14,981) | 0.81 (0.76, 0.86) | 0.84 (0.81, 0.88) | 1.13 (1.08, 1.19) | 1.16 (1.08, 1.24) | 0.86 (0.81, 0.92) |
|  | Ever (4,641,460/12,330) | 0.86 (0.81, 0.92) | 0.93 (0.89, 0.97) | 1.15 (1.09, 1.21) | 1.20 (1.12, 1.28) | 0.87 (0.82, 0.93) |
|  | *p*-heterogeneity<sup>1</sup> | 0.19 | **<0.001** | 0.63 | 0.49 | 0.80 |
| **Drinking alcohol**<br>(g/day) | 1 to <10 (4,665,740/11,271) | 0.82 (0.77, 0.88) | 0.86 (0.82, 0.91) | 1.19 (1.12, 1.25) | 1.19 (1.10, 1.29) | 0.87 (0.81, 0.93) |
|  | ≥10 (2,379,049/5,248) | 0.88 (0.80, 0.98) | 0.89 (0.83, 0.96) | 1.11 (1.02, 1.20) | 1.24 (1.11, 1.39) | 0.83 (0.75, 0.91) |
|  | *p*-heterogeneity<sup>1</sup> | 0.25 | 0.45 | 0.16 | 0.56 | 0.44 |
| **Baseline health status** | Excellent/good (6,350,288/13,994) | 0.85 (0.80, 0.90) | 0.88 (0.85, 0.92) | 1.16 (1.10, 1.22) | 1.15 (1.07, 1.24) | 0.83 (0.78, 0.88) |
|  | Fair/poor (3,950,744/12,372) | 0.83 (0.78, 0.88) | 0.88 (0.84, 0.92) | 1.13 (1.07, 1.19) | 1.19 (1.11, 1.27) | 0.90 (0.84, 0.96) |
|  | *p*-heterogeneity<sup>1</sup> | 0.58 | 1.00 | 0.49 | 0.50 | 0.08 |
| **Frequency of strenuous exercise** | Rare/<1 per wk (5,782,632/15,772) | 0.83 (0.79, 0.88) | 0.88 (0.84, 0.91) | 1.13 (1.08, 1.18) | 1.17 (1.10, 1.24) | 0.89 (0.84, 0.94) |
|  | 1-6 per wk (4,361,583/9,948) | 0.87 (0.81, 0.93) | 0.90 (0.85, 0.95) | 1.18 (1.11, 1.25) | 1.19 (1.10, 1.30) | 0.84 (0.78, 0.90) |
|  | Daily (294,246/873) | 0.76 (0.60, 0.95) | 0.73 (0.61, 0.87) | 1.08 (0.89, 1.32) | 1.26 (0.96, 1.65) | 0.77 (0.61, 0.98) |
|  | *p*-heterogeneity<sup>1</sup> | 0.39 | 0.09 | 0.43 | 0.84 | 0.28 |
| **Menopausal hormone therapy use** | Never (4,838,384/13,690) | 0.87 (0.82, 0.92) | 0.88 (0.84, 0.92) | 1.12 (1.07, 1.18) | 1.16 (1.08, 1.24) | 0.87 (0.82, 0.93) |
|  | Ever (5,616,314/12,917) | 0.81 (0.76, 0.86) | 0.88 (0.84, 0.92) | 1.15 (1.10, 1.21) | 1.20 (1.12, 1.29) | 0.86 (0.81, 0.92) |
|  | *p*-heterogeneity<sup>1</sup> | 0.10 | 1.00 | 0.45 | 0.50 | 0.80 |
| **Deprivation status** | Least deprived (2,511,975/6,081) | 0.86 (0.79, 0.94) | 0.86 (0.81, 0.92) | 1.17 (1.08, 1.26) | 1.35 (1.21, 1.50) | 0.83 (0.76, 0.91) |
|  | Most deprived (1,491,101/4,216) | 0.83 (0.75, 0.92) | 0.80 (0.74, 0.87) | 1.18 (1.09, 1.29) | 1.15 (1.03, 1.29) | 0.85 (0.77, 0.95) |
|  | *p*-heterogeneity<sup>1</sup> | 0.60 | 0.17 | 0.88 | **0.04** | 0.74 |
| **Excluding first five years of follow-up** (7,969,339/25,940) |  | 0.84 (0.80, 0.87) | 0.89 (0.86, 0.91) | 1.14 (1.10, 1.18) | 1.19 (1.13, 1.25) | 0.87 (0.83, 0.91) |

<sup>1</sup> *p*-heterogeneity was calculated with the inverse variance-weighted average of stratum-specific log hazard ratios.

**Table 4.**
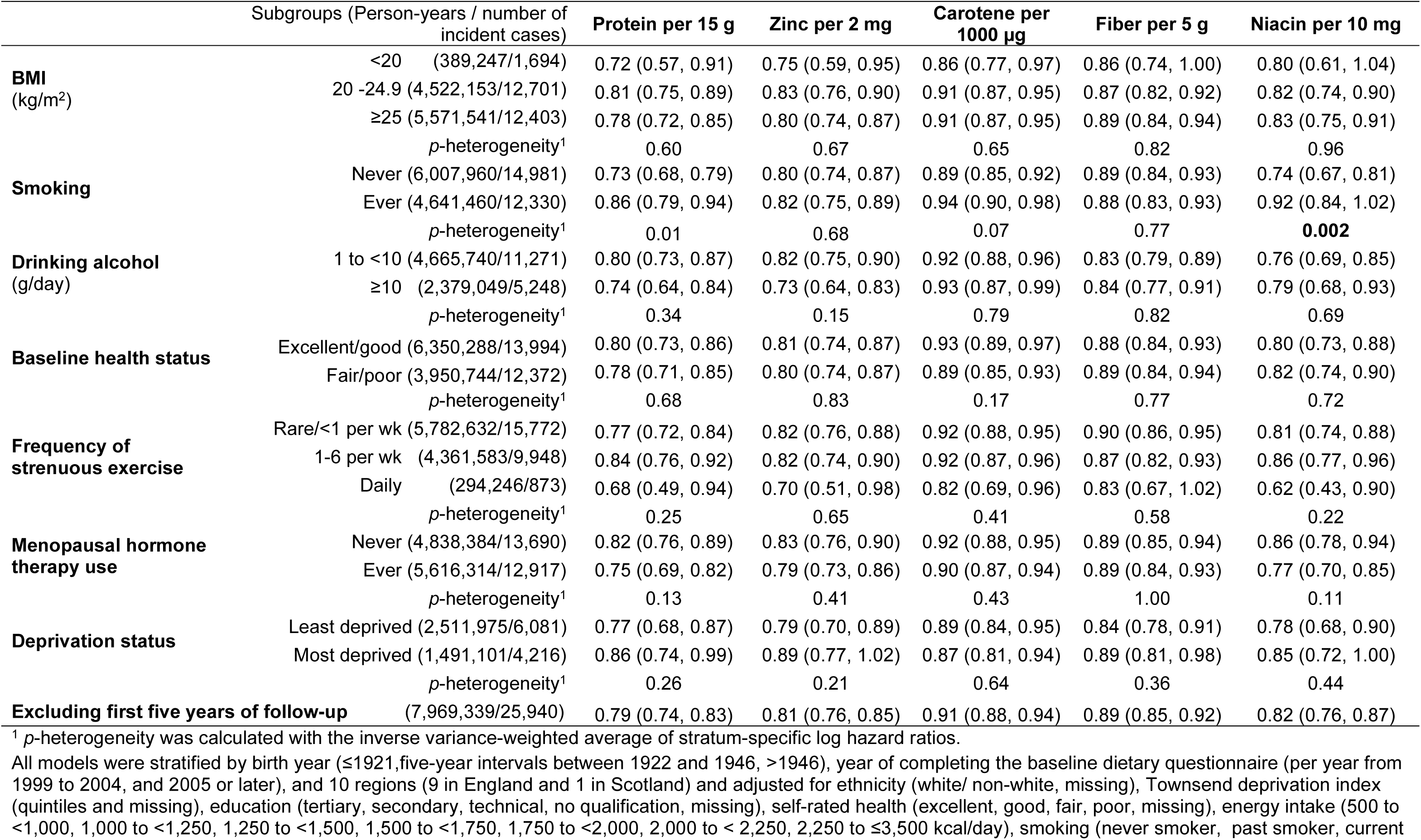
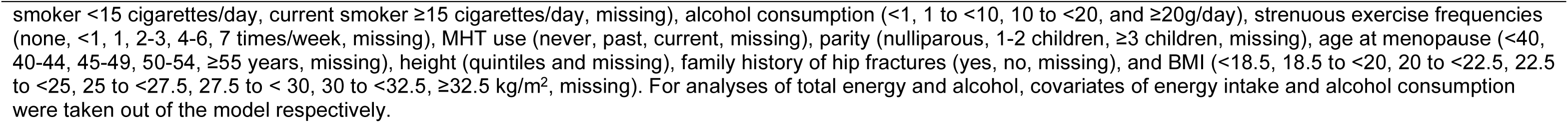
Stratified and sensitivity analyses for associations between hip fracture risk and top 5 nutrients.

|  | Subgroups (Person-years / number of incident cases) | Protein per 15 g | Zinc per 2 mg | Carotene per 1000 µg | Fiber per 5 g | Niacin per 10 mg |
| --- | --- | --- | --- | --- | --- | --- |
| **BMI**<br>(kg/m²) | <20 (389,247/1,694) | 0.72 (0.57, 0.91) | 0.75 (0.59, 0.95) | 0.86 (0.77, 0.97) | 0.86 (0.74, 1.00) | 0.80 (0.61, 1.04) |
|  | 20 -24.9 (4,522,153/12,701) | 0.81 (0.75, 0.89) | 0.83 (0.76, 0.90) | 0.91 (0.87, 0.95) | 0.87 (0.82, 0.92) | 0.82 (0.74, 0.90) |
|  | ≥25 (5,571,541/12,403) | 0.78 (0.72, 0.85) | 0.80 (0.74, 0.87) | 0.91 (0.87, 0.95) | 0.89 (0.84, 0.94) | 0.83 (0.75, 0.91) |
|  | *p*-heterogeneity¹ | 0.60 | 0.67 | 0.65 | 0.82 | 0.96 |
| **Smoking** | Never (6,007,960/14,981) | 0.73 (0.68, 0.79) | 0.80 (0.74, 0.87) | 0.89 (0.85, 0.92) | 0.89 (0.84, 0.93) | 0.74 (0.67, 0.81) |
|  | Ever (4,641,460/12,330) | 0.86 (0.79, 0.94) | 0.82 (0.75, 0.89) | 0.94 (0.90, 0.98) | 0.88 (0.83, 0.93) | 0.92 (0.84, 1.02) |
|  | *p*-heterogeneity¹ | 0.01 | 0.68 | 0.07 | 0.77 | **0.002** |
| **Drinking alcohol**<br>(g/day) | 1 to <10 (4,665,740/11,271) | 0.80 (0.73, 0.87) | 0.82 (0.75, 0.90) | 0.92 (0.88, 0.96) | 0.83 (0.79, 0.89) | 0.76 (0.69, 0.85) |
|  | ≥10 (2,379,049/5,248) | 0.74 (0.64, 0.84) | 0.73 (0.64, 0.83) | 0.93 (0.87, 0.99) | 0.84 (0.77, 0.91) | 0.79 (0.68, 0.93) |
|  | *p*-heterogeneity¹ | 0.34 | 0.15 | 0.79 | 0.82 | 0.69 |
| **Baseline health status** | Excellent/good (6,350,288/13,994) | 0.80 (0.73, 0.86) | 0.81 (0.74, 0.87) | 0.93 (0.89, 0.97) | 0.88 (0.84, 0.93) | 0.80 (0.73, 0.88) |
|  | Fair/poor (3,950,744/12,372) | 0.78 (0.71, 0.85) | 0.80 (0.74, 0.87) | 0.89 (0.85, 0.93) | 0.89 (0.84, 0.94) | 0.82 (0.74, 0.90) |
|  | *p*-heterogeneity¹ | 0.68 | 0.83 | 0.17 | 0.77 | 0.72 |
| **Frequency of strenuous exercise** | Rare/<1 per wk (5,782,632/15,772) | 0.77 (0.72, 0.84) | 0.82 (0.76, 0.88) | 0.92 (0.88, 0.95) | 0.90 (0.86, 0.95) | 0.81 (0.74, 0.88) |
|  | 1-6 per wk (4,361,583/9,948) | 0.84 (0.76, 0.92) | 0.82 (0.74, 0.90) | 0.92 (0.87, 0.96) | 0.87 (0.82, 0.93) | 0.86 (0.77, 0.96) |
|  | Daily (294,246/873) | 0.68 (0.49, 0.94) | 0.70 (0.51, 0.98) | 0.82 (0.69, 0.96) | 0.83 (0.67, 1.02) | 0.62 (0.43, 0.90) |
|  | *p*-heterogeneity¹ | 0.25 | 0.65 | 0.41 | 0.58 | 0.22 |
| **Menopausal hormone therapy use** | Never (4,838,384/13,690) | 0.82 (0.76, 0.89) | 0.83 (0.76, 0.90) | 0.92 (0.88, 0.95) | 0.89 (0.85, 0.94) | 0.86 (0.78, 0.94) |
|  | Ever (5,616,314/12,917) | 0.75 (0.69, 0.82) | 0.79 (0.73, 0.86) | 0.90 (0.87, 0.94) | 0.89 (0.84, 0.93) | 0.77 (0.70, 0.85) |
|  | *p*-heterogeneity¹ | 0.13 | 0.41 | 0.43 | 1.00 | 0.11 |
| **Deprivation status** | Least deprived (2,511,975/6,081) | 0.77 (0.68, 0.87) | 0.79 (0.70, 0.89) | 0.89 (0.84, 0.95) | 0.84 (0.78, 0.91) | 0.78 (0.68, 0.90) |
|  | Most deprived (1,491,101/4,216) | 0.86 (0.74, 0.99) | 0.89 (0.77, 1.02) | 0.87 (0.81, 0.94) | 0.89 (0.81, 0.98) | 0.85 (0.72, 1.00) |
|  | *p*-heterogeneity¹ | 0.26 | 0.21 | 0.64 | 0.36 | 0.44 |
| **Excluding first five years of follow-up** (7,969,339/25,940) |  | 0.79 (0.74, 0.83) | 0.81 (0.76, 0.85) | 0.91 (0.88, 0.94) | 0.89 (0.85, 0.92) | 0.82 (0.76, 0.87) |

<sup>1</sup> *p*-heterogeneity was calculated with the inverse variance-weighted average of stratum-specific log hazard ratios.
smoker <15 cigarettes/day, current smoker ≥ 15 cigarettes/day, missing), alcohol consumption (<1, 1 to <10, 10 to <20, and ≥ 20g/day), strenuous exercise frequencies (none, <1, 1, 2-3, 4-6, 7 times/week, missing), MHT use (never, past, current, missing), parity (nulliparous, 1-2 children, ≥ 3 children, missing), age at menopause (<40, 40-44, 45-49, 50-54, ≥ 55 years, missing), height (quintiles and missing), family history of hip fractures (yes, no, missing), and BMI (<18.5, 18.5 to <20, 20 to <22.5, 22.5 to <25, 25 to <27.5, 27.5 to <30, 30 to <32.5, ≥ 32.5 kg/m<sup>2</sup>, missing). For analyses of total energy and alcohol, covariates of energy intake and alcohol consumption were taken out of the model respectively.

In the “healthiest” subgroup (N=31,391, 1,771 incident cases), the associations of the top ten foods and nutrients were consistent in direction, but only vegetables, fiber, and pasta remained statistically significant at conventional level (p<0.05, **STable 11**). The results remained consistent after excluding the first five years of follow-up (**Table 3**, **Table 4**, and **STable 12)**.

## Discussion

In the largest prospective cohort study to date including nearly 542,000 women and 27,318 incident hip fractures, we conducted a comprehensive investigation into the associations between 99 dietary factors and hip fracture risk. After adjusting for multiple testing, 60 dietary factors were associated with hip fracture risk. Overall, dietary factors widely thought to be healthy, such as chicken, vegetables, protein, and zinc, were associated with lower risk, while “unhealthy” dietary factors, such as chips, fizzy drinks, SFA, and free sugars, were associated with higher risk. Many of these dietary factors that were significantly associated with hip fracture risk were also highly intercorrelated. Consistent results in sensitivity analyses suggested there was no strong indication of substantial residual confounding or reverse causation.

The diet-wide analysis approach allows us to evaluate 25 dietary factors reported in previous prospective cohort studies and identify 35 new associations. Among the top five foods and top five nutrients (based on p-value) associated with hip fracture risk, only six were reported before, including inverse associations with protein (9), and niacin (30), vegetables (9), carotene (9), fiber (31), and positive associations with fizzy drinks (32), which were generally consistent with our findings except niacin and fiber (for which previous studies have reported null associations). The discussion below focuses on these top foods and nutrients, as well as other dietary factors that were highly correlated with these factors in the current study.

Among the investigated nutrients, protein displayed the strongest inverse association (based on p-value) with hip fracture risk in the current study, which was a 21% lower risk per 15g/day. In line with our findings, a recent meta-analysis (7) of four prospective cohort studies (n=∼174,000; 4,185 cases) reported a pooled RR of 0.52 (95% CI 0.33, 0.82) per 100 g/day. These results for protein also aligned with our observed inverse associations of foods and nutrients which were strongly to moderately correlated with protein, including zinc, phosphorus, vitamin B_6_, thiamin, potassium, magnesium, niacin, selenium, vitamin B_12_, calcium, riboflavin, folate, and chicken. Even though chicken is the food with the strongest association with a lower risk of hip fracture, no previous prospective cohort studies have reported specifically on chicken. Of these nutrients, existing evidence of prospective cohort studies on phosphorus (33), vitamin B_6_, folate, and vitamin B_12_ (34, 35), thiamin and riboflavin (34), magnesium (36), niacin (30, 34), and calcium (37) has been inconsistent, and no studies have been found that investigated zinc, potassium, and selenium with hip fracture risk.

Vegetables was one of the top food groups associated with a lower risk of hip fracture and was not materially associated with either chicken (top 1 food) or protein (top 1 nutrient). We found a 12% lower risk per 100g/day higher intake of vegetables. A meta-analysis (38) of four prospective studies (n=∼330,000; ∼6,000 cases) reported similar results, with an inverse association of highest versus lowest consumption of vegetables (HR 0.81, 95% CI 0.68, 0.96). Carotene intake was highly correlated, and fiber intake was moderately correlated, with vegetable intake in our study. As might be expected from this high correlation, a 9% lower risk was observed per 1,000 µg/day carotene intake, aligning with two smaller cohort studies (39, 40) which reported the same direction of association between carotene intake and hip fracture risk. In contrast to our finding of an inverse association for fiber (11% lower risk per 5g/day), a study in UK Biobank (n=132,807; 930 cases), whose participants were on average 10 years younger than ours, found no association (41), as did a small study of three male cohorts (n=2,879; 72 cases) (31).

Among the foods positively associated with hip fracture risk, fizzy drinks ranked among the top two, with an 18% higher risk per 50g/day. Two large cohorts (each ∼70,000 postmenopausal women; ∼2,000 cases) also found higher hip fracture risk with soft drink consumption (26% higher risk for the highest vs no consumption in the Women’s Health Initiative and 14% higher risk per 355 mL/day in the Nurses’ Health Study) (32, 42). Two smaller US cohorts with 1512 cases and 275 cases respectively have reported directionally consistent but non-significant associations (43, 44).

Retinol was the nutrient most strongly positively associated with hip fracture risk, with a 12% higher risk per 250 µg/day. Similarly, the Nurses’ Health Study reported a 33% higher risk per 500 µg/day dietary retinol intake (45). A non-significant elevated risk associated with total retinol was also observed in the Women’s Health Initiative Observational study (highest vs lowest intake HR 1.13, 95%CI 0.81, 1.59) (46) and the Iowa Women’s Health Study (highest vs the lowest intake HR 1.10, 95%CI 0.84–1.43) (47), though the latter found no association among people who did not use supplements.

Alcohol was the only nutrient that showed little correlation with other nutrients or foods. We observed a 4% lower risk per 10 g/day of alcohol intake overall, but the positive association among regular drinkers in stratified analysis suggested potential residual confounding. Evidence from a Mendelian randomization (MR) analysis in a large Chinese cohort showed null associations between alcohol consumption and hip fracture risk (48), and a UK MR study found null results for total fracture risk (49). Therefore, it is likely the observed inverse association between alcohol intake and hip fracture risk is due to confounding.

Our study found that among dairy-related dietary factors commonly linked to bone health —such as calcium, vitamin D, milk, yogurt and cheese—only calcium and yoghurt were both associated with a 6% lower risk per 300mg/day and per 50g/day, respectively. Previous randomized trials (6, 50) also indicated that higher calcium and dairy intakes only reduced fracture risk in middle-aged and older populations with low baseline calcium intake (<700 mg/day). Recent MR analyses have not supported a causal association between genetically predicted milk intake and hip fracture risk (51), nor between circulating vitamin D and hip fracture risk (52). Further research should explore these associations across different intake levels.

There are several potential mechanisms linking dietary factors to hip fracture risk. First, some dietary factors are directly involved in bone formation, such as protein in collagen matrix formation (53) and magnesium in mineralization through its role in vitamin D activation (54). Second, dietary factors may influence bone resorption. Vegetables and fruits intake provide antioxidants that, in human observational and intervention studies, are associated with lower levels of oxidative stress (55, 56), which has been linked to bone mass loss (10, 57). Third, dietary factors may influence the endocrine pathways that modulate bone metabolism, with protein intake and fiber intake positively associated with insulin-like growth factor-1 which directly promotes osteoblast activities (58). Lastly, some dietary factors, such as protein (59), are suggested to preserve muscle mass and strength, which can help to maintain bone mineral density (60) and decrease the risk of falls (61).

A key observation in this study was that dietary factors widely regarded as “healthy” were generally associated with a lower risk while those considered “unhealthy” were associated with a higher risk. This raises concerns about potential residual confounding by overall diet quality, lifestyle, health and health-related behaviors. Although we took steps to control for confounding through statistical adjustments, stratification, and restriction to the least confounded group, we cannot rule out potential residual confounding. This could result from the imperfect adjustment of confounding variables due to inaccurate measurement from self-reported information, unmeasured confounders, and change of certain key confounders over time, such as BMI, physical activity, and MHT use. Further diet-wide studies in other cohorts with different confounding structures may help validate these findings.

The study’s strengths include being the largest prospective cohort analysis of diet and hip fracture risk to date, with a wide range of dietary factors. We used a 24-hour dietary recall assessment, conducted an average of 10 years after baseline, to calibrate baseline dietary intake from the FFQ, thereby reducing measurement error and providing better measures of usual dietary intake. However, some limitations exist: information on certain dietary factors possibly linked to bone health, such as vitamin K, was unavailable, residual confounding from health behaviors may persist, and the findings have limited generalizability to men or to non-white ethnic groups.

To conclude, in this largest study to date with nearly 542,000 postmenopausal women and over 27,000 incident hip fractures, we found that 60 dietary factors were associated with hip fracture risk. Among them, “healthy” dietary factors, such as chicken, vegetables, and protein, were associated with lower risk, while “unhealthy” dietary factors, such as chips and fizzy drinks were associated with higher risk. Because residual confounding cannot be ruled out, those findings which are most biologically plausible need to be confirmed in other cohorts with different confounding structures and, where feasible, in randomized controlled trials to determine which, if any, dietary factors are suitable targets for hip fracture prevention.

## Data Availability

Data access policies for the Million Women Study are available at the study website [https://www.ceu.ox.ac.uk/research/the-million-women-study].

## Acknowledgments

The authors would like to acknowledge Sau Wan Kan and Kirstin Pirie for their enthusiastic help and suggestions on data processing. We thank the women who participated in the Million Women Study and staff who contributed to the data collection from NHS breast screening centers. This work uses data provided by patients and collected by the NHS as part of their care and support; we thank NHS England and Public Health Scotland for the health outcomes data.

## Authors’ contributions

G.K.R. was involved in the concept, design and data acquisition for the Million Women Study. K.P., T.Y.N.T., and Y.T. developed the project plan. Y.T. analyzed the data, and all authors interpreted the data. Y.T. drafted the first version of the manuscript. All authors contributed to drafting revised versions of the manuscript and gave their final approval of the version to be published.

## Data sharing plan

Data access policies for the Million Women Study are available at the study website [https://www.ceu.ox.ac.uk/research/the-million-women-study]. For the purpose of open access, the authors have applied a Creative Commons Attribution (CC BY) license to any Author Accepted Manuscript version arising.

## Abbreviations

BMI: Body mass index
CI: Confidence interval
FDR: False discovery rate
FFQ: Food frequency questionnaire
HR: Hazard ratio
ICD: International classification of diseases
MHT: Menopausal hormone therapy
MR: Mendelian randomization
NHS: National Health Service
RR: Risk ratio
SFA: Saturated fatty acids

## Reference

1. National Falls and Fragility Fracture Audit Programme. National Hip Fracture Database [cited 2025 December 9, 2025]. Available from: https://www.nhfd.co.uk/20/NHFDcharts.nsf/vwCharts/OverallPerformance.

2. Sing CW, Lin TC, Bartholomew S, Bell JS, Bennett C, Beyene K, et al. Global Epidemiology of Hip Fractures: Secular Trends in Incidence Rate, Post-Fracture Treatment, and All-Cause Mortality. J Bone Miner Res. 2023;38(8):1064–75. Epub 20230529. doi: 10.1002/jbmr.4821. PubMed PMID: 37118993.

3. Webster J, Oguzman E, Morris EJA, Shepperd S, Griffin XL, Johansen A, et al. Trends and variation in the incidence of hip fracture in England before, during, and after the COVID-19 pandemic (2014–2024): a population-based observational study. The Lancet Regional Health – Europe. 2025;57:101427. doi: 10.1016/j.lanepe.2025.101427.

4. National Hip Fracture Database. A broken hip – three steps to recovery--The 2024 National Hip Fracture Database report on 2023. Falls and Fragility Fracture Audit Programme,, 2024.

5. Yao P, Bennett D, Mafham M, Lin X, Chen Z, Armitage J, et al. Vitamin D and Calcium for the Prevention of Fracture: A Systematic Review and Meta-analysis. JAMA Netw Open. 2019;2(12):e1917789. Epub 20191202. doi: 10.1001/jamanetworkopen.2019.17789. PubMed PMID: 31860103; PubMed Central PMCID: PMC6991219.

6. Iuliano S, Poon S, Robbins J, Bui M, Wang X, De Groot L, et al. Effect of dietary sources of calcium and protein on hip fractures and falls in older adults in residential care: cluster randomised controlled trial. Bmj. 2021;375:n2364. Epub 20211020. doi: 10.1136/bmj.n2364. PubMed PMID: 34670754; PubMed Central PMCID: PMC8527562.

7. Zeraattalab-Motlagh S, Mortazavi AS, Ghoreishy SM, Mohammadi H. Association between total and animal proteins with risk of fracture: A systematic review and dose-response meta-analysis of cohort studies. Osteoporos Int. 2024;35(1):11–23. Epub 20231019. doi: 10.1007/s00198-023-06948-8. PubMed PMID: 37855886.

8. Brondani JE, Comim FV, Flores LM, Martini LA, Premaor MO. Fruit and vegetable intake and bones: A systematic review and meta-analysis. PLoS One. 2019;14(5):e0217223. Epub 20190531. doi: 10.1371/journal.pone.0217223. PubMed PMID: 31150426; PubMed Central PMCID: PMC6544223.

9. Webster J, Rycroft CE, Greenwood DC, Cade JE. Dietary risk factors for hip fracture in adults: An umbrella review of meta-analyses of prospective cohort studies. PLoS One. 2021;16(11):e0259144. Epub 20211110. doi: 10.1371/journal.pone.0259144. PubMed PMID: 34758048; PubMed Central PMCID: PMC8580223.

10. Nieves JW. Chapter 71 – Nutrients beyond calcium and vitamin D to treat osteoporosis. In: Dempster DW, Cauley JA, Bouxsein ML, Cosman F, editors. Marcus and Feldman’s Osteoporosis (Fifth Edition): Academic Press; 2021. p. 1679–93.

11. Mishra S, Baruah K, Malik VS, Ding EL. Dairy intake and risk of hip fracture in prospective cohort studies: non-linear algorithmic dose-response analysis in 486 950 adults. Journal of Nutritional Science. 2023;12:e96. Epub 2023/09/11. doi: 10.1017/jns.2023.63.

12. Green J, Reeves GK, Floud S, Barnes I, Cairns BJ, Gathani T, et al. Cohort Profile: the Million Women Study. Int J Epidemiol. 2019;48(1):28–9e. doi: 10.1093/ije/dyy065. PubMed PMID: 29873753; PubMed Central PMCID: PMC6380310.

13. Unit. CE. Million Women Study In: Oxford. Uo, editor. 1996.

14. The Million Women Study [cited 2024 August 08]. Available from: https://www.ceu.ox.ac.uk/research/the-million-women-study.

15. The Million Women study 3-year re-survey questionnaire [cited 2024 August 08]. Available from: https://www.ceu.ox.ac.uk/files/about/mws-q3.pdf.

16. Roddam AW, Spencer E, Banks E, Beral V, Reeves G, Appleby P, et al. Reproducibility of a short semi-quantitative food group questionnaire and its performance in estimating nutrient intake compared with a 7-day diet diary in the Million Women Study. Public Health Nutr. 2005;8(2):201–13. doi: 10.1079/phn2004676. PubMed PMID: 15877913.

17. Holland B, Welch, A.A., Unwin, I.D., Buss, D.H., Paul, A.A. and Southgate, D.A.T. McCance and Widdowson’s The composition of foods. 5th ed. Cambridge, UK: Royal Society of Chemistry; 1991.

18. Perez-Cornago A, Pollard Z, Young H, van Uden M, Andrews C, Piernas C, et al. Description of the updated nutrition calculation of the Oxford WebQ questionnaire and comparison with the previous version among 207,144 participants in UK Biobank. Eur J Nutr. 2021;60(7):4019–30. Epub 20210506. doi: 10.1007/s00394-021-02558-4. PubMed PMID: 33956230; PubMed Central PMCID: PMC8437868.

19. Liu B, Young H, Crowe FL, Benson VS, Spencer EA, Key TJ, et al. Development and evaluation of the Oxford WebQ, a low-cost, web-based method for assessment of previous 24 h dietary intakes in large-scale prospective studies. Public Health Nutr. 2011;14(11):1998–2005. Epub 20110601. doi: 10.1017/s1368980011000942. PubMed PMID: 21729481.

20. Greenwood DC, Hardie LJ, Frost GS, Alwan NA, Bradbury KE, Carter M, et al. Validation of the Oxford WebQ Online 24-Hour Dietary Questionnaire Using Biomarkers. Am J Epidemiol. 2019;188(10):1858–67. doi: 10.1093/aje/kwz165. PubMed PMID: 31318012; PubMed Central PMCID: PMC7254925.

21. Townsend P, Phillimore P, Beattie A. Health and deprivation: inequality and the North: Routledge; 2023.

22. Albright F, Smith PH, Richardson AM. POSTMENOPAUSAL OSTEOPOROSIS: ITS CLINICAL FEATURES. Journal of the American Medical Association. 1941;116(22):2465–74. doi: 10.1001/jama.1941.02820220007002.

23. Armstrong ME, Lacombe J, Wotton CJ, Cairns BJ, Green J, Floud S, et al. The Associations Between Seven Different Types of Physical Activity and the Incidence of Fracture at Seven Sites in Healthy Postmenopausal UK Women. J Bone Miner Res. 2020;35(2):277–90. Epub 20191115. doi: 10.1002/jbmr.3896. PubMed PMID: 31618477; PubMed Central PMCID: PMC7027536.

24. MacMahon S, Peto R, Cutler J, Collins R, Sorlie P, Neaton J, et al. Blood pressure, stroke, and coronary heart disease. Part 1, Prolonged differences in blood pressure: prospective observational studies corrected for the regression dilution bias. Lancet. 1990;335(8692):765–74. doi: 10.1016/0140-6736(90)90878-9. PubMed PMID: 1969518.

25. Key TJ, Balkwill A, Bradbury KE, Reeves GK, Kuan AS, Simpson RF, et al. Foods, macronutrients and breast cancer risk in postmenopausal women: a large UK cohort. Int J Epidemiol. 2019;48(2):489–500. doi: 10.1093/ije/dyy238. PubMed PMID: 30412247; PubMed Central PMCID: PMC6469308.

26. Papier K, Bradbury KE, Balkwill A, Barnes I, Smith-Byrne K, Gunter MJ, et al. Diet-wide analyses for risk of colorectal cancer: prospective study of 12,251 incident cases among 542,778 women in the UK. Nat Commun. 2025;16(1):375. Epub 20250108. doi: 10.1038/s41467-024-55219-5. PubMed PMID: 39779669.

27. Papier K, Fensom GK, Knuppel A, Appleby PN, Tong TYN, Schmidt JA, et al. Meat consumption and risk of 25 common conditions: outcome-wide analyses in 475,000 men and women in the UK Biobank study. BMC Med. 2021;19(1):53. Epub 20210302. doi: 10.1186/s12916-021-01922-9. PubMed PMID: 33648505; PubMed Central PMCID: PMC7923515.

28. Benjamini Y, Hochberg Y. Controlling the False Discovery Rate: A Practical and Powerful Approach to Multiple Testing. Journal of the Royal Statistical Society: Series B (Methodological). 1995;57(1):289–300. doi: 10.1111/j.2517-6161.1995.tb02031.x.

29. Unnanuntana A, Rebolledo BJ, Khair MM, DiCarlo EF, Lane JM. Diseases affecting bone quality: beyond osteoporosis. Clin Orthop Relat Res. 2011;469(8):2194–206. doi: 10.1007/s11999-010-1694-9. PubMed PMID: 21107923; PubMed Central PMCID: PMC3126973.

30. Carbone LD, Bůžková P, Fink HA, Raiford M, Le B, Isales CM, et al. Association of Dietary Niacin Intake With Incident Hip Fracture, BMD, and Body Composition: The Cardiovascular Health Study. J Bone Miner Res. 2019;34(4):643–52. Epub 20190119. doi: 10.1002/jbmr.3639. PubMed PMID: 30659655; PubMed Central PMCID: PMC6663556.

31. Dai Z, Hirani V, Sahni S, Felson DT, Naganathan V, Blyth F, et al. Association of dietary fiber and risk of hip fracture in men from the Framingham Osteoporosis Study and the Concord Health and Ageing in Men Project. Nutr Health. 2022;28(2):229–38. Epub 20210504. doi: 10.1177/02601060211011798. PubMed PMID: 33940973; PubMed Central PMCID: PMC10622166.

32. Fung TT, Arasaratnam MH, Grodstein F, Katz JN, Rosner B, Willett WC, et al. Soda consumption and risk of hip fractures in postmenopausal women in the Nurses’ Health Study. Am J Clin Nutr. 2014;100(3):953–8. Epub 20140806. doi: 10.3945/ajcn.114.083352. PubMed PMID: 25099544; PubMed Central PMCID: PMC4135502.

33. Huang Z, Himes JH, McGovern PG. Nutrition and subsequent hip fracture risk among a national cohort of white women. Am J Epidemiol. 1996;144(2):124–34. doi: 10.1093/oxfordjournals.aje.a008899. PubMed PMID: 8678043.

34. Dai Z, Wang R, Ang LW, Yuan JM, Koh WP. Dietary B vitamin intake and risk of hip fracture: the Singapore Chinese Health Study. Osteoporos Int. 2013;24(7):2049–59. Epub 20121213. doi: 10.1007/s00198-012-2233-1. PubMed PMID: 23238962; PubMed Central PMCID: PMC9254692.

35. Meyer HE, Willett WC, Fung TT, Holvik K, Feskanich D. Association of High Intakes of Vitamins B6 and B12 From Food and Supplements With Risk of Hip Fracture Among Postmenopausal Women in the Nurses’ Health Study. JAMA Netw Open. 2019;2(5):e193591. Epub 20190503. doi: 10.1001/jamanetworkopen.2019.3591. PubMed PMID: 31074816; PubMed Central PMCID: PMC6512300.

36. Orchard TS, Larson JC, Alghothani N, Bout-Tabaku S, Cauley JA, Chen Z, et al. Magnesium intake, bone mineral density, and fractures: results from the Women’s Health Initiative Observational Study. Am J Clin Nutr. 2014;99(4):926–33. Epub 20140205. doi: 10.3945/ajcn.113.067488. PubMed PMID: 24500155; PubMed Central PMCID: PMC3953885.

37. Bischoff-Ferrari HA, Dawson-Hughes B, Baron JA, Burckhardt P, Li R, Spiegelman D, et al. Calcium intake and hip fracture risk in men and women: a meta-analysis of prospective cohort studies and randomized controlled trials. Am J Clin Nutr. 2007;86(6):1780–90. doi: 10.1093/ajcn/86.5.1780. PubMed PMID: 18065599.

38. Luo S, Li Y, Luo H, Yin X, Lin du R, Zhao K, et al. Increased intake of vegetables, but not fruits, may be associated with reduced risk of hip fracture: A meta-analysis. Sci Rep. 2016;6:19783. Epub 20160125. doi: 10.1038/srep19783. PubMed PMID: 26806285; PubMed Central PMCID: PMC4726403.

39. Sahni S, Hannan MT, Blumberg J, Cupples LA, Kiel DP, Tucker KL. Protective effect of total carotenoid and lycopene intake on the risk of hip fracture: a 17-year follow-up from the Framingham Osteoporosis Study. J Bone Miner Res. 2009;24(6):1086–94. doi: 10.1359/jbmr.090102. PubMed PMID: 19138129; PubMed Central PMCID: PMC2683648.

40. Dai Z, Wang R, Ang LW, Low YL, Yuan JM, Koh WP. Protective effects of dietary carotenoids on risk of hip fracture in men: the Singapore Chinese Health Study. J Bone Miner Res. 2014;29(2):408–17. doi: 10.1002/jbmr.2041. PubMed PMID: 23857780; PubMed Central PMCID: PMC3894263.

41. Yang H, Jiang Y, Luo Y, Qin K, Yang C, Liang D, et al. Associations of protein intake with the risk of fractures: A prospective cohort study of UK biobank participants. Arch Gerontol Geriatr. 2025;133:105805. Epub 20250225. doi: 10.1016/j.archger.2025.105805. PubMed PMID: 40086418.

42. Kremer PA, Laughlin GA, Shadyab AH, Crandall CJ, Masaki K, Orchard T, et al. Association between soft drink consumption and osteoporotic fractures among postmenopausal women: the Women’s Health Initiative. Menopause. 2019;26(11):1234–41. doi: 10.1097/gme.0000000000001389. PubMed PMID: 31613830.

43. Hansen SA, Folsom AR, Kushi LH, Sellers TA. Association of fractures with caffeine and alcohol in postmenopausal women: the Iowa Women’s Health Study. Public Health Nutr. 2000;3(3):253–61. doi: 10.1017/s136898000000029x. PubMed PMID: 10979145.

44. White SC, Atchison KA, Gornbein JA, Nattiv A, Paganini-Hill A, Service SK. Risk factors for fractures in older men and women: The Leisure World Cohort Study. Gend Med. 2006;3(2):110–23. doi: 10.1016/s1550-8579(06)80200-7. PubMed PMID: 16860270.

45. Feskanich D, Singh V, Willett WC, Colditz GA. Vitamin A intake and hip fractures among postmenopausal women. Jama. 2002;287(1):47–54. doi: 10.1001/jama.287.1.47. PubMed PMID: 11754708.

46. Caire-Juvera G, Ritenbaugh C, Wactawski-Wende J, Snetselaar LG, Chen Z. Vitamin A and retinol intakes and the risk of fractures among participants of the Women’s Health Initiative Observational Study. Am J Clin Nutr. 2009;89(1):323–30. Epub 20081203. doi: 10.3945/ajcn.2008.26451. PubMed PMID: 19056568; PubMed Central PMCID: PMC2715292.

47. Lim LS, Harnack LJ, Lazovich D, Folsom AR. Vitamin A intake and the risk of hip fracture in postmenopausal women: the Iowa Women’s Health Study. Osteoporos Int. 2004;15(7):552–9. doi: 10.1007/s00198-003-1577-y. PubMed PMID: 14760518; PubMed Central PMCID: PMC2020807.

48. Im PK, Wright N, Yang L, Chan KH, Chen Y, Guo Y, et al. Alcohol consumption and risks of more than 200 diseases in Chinese men. Nat Med. 2023;29(6):1476–86. Epub 20230608. doi: 10.1038/s41591-023-02383-8. PubMed PMID: 37291211; PubMed Central PMCID: PMC10287564.

49. Yuan S, Michaëlsson K, Wan Z, Larsson SC. Associations of Smoking and Alcohol and Coffee Intake with Fracture and Bone Mineral Density: A Mendelian Randomization Study. Calcif Tissue Int. 2019;105(6):582–8. Epub 20190904. doi: 10.1007/s00223-019-00606-0. PubMed PMID: 31482193.

50. Tang BM, Eslick GD, Nowson C, Smith C, Bensoussan A. Use of calcium or calcium in combination with vitamin D supplementation to prevent fractures and bone loss in people aged 50 years and older: a meta-analysis. Lancet. 2007;370(9588):657–66. doi: 10.1016/s0140-6736(07)61342-7. PubMed PMID: 17720017.

51. Bergholdt HKM, Larsen MK, Varbo A, Nordestgaard BG, Ellervik C. Lactase persistence, milk intake, hip fracture and bone mineral density: a study of 97 811 Danish individuals and a meta-analysis. J Intern Med. 2018;284(3):254–69. Epub 20180410. doi: 10.1111/joim.12753. PubMed PMID: 29537719.

52. Fang A, Zhao Y, Yang P, Zhang X, Giovannucci EL. Vitamin D and human health: evidence from Mendelian randomization studies. Eur J Epidemiol. 2024;39(5):467–90. Epub 20240112. doi: 10.1007/s10654-023-01075-4. PubMed PMID: 38214845.

53. Shaw G, Lee-Barthel A, Ross ML, Wang B, Baar K. Vitamin C-enriched gelatin supplementation before intermittent activity augments collagen synthesis. Am J Clin Nutr. 2017;105(1):136–43. Epub 20161116. doi: 10.3945/ajcn.116.138594. PubMed PMID: 27852613; PubMed Central PMCID: PMC5183725.

54. Uwitonze AM, Razzaque MS. Role of Magnesium in Vitamin D Activation and Function. J Am Osteopath Assoc. 2018;118(3):181–9. doi: 10.7556/jaoa.2018.037. PubMed PMID: 29480918.

55. Thompson HJ, Heimendinger J, Sedlacek S, Haegele A, Diker A, O’Neill C, et al. 8-Isoprostane F2α excretion is reduced in women by increased vegetable and fruit intake2. The American Journal of Clinical Nutrition. 2005;82(4):768–76. doi: 10.1093/ajcn/82.4.768.

56. Bacchetti T, Turco I, Urbano A, Morresi C, Ferretti G. Relationship of fruit and vegetable intake to dietary antioxidant capacity and markers of oxidative stress: A sex-related study. Nutrition. 2019;61:164–72. doi: 10.1016/j.nut.2018.10.034.

57. Manolagas SC. From Estrogen-Centric to Aging and Oxidative Stress: A Revised Perspective of the Pathogenesis of Osteoporosis. Endocrine Reviews. 2010;31(3):266–300. doi: 10.1210/er.2009-0024.

58. Watling CZ, Kelly RK, Tong TYN, Piernas C, Watts EL, Tin Tin S, et al. Associations of circulating insulin-like growth factor-I with intake of dietary proteins and other macronutrients. Clin Nutr. 2021;40(7):4685–93. Epub 20210420. doi: 10.1016/j.clnu.2021.04.021. PubMed PMID: 34237695; PubMed Central PMCID: PMC8345002.

59. Houston DK, Nicklas BJ, Ding J, Harris TB, Tylavsky FA, Newman AB, et al. Dietary protein intake is associated with lean mass change in older, community-dwelling adults: the Health, Aging, and Body Composition (Health ABC) Study. Am J Clin Nutr. 2008;87(1):150–5. doi: 10.1093/ajcn/87.1.150. PubMed PMID: 18175749.

60. Gandham A, Vandenput L, Turbic A, MacRae M, Nassar H, Kolterman OG, et al. Associations between muscle mass and strength and bone microarchitecture in Caucasian postmenopausal women. Osteoporosis International. 2025;36(10):2031–9. doi: 10.1007/s00198-025-07653-4.

61. Chao YP, Fang WH, Peng TC, Wu LW, Yang HF, Kao TW. Longitudinal Cohort Study Investigating Fall Risk Across Diverse Muscle Health Statuses Among Older People in the Community. J Cachexia Sarcopenia Muscle. 2025;16(2):e13788. doi: 10.1002/jcsm.13788. PubMed PMID: 40162590; PubMed Central PMCID: PMC11955838.

