## Supplemental figures and tables for "Diet-wide association study of foods and nutrients with hip fracture risk: a prospective cohort study of 27,318 incident cases among 541,887 postmenopausal women"

### Supplementary Figures

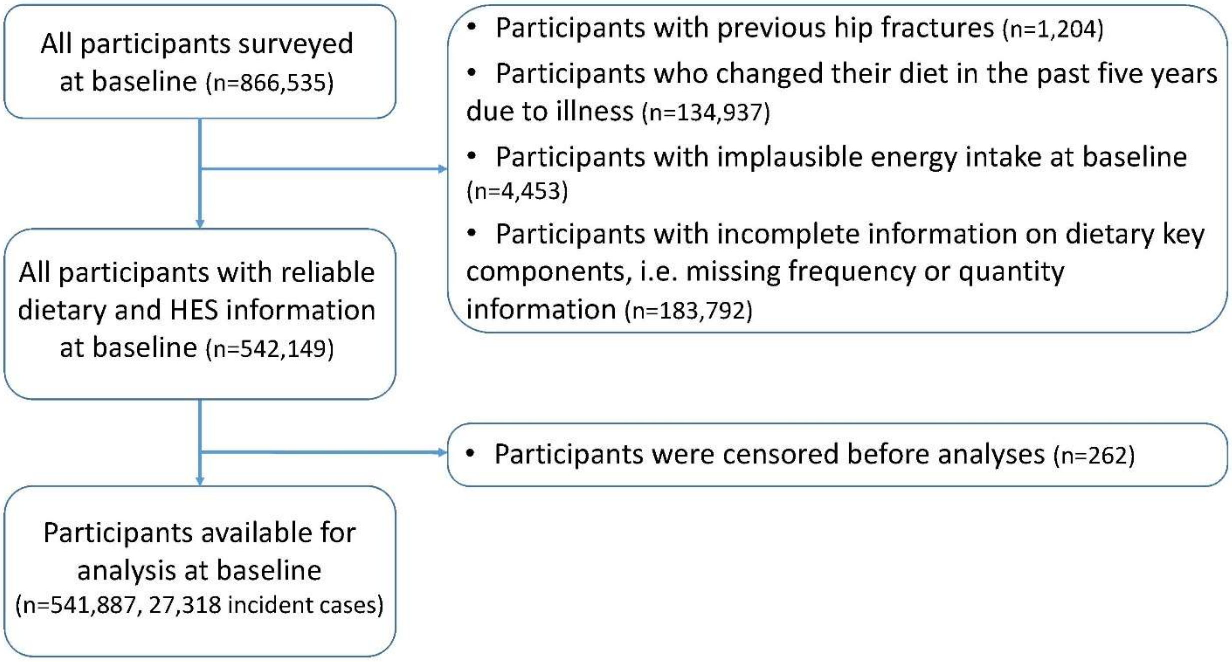

### SFigure 1. Flowchart of participants of the study

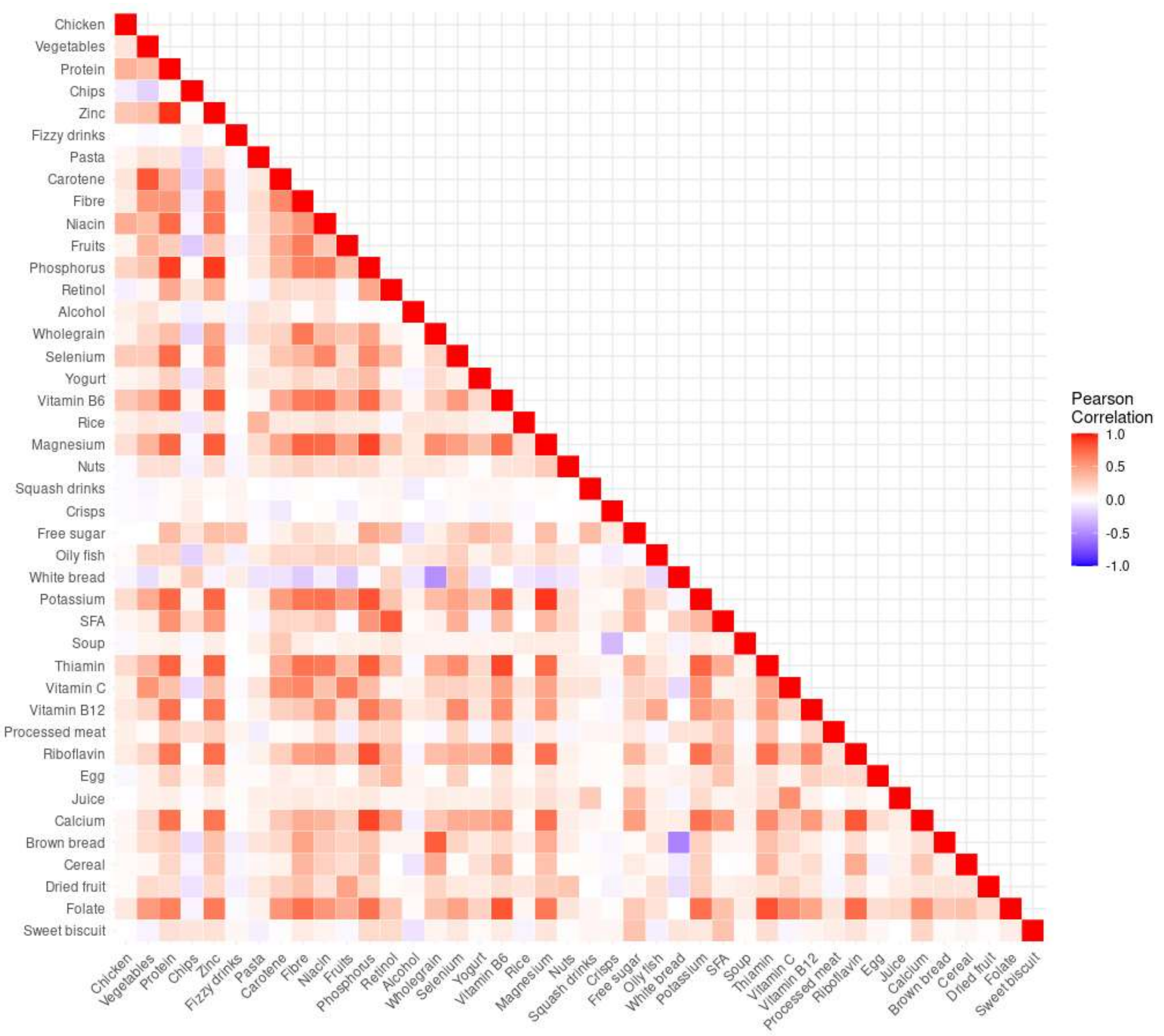

**SFigure 2. Heat map of correlations among significant dietary factors**

Note: SFA -- saturated fatty acids

### Supplementary Tables

### STable 1. Dietary factors and their increments

| Dietary Factors | Incre-ment | Baseline Mean Intake (N=541,887) | | | Resurvey Mean Intake (N=39,616) | | |
|---|---|---|---|---|---|---|---|
| | | Lowest[1] | Highest[1] | Mean | Lowest[1] | Highest[1] | Mean |
| **36 Foods with re-surveyed data for validation** | | | | | | | |
| Fruit (excluding juice) (g/day) | 200 | 33.2 | 325 | 168 | 144 | 310 | 246 |
| Dried Fruit (g/day) | 10 | 0.0 | 68.7 | 16.5 | 6.2 | 15.3 | 9.5 |
| Vegetables (g/day) | 100 | 73.5 | 482 | 241 | 184 | 308 | 257 |
| Milk (g/day) | 200 | 74.6 | 426 | 241 | 109 | 280 | 197 |
| Yoghurt (g/day) | 50 | 9.7 | 143 | 62.5 | 46.2 | 96.8 | 65.7 |
| Cheese (g/day) | 10 | 3.2 | 29.8 | 14.7 | 14.9 | 25.5 | 20.2 |
| Red & Processed Meat (g/day) | 30 | 1.9 | 78.0 | 36.6 | 25.6 | 62.8 | 47.1 |
| Processed Red Meat (g/day) | 10 | 0.0 | 26.1 | 9.6 | 9.2 | 17.7 | 12.7 |
| Red Meat (g/day) | 20 | 0.0 | 63.2 | 27.1 | 21.0 | 46.1 | 34.4 |
| Chicken (g/day) | 20 | 3.3 | 47.4 | 20.4 | 18.0 | 34.9 | 29.0 |
| Oily Fish (g/day) | 10 | 0.0 | 36.4 | 11.4 | 9.0 | 19.1 | 13.1 |
| Non-Oily Fish (g/day) | 10 | 2.8 | 46.0 | 21.7 | 12.3 | 19.8 | 16.5 |
| Eggs (g/day) | 10 | 4.7 | 43.2 | 21.1 | 14.3 | 26.7 | 19.7 |
| Nuts (g/day) | 10 | 0.0 | 22.6 | 5.6 | 4.1 | 10.2 | 6.5 |
| Wholegrains (g/day) | 20 | 7.3 | 63.4 | 33.9 | 15.9 | 29.8 | 24.1 |
| Chips (g/day) | 25 | 0.0 | 60.8 | 17.1 | 22.6 | 47.7 | 28.5 |
| Potatoes (g/day) | 40 | 26.4 | 129 | 75.6 | 67.0 | 107 | 83.7 |
| Pasta (g/day) | 20 | 0.0 | 80.9 | 38.9 | 12.7 | 27.6 | 22.7 |
| Rice (g/day) | 10 | 0.0 | 51.3 | 19.6 | 7.5 | 20.6 | 14.3 |
| White Bread (g/day) | 10 | 0.0 | 125 | 30.2 | 1.9 | 11.0 | 4.1 |
| Brown Bread (g/day) | 10 | 0.0 | 123 | 40.8 | 4.8 | 13.1 | 8.4 |
| Crispbread (g/day) | 10 | 0.0 | 36.0 | 10.0 | 6.3 | 15.7 | 9.1 |
| Sweet Biscuits (g/day) | 10 | 0.0 | 34.6 | 11.4 | 4.6 | 12.7 | 6.9 |
| Cakes (g/day) | 10 | 0.0 | 45.4 | 20.2 | 5.6 | 12.4 | 8.5 |
| Cereal (g/day) | 40 | 30.4 | 122 | 52.8 | 55.8 | 97.6 | 69.5 |
| Crisps (g/day) | 10 | 0.0 | 25.3 | 1.5 | 4.6 | 12.1 | 4.9 |
| Boiled Sweets (g/day) | 5 | 0.0 | 69.6 | 3.5 | 2.0 | 6.0 | 2.1 |
| Jam (g/day) | 5 | 0.0 | 12.7 | 1.3 | 4.3 | 6.8 | 4.5 |
| Chocolate (g/day) | 10 | 1.2 | 65.0 | 20.1 | 5.1 | 12.0 | 7.4 |
| Soup (g/day) | 40 | 0.0 | 137 | 36.7 | 35.2 | 78.3 | 48.0 |
| Sauce (g/day) | 10 | 2.1 | 79.7 | 34.9 | 4.9 | 10.7 | 7.5 |
| Tea (g/day) | 500 | 66.7 | 1391 | 643 | 222 | 863 | 534 |
| Coffee (g/day) | 500 | 95.4 | 1348 | 383 | 181 | 727 | 337 |
| Fruit Juice (g/day) | 100 | 3.1 | 434 | 139 | 43.3 | 155 | 86.6 |
| Fizzy Drinks (g/day) | 50 | 0.0 | 899 | 96.4 | 18.8 | 63.1 | 23.1 |

| | | | | | | | |
|---|---|---|---|---|---|---|---|
| Squash Drinks (g/day) | 50 | 0.0 | 690 | 99.0 | 24.1 | 87.0 | 32.0 |
| **28 Nutrients with re-surveyed data for validation** | | | | | | | |
| Alcohol (g/day) | 10 | 0.0 | 22.1 | 6.2 | 1.5 | 27.4 | 12.1 |
| Energy (kj/day) | 1000 | 4688 | 9553 | 6981 | 7214 | 8813 | 8039 |
| Protein (g/day) | 15 | 44.0 | 91.7 | 66.8 | 67.8 | 82.6 | 76.4 |
| Total Fat (g/day) | 20 | 36.2 | 96.2 | 63.6 | 61.4 | 81.4 | 71.6 |
| SFA (g/day) | 10 | 10.8 | 37.9 | 22.4 | 22.0 | 31.8 | 26.8 |
| PUFA (g/day) | 5 | 6.2 | 19.0 | 11.8 | 10.6 | 13.2 | 12.0 |
| MUFA (g/day) | 5 | 12.7 | 35.7 | 23.2 | 22.4 | 29.2 | 26.2 |
| Carbohydrate (g/day) | 50 | 132 | 291 | 207 | 202 | 265 | 234 |
| Total Sugars (g/day) | 50 | 52.8 | 157.3 | 99.8 | 100.3 | 143.4 | 121.4 |
| Free Sugars (g/day) | 30 | 20.2 | 107.4 | 57.2 | 39.4 | 66.0 | 50.2 |
| Total Fiber (g/day) | 5 | 7.8 | 20.8 | 13.9 | 15.0 | 20.8 | 18.3 |
| Calcium (mg/day) | 300 | 492 | 1252 | 842 | 818 | 1113 | 973 |
| Folate (µg/day) | 100 | 160 | 354 | 251 | 265 | 345 | 309 |
| Retinol (µg/day) | 250 | 155 | 958 | 423 | 384 | 612 | 471 |
| Carotene (µg/day) | 1000 | 579 | 2492 | 1378 | 2652 | 4037 | 3475 |
| Vitamin D (µg/day) | 1 | 1.3 | 5.7 | 3.1 | 3.0 | 4.1 | 3.6 |
| Vitamin E (mg/day) | 2 | 4.8 | 15.4 | 9.3 | 10.3 | 11.8 | 11.2 |
| Thiamin (mg/day) | 1 | 0.8 | 1.8 | 1.3 | 1.6 | 2.0 | 1.8 |
| Riboflavin (mg/day) | 1 | 1.1 | 2.6 | 1.8 | 1.6 | 2.1 | 1.9 |
| Niacin (mg/day) | 10 | 11.4 | 26.8 | 18.7 | 31.4 | 38.0 | 35.3 |
| Vitamin B6 (mg/day) | 1 | 1.2 | 2.5 | 1.8 | 1.7 | 2.1 | 1.9 |
| Vitamin B12 (µg/day) | 2 | 2.3 | 8.0 | 4.6 | 5.2 | 7.0 | 6.3 |
| Vitamin C (mg/day) | 100 | 40.5 | 168 | 96.9 | 100 | 167 | 139 |
| Magnesium (mg/day) | 100 | 197 | 403 | 295 | 285 | 356 | 324 |
| Potassium (mg/day) | 1000 | 2236 | 4457 | 3298 | 3281 | 4094 | 3726 |
| Zinc (mg/day) | 2 | 4.8 | 10.6 | 7.6 | 8.2 | 10.1 | 9.3 |
| Phosphorus (mg/day) | 300 | 814 | 1707 | 1238 | 1223 | 1519 | 1387 |
| Selenium (mg/day) | 10 | 29.5 | 74.7 | 49.9 | 45.5 | 55.4 | 50.9 |
| **35 Foods with baseline binary classifications** | | | | | | | |
| Apples, Bananas, Grapefruit, Oranges, Pears, Stone Fruit, Stewed Fruit, Carrot, Courgettes, Beetroot, Parsnip, Lettuce, Tomatoes, Swede, Spinach, Sweetcorn, Avocado, Celery, Green Beans, Green / Red Peppers, Mushrooms, Cucumbers, Broccoli, Cabbage, Cauliflower, Brussel Sprouts, Onion, Garlic, Leeks, Green Peas, Baked Beans, Chick Peas/ Lentils, Soya/Tofu, Soya Milk, Ice Cream | | | | | | | |

[1] Based on baseline categories

Abbreviation: SFA saturated fatty acids, MUFA mono-unsaturated fatty acids, PUFA poly-unsaturated fatty acids.

Note: The baseline categories for dietary factors with quantitative intake data was further described and grouped as follows based on the distribution of the data. **Fruit (excluding juice)** (fresh fruit, dried fruit, and stewed or tinned fruit) is grouped as ≤5, 6-9, 10-14, 15-19, 20+ serving per week (pw). **Dried fruit** (prunes, raisins, etc.) is grouped as <1, 1-2, 3+ servings pw. **Vegetables** (26 individual vegetables and legumes but not potatoes) is grouped as <10, 10-14, 15-19, 20-29, 30+ servings pw. **Milk** (full cream milk, semi-skimmed milk, and skimmed milk people drink and used in milky drinks, breakfast cereal, and tea /

coffee) is divided into fifth. **Yoghurt** (plain and flavoured yoghurt and milk-based desserts) is grouped as ≤2, 3-4, 5-6, 7+ times pw. **Cheese** is grouped as ≤1, 2, 3, 4+ times pw. **Processed meat** (sausages, bacon, and ham), **red meat** (beef, pork, and lamb), **red and processed meat** (red meat and processed meat), **chicken** (chicken and other poultry), **oily fish** (sardines, salmon, trout, kippers, and mackerel) and **non-oily fish** (fish and chips, tuna, white fish, and seafood) are divided into fifth. **Egg** is grouped as ≤1, 2, 3, 4+ whole egg pw. **Nuts** (including peanut butter) is grouped as <1, 1, 2+ tablespoon (Tbs) pw. **Wholegrains** (brown/ wholemeal type of the following foods: pasta, rice, bread slice, crackers/crispbread, sweet biscuits, cake/puddings; and bran cereal, biscuit cereal, oat cereal, muesli, and other cereal types) is divided into fifth. **Chips, pasta** (white and brown), **rice** (white and brown) are grouped as 0, 1, 2+ times pw. **Potatoes** (fried, boiled, and mashed potatoes) is grouped as <3, 3, 4, 5, 6+ times pw. **White bread** and **brown bread** are grouped as 0, 1-9, 10-19, 20+ slices pw. **Crackers/crispbreads** is grouped as 0, 1-6, 7+ number pw. **Sweet biscuits** is grouped as 0, 1-9, 10+ number pw. **Cakes** (cakes / puddings / pies / buns) is grouped as 0, 1-2, 3+ number pw. **Breakfast cereal** is grouped as <7, 7, 8+ bowls pw. **Crisps** is grouped as 0, 1, 2, 3+ packets pw. **Boiled sweets** is grouped as 0, 1, 2-6, 7+ number pw. **Jam** is grouped as 0, 1, 2, 3+ Tbs pw. **Chocolate** is grouped as ≤1, 2-3, 4-5, 6+ pieces pw. **Soup** is grouped as 0, 1, 2, 3+ bowls pw. **Sauce** (gravy / cream / cheese sauces) is grouped as ≤1, 2-3, 4-5, 6+ Tbs pw. **Tea** is grouped as 0-1, 2-3, 4-5, 6+ cups per day (pd). **Coffee** is grouped as 0-1, 2-3, 4-5, 6+ cups pd. **Fruit juice** is grouped as <2, 2-4, 5-7, 8+ glasses pw. **Fizzy/soft drinks** is grouped as 0, 1, 2+ glasses pd. **Fruit squash** is grouped as 0, 1, 2+ glasses pd. **Alcohol** is grouped as 0, 1-5, 6-10, 11+ drinks pw. Other nutrients are divided into fifths.

### STable 2. Baseline characteristics by incident hip fractures

| | Non-cases (N=514,569) | Cases (N=27,318) | Total (N=541,887) |
|---|---|---|---|
| **Socioeconomic** | | | |
| **Ethnicity** | | | |
| White | 505,339 (98.2%) | 26,847 (98.3%) | 532,186 (98.2%) |
| non-White | 3,737 (0.7%) | 109 (0.4%) | 3,846 (0.7%) |
| Missing | 5,493 (1.1%) | 362 (1.3%) | 5,855 (1.1%) |
| **Townsend deprivation index** | | | |
| Q1, least deprived | 119,957 (23.3%) | 6,081 (22.3%) | 126,038 (23.3%) |
| Q2 | 114,393 (22.2%) | 5,970 (21.9%) | 120,363 (22.2%) |
| Q3 | 106,530 (20.7%) | 5,723 (20.9%) | 112,253 (20.7%) |
| Q4 | 95,746 (18.6%) | 5,137 (18.8%) | 100,883 (18.6%) |
| Q5, most deprived | 74,094 (14.4%) | 4,216 (15.4%) | 78,310 (14.5%) |
| Missing | 3,849 (0.7%) | 191 (0.7%) | 4,040 (0.7%) |
| **Education & qualification** | | | |
| Tertiary | 88,066 (17.1%) | 4,268 (15.6%) | 92,334 (17.0%) |
| Secondary | 162,279 (31.5%) | 7,824 (28.6%) | 170,103 (31.4%) |
| Technical | 86,962 (16.9%) | 4,752 (17.4%) | 91,714 (16.9%) |
| No qualification | 169,290 (32.9%) | 9,969 (36.5%) | 179,259 (33.1%) |
| Missing | 7,972 (1.5%) | 505 (1.8%) | 8,477 (1.6%) |
| **Lifestyle** | | | |
| **Smoking status** | | | |
| Never | 285,249 (55.4%) | 14,981 (54.8%) | 300,230 (55.4%) |
| Former | 169,133 (32.9%) | 8,447 (30.9%) | 177,580 (32.8%) |
| Current smoker <15 cigarettes/day | 26,223 (5.1%) | 1,662 (6.1%) | 27,885 (5.1%) |
| Current smoker ≥15 cigarettes/day | 33,823 (6.6%) | 2,221 (8.1%) | 36,044 (6.7%) |
| Missing | 141 (0.0%) | 7 (0.0%) | 148 (0.0%) |
| **Alcohol consumption** | | | |
| Non-drinker | 175,691 (34.1%) | 10,799 (39.5%) | 186,490 (34.4%) |
| 1 to <10 | 223,448 (43.4%) | 11,271 (41.3%) | 234,719 (43.3%) |
| 10 to <20 | 89,153 (17.3%) | 4,052 (14.8%) | 93,205 (17.2%) |
| ≥20 | 26,277 (5.1%) | 1,196 (4.4%) | 27,473 (5.1%) |
| **Strenuous physical exercise frequency** | | | |
| Rare/Never | 208,980 (40.6%) | 12,425 (45.5%) | 221,405 (40.9%) |
| < Once | 72,295 (14.0%) | 3,347 (12.3%) | 75,642 (14.0%) |
| Once | 103,998 (20.2%) | 4,918 (18.0%) | 108,916 (20.1%) |
| 2-3 times | 86,373 (16.8%) | 4,167 (15.3%) | 90,540 (16.7%) |
| 4-6 times | 18,268 (3.6%) | 863 (3.2%) | 19,131 (3.5%) |
| Daily | 14,118 (2.7%) | 873 (3.2%) | 14,991 (2.8%) |
| Missing | 10,537 (2.0%) | 725 (2.7%) | 11,262 (2.1%) |
| **Energy intake (kcal/day)** | 1655.15 (416.10) | 1665.25 (429.07) | 1655.65 (416.77) |

| | | | |
|---|---|---|---|
| **Reproductive** | | | |
| **Menopause age** | | | |
| 25-39 | 26,613 (5.2%) | 1,608 (5.9%) | 28,221 (5.2%) |
| 40-44 | 52,397 (10.2%) | 3,129 (11.5%) | 55,526 (10.2%) |
| 45-49 | 123,073 (23.9%) | 7,018 (25.7%) | 130,091 (24.0%) |
| 50-54 | 205,185 (39.9%) | 11,286 (41.3%) | 216,471 (39.9%) |
| 55-60 | 38,895 (7.6%) | 2,388 (8.7%) | 41,283 (7.6%) |
| Missing | 68,406 (13.3%) | 1,889 (6.9%) | 70,295 (13.0%) |
| **Parity** | | | |
| No child | 58,838 (11.4%) | 3,583 (13.1%) | 62,421 (11.5%) |
| One child | 62,362 (12.1%) | 3,463 (12.7%) | 65,825 (12.1%) |
| Two children | 234,922 (45.7%) | 11,419 (41.8%) | 246,341 (45.5%) |
| Three or more children | 157,379 (30.6%) | 8,774 (32.1%) | 166,153 (30.7%) |
| Missing | 1,068 (0.2%) | 79 (0.3%) | 1,147 (0.2%) |
| **MHT use** | | | |
| Never | 234,287 (45.5%) | 13,690 (50.1%) | 247,977 (45.8%) |
| Past | 126,341 (24.6%) | 6,439 (23.6%) | 132,780 (24.5%) |
| Current | 144,311 (28.0%) | 6,478 (23.7%) | 150,789 (27.8%) |
| Missing | 9,630 (1.9%) | 711 (2.6%) | 10,341 (1.9%) |
| **Health-related** | | | |
| **Height** | | | |
| Q1 | 153,615 (29.9%) | 6,848 (25.1%) | 160,463 (29.6%) |
| Q2 | 64,540 (12.5%) | 3,260 (11.9%) | 67,800 (12.5%) |
| Q3 | 90,185 (17.5%) | 4,598 (16.8%) | 94,783 (17.5%) |
| Q4 | 121,844 (23.7%) | 6,925 (25.3%) | 128,769 (23.8%) |
| Q5 | 79,305 (15.4%) | 5,313 (19.4%) | 84,618 (15.6%) |
| Missing | 5,080 (1.0%) | 374 (1.4%) | 5,454 (1.0%) |
| **Family history of hip fractures** | | | |
| No | 466,791 (90.7%) | 23,881 (87.4%) | 490,672 (90.5%) |
| Yes | 38,873 (7.6%) | 2,798 (10.2%) | 41,671 (7.7%) |
| Missing | 8,905 (1.7%) | 639 (2.3%) | 9,544 (1.8%) |
| **BMI (kg/m$^2$)** | | | |
| <18.5 | 4,241 (0.8%) | 493 (1.8%) | 4,734 (0.9%) |
| 18.5-19.9 | 14,718 (2.9%) | 1,201 (4.4%) | 15,919 (2.9%) |
| 20.0-22.4 | 81,273 (15.8%) | 5,257 (19.2%) | 86,530 (16.0%) |
| 22.5-24.9 | 134,533 (26.1%) | 7,444 (27.2%) | 141,977 (26.2%) |
| 25.0-27.4 | 118,554 (23.0%) | 5,910 (21.6%) | 124,464 (23.0%) |
| 27.5-29.9 | 71,766 (13.9%) | 3,218 (11.8%) | 74,984 (13.8%) |
| 30.0-32.4 | 37,637 (7.3%) | 1,608 (5.9%) | 39,245 (7.2%) |
| ≥32.5 | 43,521 (8.5%) | 1,667 (6.1%) | 45,188 (8.3%) |
| Missing | 8,326 (1.6%) | 520 (1.9%) | 8,846 (1.6%) |
| **Self-rated health** | | | |
| Excellent | 100,326 (19.5%) | 4,396 (16.1%) | 104,722 (19.3%) |

| | | | |
|---|---|---|---|
| Good | 305,880 (59.4%) | 15,813 (57.9%) | 321,693 (59.4%) |
| Fair | 79,793 (15.5%) | 5,172 (18.9%) | 84,965 (15.7%) |
| Poor | 6,157 (1.2%) | 482 (1.8%) | 6,639 (1.2%) |
| Missing | 22,413 (4.4%) | 1,455 (5.3%) | 23,868 (4.4%) |

Statistics are mean (SD) or N (%)

**STable 3. Risks of hip fracture with 99 dietary factors with varying levels of adjustments** (Person-years = 10,652,303, 27,318 incident cases)

| Dietary factors | Model 1 | Model 2 | Model 3 | |
|---|---|---|---|---|
| | HR (95% CI) | HR (95% CI) | HR (95% CI) | p-value |
| **Chicken per 20g/day** | 0.76 (0.73, 0.80) | 0.81 (0.78, 0.85) | 0.84 (0.80, 0.87) | 7.45E-17 |
| **Broccoli ≥1 per week** | 0.84 (0.82, 0.87) | 0.90 (0.87, 0.92) | 0.89 (0.87, 0.92) | 1.28E-15 |
| **Vegetables per 100g/day** | 0.83 (0.80, 0.85) | 0.87 (0.84, 0.90) | 0.88 (0.85, 0.91) | 1.48E-15 |
| **Protein per 15g/day** | 0.85 (0.82, 0.88) | 0.74 (0.70, 0.78) | 0.79 (0.75, 0.84) | 1.91E-15 |
| **Chips per 25g/day** | 1.19 (1.15, 1.23) | 1.13 (1.09, 1.17) | 1.14 (1.10, 1.18) | 1.01E-13 |
| **Zinc per 2mg/day** | 0.84 (0.81, 0.87) | 0.77 (0.73, 0.82) | 0.81 (0.77, 0.86) | 4.26E-13 |
| **Garlic ≥1 per week** | 0.87 (0.85, 0.89) | 0.91 (0.88, 0.93) | 0.91 (0.88, 0.93) | 5.33E-13 |
| **Fizzy drinks per 50g/day** | 1.13 (1.07, 1.18) | 1.09 (1.04, 1.14) | 1.18 (1.12, 1.24) | 3.82E-11 |
| **Pasta per 20g/day** | 0.80 (0.77, 0.83) | 0.85 (0.82, 0.89) | 0.87 (0.83, 0.91) | 6.72E-11 |
| **Carotene per 1000 µg/day** | 0.88 (0.85, 0.90) | 0.91 (0.88, 0.94) | 0.91 (0.88, 0.94) | 1.43E-10 |
| **Fiber per 5g/day** | 0.85 (0.82, 0.88) | 0.90 (0.87, 0.93) | 0.89 (0.85, 0.92) | 3.06E-10 |
| **Niacin per 10mg/day** | 0.76 (0.72, 0.81) | 0.76 (0.71, 0.81) | 0.82 (0.77, 0.87) | 3.19E-09 |
| **Fruit (excluding juice) per 200g/day** | 0.77 (0.74, 0.81) | 0.89 (0.85, 0.93) | 0.88 (0.84, 0.92) | 1.61E-08 |
| **Phosphorus per 300mg/day** | 0.89 (0.86, 0.92) | 0.82 (0.78, 0.87) | 0.85 (0.80, 0.90) | 3.16E-08 |
| **Onion ≥1 per week** | 0.87 (0.84, 0.89) | 0.91 (0.88, 0.94) | 0.93 (0.90, 0.95) | 3.58E-07 |
| **Peppers ≥1 per week** | 0.88 (0.86, 0.90) | 0.94 (0.91, 0.96) | 0.94 (0.91, 0.96) | 4.27E-07 |
| **Retinol per 250 µg/day** | 1.15 (1.11, 1.20) | 1.14 (1.09, 1.19) | 1.12 (1.07, 1.17) | 4.98E-07 |
| **Alcohol per 10g/day** | 0.97 (0.96, 0.99) | 0.97 (0.96, 0.99) | 0.96 (0.95, 0.98) | 5.72E-07 |
| **Wholegrains per 20g/day** | 0.79 (0.75, 0.84) | 0.90 (0.85, 0.95) | 0.87 (0.82, 0.92) | 5.86E-07 |
| **Selenium per 10mg/day** | 0.89 (0.86, 0.92) | 0.86 (0.82, 0.90) | 0.89 (0.85, 0.93) | 1.13E-06 |
| **Yoghurt per 50g/day** | 0.86 (0.83, 0.89) | 0.90 (0.87, 0.93) | 0.93 (0.90, 0.95) | 1.17E-06 |
| **Vitamin B6 per 1mg/day** | 0.67 (0.62, 0.73) | 0.69 (0.62, 0.78) | 0.75 (0.67, 0.85) | 1.52E-06 |
| **Rice per 10g/day** | 0.89 (0.87, 0.92) | 0.93 (0.90, 0.95) | 0.94 (0.91, 0.96) | 1.93E-06 |
| **Magnesium per100mg/day** | 0.85 (0.81, 0.90) | 0.84 (0.78, 0.90) | 0.85 (0.80, 0.91) | 3.24E-06 |
| **Apple ≥1 per week** | 0.85 (0.83, 0.88) | 0.93 (0.90, 0.95) | 0.94 (0.91, 0.96) | 4.07E-06 |
| **Banana ≥1 per week** | 0.87 (0.84, 0.90) | 0.92 (0.89, 0.95) | 0.93 (0.90, 0.96) | 8.07E-06 |
| **Courgette ≥1 per week** | 0.90 (0.88, 0.93) | 0.95 (0.92, 0.98) | 0.94 (0.91, 0.97) | 2.70E-05 |
| **Carrot ≥1 per week** | 0.85 (0.82, 0.88) | 0.92 (0.88, 0.95) | 0.92 (0.89, 0.96) | 4.47E-05 |
| **Brussel sprouts ≥1 per week** | 1.05 (1.02, 1.07) | 1.04 (1.02, 1.07) | 1.05 (1.02, 1.08) | 8.88E-05 |
| **Nuts per 10g/day** | 0.89 (0.85, 0.94) | 0.94 (0.90, 0.99) | 0.90 (0.86, 0.95) | 1.18E-04 |
| **Squash drinks per 50g/day** | 1.05 (1.01, 1.08) | 1.05 (1.02, 1.08) | 1.06 (1.03, 1.10) | 1.70E-04 |
| **Crisps per 10g/day** | 1.21 (1.11, 1.31) | 1.15 (1.05, 1.25) | 1.18 (1.08, 1.28) | 1.83E-04 |
| **Free sugars per 30g/day** | 1.09 (1.05, 1.13) | 1.11 (1.05, 1.16) | 1.10 (1.04, 1.15) | 2.69E-04 |
| **Oily fish per 10g/day** | 0.90 (0.87, 0.92) | 0.95 (0.92, 0.98) | 0.94 (0.91, 0.97) | 2.90E-04 |
| **Stewed fruit ≥1 per week** | 1.02 (1.00, 1.05) | 1.05 (1.02, 1.07) | 1.05 (1.02, 1.07) | 3.08E-04 |
| **White bread per 10g/day** | 1.12 (1.08, 1.17) | 1.04 (1.00, 1.09) | 1.08 (1.03, 1.12) | 3.59E-04 |
| **Potassium per 1000mg/day** | 0.88 (0.84, 0.92) | 0.86 (0.81, 0.92) | 0.90 (0.85, 0.96) | 6.02E-04 |
| **SFA per 10g/day** | 1.12 (1.08, 1.16) | 1.13 (1.07, 1.19) | 1.09 (1.04, 1.15) | 7.45E-04 |
| **Soup per 40g/day** | 1.02 (0.98, 1.05) | 1.04 (1.00, 1.08) | 1.06 (1.03, 1.10) | 8.24E-04 |

| | | | | |
|---|---|---|---|---|
| **Thiamin per 1mg/day** | 0.73 (0.66, 0.80) | 0.72 (0.63, 0.83) | 0.79 (0.69, 0.91) | 1.03E-03 |
| **Avocado ≥1 per week** | 0.93 (0.89, 0.97) | 0.95 (0.91, 0.99) | 0.93 (0.89, 0.97) | 1.12E-03 |
| **Lettuce ≥1 per week** | 0.90 (0.88, 0.93) | 0.95 (0.92, 0.97) | 0.95 (0.93, 0.98) | 1.24E-03 |
| **Vitamin C per 100mg/day** | 0.76 (0.72, 0.81) | 0.90 (0.85, 0.95) | 0.91 (0.86, 0.97) | 1.73E-03 |
| **Vitamin $B_{12}$ per 2 µg/day** | 0.89 (0.85, 0.92) | 0.89 (0.85, 0.93) | 0.93 (0.88, 0.97) | 2.00E-03 |
| **Stone fruit ≥1 per week** | 0.91 (0.89, 0.93) | 0.96 (0.93, 0.98) | 0.96 (0.94, 0.99) | 3.05E-03 |
| **Processed meat per 10g/day** | 1.02 (0.98, 1.06) | 1.01 (0.97, 1.05) | 1.06 (1.02, 1.10) | 4.81E-03 |
| **Cauliflower ≥1 per week** | 1.00 (0.98, 1.03) | 1.01 (0.99, 1.04) | 1.04 (1.01, 1.06) | 6.21E-03 |
| **Riboflavin per 1mg/day** | 0.85 (0.79, 0.90) | 0.85 (0.79, 0.92) | 0.89 (0.82, 0.97) | 6.25E-03 |
| **Eggs per 10g/day** | 1.02 (0.99, 1.05) | 1.01 (0.98, 1.04) | 1.04 (1.01, 1.07) | 7.81E-03 |
| **Cucumber ≥1 per week** | 0.91 (0.89, 0.93) | 0.95 (0.93, 0.97) | 0.97 (0.94, 0.99) | 8.02E-03 |
| **Green peas ≥1 per week** | 1.03 (1.00, 1.05) | 1.03 (1.00, 1.06) | 1.04 (1.01, 1.06) | 8.66E-03 |
| **Juice per 100g/day** | 0.99 (0.96, 1.02) | 1.04 (1.00, 1.07) | 1.04 (1.01, 1.08) | 8.80E-03 |
| **Baked beans ≥1 per week** | 1.02 (0.99, 1.04) | 1.02 (1.00, 1.05) | 1.03 (1.01, 1.06) | 9.11E-03 |
| **Mushrooms ≥1 per week** | 0.91 (0.89, 0.93) | 0.95 (0.92, 0.97) | 0.97 (0.94, 0.99) | 1.07E-02 |
| **Calcium per 300mg/day** | 0.95 (0.91, 0.98) | 0.94 (0.89, 0.98) | 0.94 (0.90, 0.99) | 1.31E-02 |
| **Brown bread per 10g/day** | 0.90 (0.86, 0.95) | 0.97 (0.93, 1.02) | 0.94 (0.90, 0.99) | 1.80E-02 |
| **Cereal per 40g/day** | 0.95 (0.92, 0.98) | 0.98 (0.95, 1.02) | 0.96 (0.93, 0.99) | 1.82E-02 |
| **Dried fruit per 10g/day** | 0.92 (0.89, 0.95) | 0.99 (0.96, 1.03) | 0.96 (0.93, 0.99) | 1.98E-02 |
| **Folate per 100 µg/day** | 0.88 (0.84, 0.92) | 0.92 (0.87, 0.97) | 0.94 (0.89, 0.99) | 2.21E-02 |
| **Sweet Biscuits per 10g/day** | 1.08 (1.04, 1.13) | 1.06 (1.02, 1.10) | 1.05 (1.01, 1.09) | 2.50E-02 |
| FDR-CORRECTED NON-SIGNIFICANT BELOW | | | | |
| Vitamin D per 1µg/day | 0.91 (0.88, 0.94) | 0.95 (0.92, 0.99) | 0.97 (0.93, 1.00) | 4.85E-02 |
| Tomato ≥1 per week | 0.90 (0.86, 0.93) | 0.95 (0.91, 0.99) | 0.96 (0.92, 1.00) | 5.49E-02 |
| Jam per 5g/day | 1.10 (1.00, 1.22) | 1.12 (1.01, 1.23) | 1.10 (1.00, 1.22) | 5.69E-02 |
| Sauce per 10g/day | 0.92 (0.87, 0.98) | 0.92 (0.87, 0.98) | 0.95 (0.89, 1.00) | 5.75E-02 |
| Parsnip ≥1 per week | 0.95 (0.92, 0.97) | 0.97 (0.94, 0.99) | 0.98 (0.95, 1.00) | 6.18E-02 |
| Leeks ≥1 per week | 0.94 (0.92, 0.97) | 0.97 (0.95, 1.00) | 0.98 (0.95, 1.00) | 8.18E-02 |
| Soya milk ≥1 per week | 0.95 (0.88, 1.02) | 0.98 (0.91, 1.05) | 0.94 (0.88, 1.01) | 8.98E-02 |
| Potatoes per 40g/day | 1.02 (0.99, 1.06) | 1.04 (1.00, 1.08) | 1.03 (0.99, 1.07) | 9.16E-02 |
| Pear ≥1 per week | 0.93 (0.91, 0.95) | 0.97 (0.94, 0.99) | 0.98 (0.96, 1.00) | 9.69E-02 |
| Tea per 500g/day | 1.05 (1.02, 1.07) | 1.04 (1.01, 1.06) | 1.02 (1.00, 1.05) | 9.79E-02 |
| Celery ≥1 per week | 0.94 (0.91, 0.96) | 0.97 (0.95, 1.00) | 0.98 (0.95, 1.01) | 1.25E-01 |
| Milk per 200g/day | 1.00 (0.96, 1.05) | 0.96 (0.92, 1.01) | 0.97 (0.93, 1.01) | 1.36E-01 |
| Red meat per 20g/day | 0.94 (0.92, 0.97) | 0.94 (0.92, 0.97) | 0.98 (0.95, 1.01) | 1.38E-01 |
| Energy per 1000kj/day | 0.99 (0.97, 1.01) | 0.99 (0.96, 1.01) | 0.98 (0.96, 1.01) | 1.51E-01 |
| Ice cream ≥1 per week | 1.00 (0.97, 1.03) | 1.01 (0.98, 1.04) | 1.02 (0.99, 1.05) | 1.98E-01 |
| Spinach ≥1 per week | 0.98 (0.95, 1.02) | 1.00 (0.96, 1.04) | 0.98 (0.94, 1.02) | 2.50E-01 |
| Coffee per 500g/day | 1.06 (1.02, 1.10) | 0.98 (0.95, 1.02) | 1.02 (0.98, 1.06) | 2.67E-01 |
| Non-oily fish per 10g/day | 0.93 (0.89, 0.97) | 0.95 (0.90, 0.99) | 0.98 (0.93, 1.02) | 2.83E-01 |
| Cheese per 10g/day | 1.03 (1.00, 1.06) | 1.04 (1.01, 1.07) | 1.02 (0.99, 1.05) | 2.95E-01 |
| Red & processed meat per 30g/day | 0.94 (0.91, 0.97) | 0.94 (0.91, 0.97) | 0.98 (0.95, 1.01) | 2.99E-01 |
| Total fat per 20g/day | 1.04 (1.00, 1.07) | 1.05 (0.98, 1.12) | 1.03 (0.97, 1.10) | 3.41E-01 |
| Grapefruit ≥1 per week | 0.97 (0.94, 1.00) | 1.01 (0.98, 1.04) | 1.01 (0.98, 1.04) | 3.63E-01 |

| | | | | |
|---|---|---|---|---|
| PUFA per 5g/day | 0.93 (0.87, 1.00) | 0.91 (0.83, 1.00) | 0.96 (0.88, 1.05) | 3.75E-01 |
| Cake per 10g/day | 1.01 (0.97, 1.05) | 1.01 (0.96, 1.06) | 0.98 (0.94, 1.03) | 3.97E-01 |
| Beetroot ≥1 per week | 0.98 (0.96, 1.01) | 1.00 (0.98, 1.03) | 1.01 (0.99, 1.04) | 3.98E-01 |
| Cabbage ≥1 per week | 0.98 (0.95, 1.00) | 0.97 (0.95, 1.00) | 0.99 (0.97, 1.01) | 4.02E-01 |
| Boiled sweets per 5g/day | 1.07 (1.00, 1.16) | 1.02 (0.94, 1.09) | 1.03 (0.96, 1.11) | 4.25E-01 |
| Chick peas/ lentils ≥1 per week | 0.97 (0.93, 1.01) | 1.00 (0.96, 1.04) | 0.98 (0.94, 1.02) | 4.28E-01 |
| Soya/ tofu ≥1 per week | 1.04 (0.97, 1.11) | 1.06 (0.99, 1.14) | 1.02 (0.96, 1.10) | 4.89E-01 |
| Orange ≥1 per week | 0.94 (0.92, 0.97) | 1.00 (0.98, 1.03) | 1.01 (0.98, 1.03) | 5.05E-01 |
| Vitamin E per 2mg/day | 0.95 (0.91, 0.99) | 0.95 (0.90, 1.00) | 0.98 (0.93, 1.04) | 5.33E-01 |
| MUFA per 5g/day | 1.00 (0.97, 1.02) | 1.00 (0.96, 1.04) | 0.99 (0.95, 1.03) | 5.45E-01 |
| Carbohydrate per 50g/day | 0.99 (0.96, 1.02) | 1.02 (0.97, 1.07) | 1.01 (0.96, 1.06) | 7.27E-01 |
| Sweetcorn ≥1 per week | 0.98 (0.95, 1.00) | 1.00 (0.97, 1.03) | 1.00 (0.98, 1.03) | 7.52E-01 |
| Total sugars per 50g/day | 0.98 (0.94, 1.02) | 1.02 (0.97, 1.08) | 1.01 (0.95, 1.06) | 8.00E-01 |
| Crispbread per 10g/day | 0.97 (0.94, 1.00) | 1.00 (0.96, 1.03) | 1.00 (0.97, 1.04) | 8.38E-01 |
| Swede ≥1 per week | 0.98 (0.95, 1.00) | 0.99 (0.96, 1.01) | 1.00 (0.98, 1.03) | 8.43E-01 |
| Chocolate per 10g/day | 1.03 (0.99, 1.08) | 1.02 (0.97, 1.07) | 1.00 (0.96, 1.05) | 9.90E-01 |
| Green beans ≥1 per week | 0.97 (0.95, 1.00) | 0.99 (0.97, 1.02) | 1.00 (0.98, 1.02) | 9.96E-01 |

The results were ranked on *p*-values from smallest to largest. Abbreviation: SFA saturated fatty acids, MUFA mono-unsaturated fatty acids, PUFA poly-unsaturated fatty acids.

All models were stratified by birth year (≤1921, five-year intervals between 1922 and 1946, >1946), year of completing the baseline dietary questionnaire (per year from 1999 to 2004, and 2005 or later), and 10 regions (9 in England and 1 in Scotland). Model 1 was adjusted for ethnicity (white/ non-white, missing), Townsend deprivation index (quintiles and missing), education (tertiary, secondary, technical, no qualification, missing). Model 2 was additionally adjusted for self-rated health (excellent, good, fair, poor, missing), energy intake (500 to <1,000, 1,000 to <1,250, 1,250 to <1,500, 1,500 to <1,750, 1,750 to <2,000, 2,000 to < 2,250, 2,250 to ≤3,500 kcal/day), smoking (never smoker, past smoker, current smoker <15 cigarettes/day, current smoker ≥15 cigarettes/day, missing), alcohol consumption (<1, 1 to <10, 10 to <20, and ≥20g/day), strenuous exercise frequencies (none, <1, 1, 2-3, 4-6, 7 times/week, missing), MHT use (never, past, current, missing), parity (nulliparous, 1-2 children, ≥3 children, missing), age at menopause (<40, 40-44, 45-49, 50-54, ≥55 years, missing), height (quintiles and missing), family history of hip fractures (yes, no, missing). Model 3 was additionally adjusted for BMI (<18.5, 18.5 to <20, 20 to <22.5, 22.5 to <25, 25 to <27.5, 27.5 to < 30, 30 to <32.5, ≥32.5 kg/m$^2$, missing). For analyses of total energy and alcohol, covariates of energy intake and alcohol consumption were taken out of the model respectively.

**STable 4. Risks of hip fracture with 60 significant dietary factors in subgroups of BMI**

| Dietary factors | Hazard Ratios (95% CI) | | | |
|---|---|---|---|---|
| | **BMI <20 kg/m²** (389,247/1,694)[1] | **BMI 20-24.9 kg/m²** (4,522,153/12,701)[1] | **BMI ≥25 kg/m²** (5,571,541/12,403)[1] | *p*-het[2] |
| **Chicken per 20g/day** | 0.80 (0.68, 0.94) | 0.85 (0.80, 0.90) | 0.84 (0.79, 0.89) | 0.78 |
| **Broccoli ≥1 per week** | 0.88 (0.78, 0.98) | 0.92 (0.88, 0.96) | 0.86 (0.83, 0.90) | 0.08 |
| **Vegetables per 100g/day** | 0.91 (0.80, 1.03) | 0.87 (0.83, 0.91) | 0.88 (0.84, 0.92) | 0.79 |
| **Protein per 15g/day** | 0.72 (0.57, 0.91) | 0.81 (0.75, 0.89) | 0.78 (0.72, 0.85) | 0.60 |
| **Chips per 25g/day** | 1.14 (0.98, 1.31) | 1.14 (1.08, 1.20) | 1.14 (1.08, 1.20) | 1.00 |
| **Zinc per 2mg/day** | 0.75 (0.59, 0.95) | 0.83 (0.76, 0.90) | 0.80 (0.74, 0.87) | 0.67 |
| **Garlic ≥1 per week** | 0.88 (0.79, 0.98) | 0.90 (0.86, 0.93) | 0.92 (0.88, 0.95) | 0.61 |
| **Fizzy drinks per 50g/day** | 1.57 (1.26, 1.94) | 1.16 (1.07, 1.26) | 1.17 (1.10, 1.25) | **0.03** |
| **Pasta per 20g/day** | 0.89 (0.75, 1.06) | 0.86 (0.81, 0.91) | 0.88 (0.83, 0.94) | 0.84 |
| **Carotene per 1000 µg/day** | 0.86 (0.77, 0.97) | 0.91 (0.87, 0.95) | 0.91 (0.87, 0.95) | 0.65 |
| **Fiber per 5g/day** | 0.86 (0.74, 1.00) | 0.87 (0.82, 0.92) | 0.89 (0.84, 0.94) | 0.82 |
| **Niacin per 10mg/day** | 0.80 (0.61, 1.04) | 0.82 (0.74, 0.90) | 0.83 (0.75, 0.91) | 0.96 |
| **Fruit (excluding juice) per 200g/day** | 0.80 (0.67, 0.96) | 0.86 (0.81, 0.92) | 0.90 (0.84, 0.96) | 0.38 |
| **Phosphorus per 300mg/day** | 0.85 (0.67, 1.07) | 0.88 (0.81, 0.96) | 0.82 (0.76, 0.90) | 0.51 |
| **Onion ≥1 per week** | 0.89 (0.80, 1.00) | 0.92 (0.88, 0.96) | 0.94 (0.90, 0.98) | 0.59 |
| **Peppers ≥1 per week** | 0.93 (0.83, 1.03) | 0.93 (0.89, 0.96) | 0.95 (0.91, 0.98) | 0.72 |
| **Retinol per 250 µg/day** | 1.04 (0.86, 1.25) | 1.11 (1.04, 1.19) | 1.15 (1.07, 1.23) | 0.55 |
| **Alcohol per 10g/day** | 0.97 (0.91, 1.03) | 0.95 (0.93, 0.97) | 0.97 (0.95, 0.99) | 0.36 |
| **Wholegrains per 20g/day** | 0.85 (0.68, 1.05) | 0.86 (0.79, 0.93) | 0.87 (0.81, 0.95) | 0.97 |
| **Selenium per 10mg/day** | 0.78 (0.65, 0.94) | 0.90 (0.84, 0.96) | 0.89 (0.83, 0.95) | 0.36 |
| **Yoghurt per 50g/day** | 0.94 (0.82, 1.07) | 0.95 (0.91, 1.00) | 0.89 (0.85, 0.93) | 0.14 |
| **Vitamin B6 per 1mg/day** | 1.10 (0.68, 1.76) | 0.82 (0.69, 0.97) | 0.65 (0.54, 0.77) | **0.05** |
| **Rice per 10g/day** | 0.90 (0.81, 1.01) | 0.92 (0.88, 0.96) | 0.96 (0.92, 1.00) | 0.29 |
| **Magnesium per100mg/day** | 0.79 (0.60, 1.03) | 0.86 (0.78, 0.95) | 0.85 (0.77, 0.94) | 0.85 |
| **Apple ≥1 per week** | 0.93 (0.83, 1.04) | 0.93 (0.89, 0.97) | 0.94 (0.90, 0.98) | 0.94 |
| **Banana ≥1 per week** | 0.92 (0.82, 1.03) | 0.95 (0.91, 1.00) | 0.91 (0.87, 0.95) | 0.42 |
| **Courgette ≥1 per week** | 0.90 (0.80, 1.01) | 0.94 (0.90, 0.98) | 0.94 (0.90, 0.99) | 0.78 |
| **Carrot ≥1 per week** | 0.95 (0.81, 1.10) | 0.92 (0.87, 0.98) | 0.90 (0.85, 0.96) | 0.76 |
| **Brussel sprouts ≥1 per week** | 1.10 (0.99, 1.21) | 1.03 (1.00, 1.07) | 1.06 (1.02, 1.10) | 0.33 |
| **Nuts per 10g/day** | 0.87 (0.71, 1.06) | 0.87 (0.81, 0.94) | 0.95 (0.87, 1.03) | 0.29 |
| **Squash drinks per 50g/day** | 1.18 (1.03, 1.35) | 1.04 (0.99, 1.09) | 1.07 (1.03, 1.12) | 0.20 |
| **Crisps per 10g/day** | 1.04 (0.72, 1.48) | 1.27 (1.12, 1.44) | 1.14 (1.01, 1.29) | 0.36 |
| **Free sugars per 30g/day** | 1.25 (1.02, 1.52) | 1.10 (1.02, 1.18) | 1.09 (1.01, 1.17) | 0.45 |
| **Oily fish per 10g/day** | 0.86 (0.76, 0.98) | 0.97 (0.92, 1.01) | 0.93 (0.89, 0.98) | 0.15 |
| **Stewed fruit ≥1 per week** | 1.10 (0.99, 1.22) | 1.02 (0.98, 1.06) | 1.08 (1.04, 1.12) | 0.08 |
| **White bread per 10g/day** | 1.03 (0.87, 1.22) | 1.07 (1.00, 1.13) | 1.09 (1.02, 1.15) | 0.79 |
| **Potassium per 1000mg/day** | 0.87 (0.68, 1.11) | 0.92 (0.84, 1.01) | 0.87 (0.80, 0.95) | 0.67 |
| **SFA per 10g/day** | 1.04 (0.84, 1.28) | 1.08 (1.00, 1.16) | 1.14 (1.06, 1.23) | 0.51 |
| **Soup per 40g/day** | 1.03 (0.89, 1.20) | 1.08 (1.02, 1.13) | 1.05 (0.99, 1.10) | 0.69 |
| **Thiamin per 1mg/day** | 1.17 (0.66, 2.08) | 0.77 (0.63, 0.94) | 0.75 (0.61, 0.93) | 0.36 |
| **Avocado ≥1 per week** | 0.92 (0.79, 1.08) | 0.92 (0.87, 0.98) | 0.96 (0.89, 1.03) | 0.66 |
| **Lettuce ≥1 per week** | 0.95 (0.85, 1.07) | 0.94 (0.90, 0.98) | 0.97 (0.93, 1.01) | 0.58 |
| **Vitamin C per 100mg/day** | 0.89 (0.71, 1.13) | 0.90 (0.83, 0.98) | 0.92 (0.84, 1.00) | 0.92 |
| **Vitamin $B_{12}$ per 2 µg/day** | 0.92 (0.77, 1.12) | 0.96 (0.89, 1.03) | 0.91 (0.84, 0.97) | 0.59 |

| | | | | |
|---|---|---|---|---|
| **Stone fruit ≥1 per week** | 0.98 (0.88, 1.08) | 0.95 (0.91, 0.98) | 0.97 (0.94, 1.01) | 0.68 |
| **Processed meat per 10g/day** | 1.03 (0.88, 1.22) | 1.04 (0.98, 1.10) | 1.07 (1.01, 1.14) | 0.77 |
| **Cauliflower ≥1 per week** | 1.09 (0.98, 1.20) | 1.01 (0.98, 1.05) | 1.04 (1.00, 1.08) | 0.27 |
| **Riboflavin per 1mg/day** | 0.96 (0.69, 1.32) | 0.97 (0.86, 1.09) | 0.81 (0.72, 0.91) | 0.09 |
| **Eggs per 10g/day** | 1.04 (0.93, 1.16) | 1.05 (1.00, 1.09) | 1.03 (0.99, 1.07) | 0.81 |
| **Cucumber ≥1 per week** | 1.04 (0.94, 1.15) | 0.97 (0.93, 1.00) | 0.95 (0.92, 0.99) | 0.24 |
| **Green peas ≥1 per week** | 0.98 (0.88, 1.09) | 1.04 (1.00, 1.08) | 1.03 (0.99, 1.07) | 0.59 |
| **Juice per 100g/day** | 1.08 (0.94, 1.23) | 1.04 (0.99, 1.09) | 1.06 (1.01, 1.11) | 0.79 |
| **Baked beans ≥1 per week** | 1.25 (1.13, 1.39) | 1.03 (0.99, 1.07) | 1.01 (0.97, 1.04) | **<0.001** |
| **Mushrooms ≥1 per week** | 1.04 (0.93, 1.15) | 0.95 (0.92, 0.99) | 0.97 (0.93, 1.01) | 0.26 |
| **Calcium per 300mg/day** | 0.92 (0.76, 1.11) | 0.97 (0.90, 1.04) | 0.92 (0.86, 0.99) | 0.57 |
| **Brown bread per 10g/day** | 0.85 (0.70, 1.03) | 0.95 (0.89, 1.02) | 0.94 (0.87, 1.01) | 0.58 |
| **Cereal per 40g/day** | 1.08 (0.94, 1.23) | 0.96 (0.92, 1.01) | 0.95 (0.90, 0.99) | 0.21 |
| **Dried fruit per 10g/day** | 1.02 (0.89, 1.17) | 0.96 (0.92, 1.01) | 0.94 (0.90, 1.00) | 0.53 |
| **Folate per 100 µg/day** | 1.01 (0.81, 1.27) | 0.95 (0.87, 1.03) | 0.90 (0.83, 0.98) | 0.51 |
| **Sweet Biscuits per 10g/day** | 0.98 (0.83, 1.16) | 1.09 (1.02, 1.16) | 1.03 (0.97, 1.10) | 0.32 |

[1] Person-years/ number of incident cases

[2] *p*-heterogeneity among all subgroups and was calculated with the inverse variance-weighted average of stratum-specific log hazard ratios.

Abbreviation: SFA saturated fatty acids.

All models were stratified by birth year (≤1921,five-year intervals between 1922 and 1946, >1946), year of completing the baseline dietary questionnaire (per year from 1999 to 2004, and 2005 or later), and 10 regions and adjusted for ethnicity (white/ non-white, missing), Townsend deprivation index (quintiles and missing), education (tertiary, secondary, technical, no qualification, missing), self-rated health (excellent, good, fair, poor, missing), energy intake (500 to <1,000, 1,000 to <1,250, 1,250 to <1,500, 1,500 to <1,750, 1,750 to <2,000, 2,000 to < 2,250, 2,250 to ≤3,500 kcal/day), smoking (never smoker, past smoker, current smoker <15 cigarettes/day, current smoker ≥15 cigarettes/day, missing), alcohol consumption (<1, 1 to <10, 10 to <20, and ≥20g/day), strenuous exercise frequencies (none, <1, 1, 2-3, 4-6, 7 times/week, missing), MHT use (never, past, current, missing), parity (nulliparous, 1-2 children, ≥3 children, missing), age at menopause (<40, 40-44, 45-49, 50-54, ≥55 years, missing), height (quintiles and missing), family history of hip fractures (yes, no, missing), and BMI (<18.5, 18.5 to <20, 20 to <22.5, 22.5 to <25, 25 to <27.5, 27.5 to < 30, 30 to <32.5, ≥32.5 kg/m$^2$, missing). For analyses of total energy and alcohol, covariates of energy intake and alcohol consumption were taken out of the model respectively.

**STable 5. Risks of hip fracture with 60 significant dietary factors in subgroups of smoking status**

| Dietary factors | Hazard Ratios (95% CI) Never Smoker (6,007,960/14,981)[1] | Ever Smoker (4,641,460/12,330)[1] | *p*-het[2] |
|---|---|---|---|
| **Chicken per 20g/day** | 0.81 (0.76, 0.86) | 0.86 (0.81, 0.92) | 0.19 |
| **Broccoli ≥1 per week** | 0.88 (0.85, 0.91) | 0.91 (0.87, 0.95) | 0.24 |
| **Vegetables per 100g/day** | 0.84 (0.81, 0.88) | 0.93 (0.89, 0.97) | **<0.001** |
| **Protein per 15g/day** | 0.73 (0.68, 0.79) | 0.86 (0.79, 0.94) | **0.01** |
| **Chips per 25g/day** | 1.13 (1.08, 1.19) | 1.15 (1.09, 1.21) | 0.63 |
| **Zinc per 2mg/day** | 0.80 (0.74, 0.87) | 0.82 (0.75, 0.89) | 0.68 |
| **Garlic ≥1 per week** | 0.90 (0.86, 0.93) | 0.93 (0.89, 0.96) | 0.24 |
| **Fizzy drinks per 50g/day** | 1.16 (1.08, 1.24) | 1.20 (1.12, 1.28) | 0.49 |
| **Pasta per 20g/day** | 0.86 (0.81, 0.92) | 0.87 (0.82, 0.93) | 0.80 |
| **Carotene per 1000 µg/day** | 0.89 (0.85, 0.92) | 0.94 (0.90, 0.98) | 0.07 |
| **Fiber per 5g/day** | 0.89 (0.84, 0.93) | 0.88 (0.83, 0.93) | 0.77 |
| **Niacin per 10mg/day** | 0.74 (0.67, 0.81) | 0.92 (0.84, 1.02) | **0.002** |
| **Fruit (excluding juice) per 200g/day** | 0.88 (0.83, 0.94) | 0.88 (0.82, 0.94) | 1.00 |
| **Phosphorus per 300mg/day** | 0.84 (0.77, 0.90) | 0.87 (0.80, 0.94) | 0.54 |
| **Onion ≥1 per week** | 0.90 (0.87, 0.94) | 0.96 (0.92, 1.01) | 0.04 |
| **Peppers ≥1 per week** | 0.93 (0.90, 0.96) | 0.95 (0.92, 0.99) | 0.39 |
| **Retinol per 250 µg/day** | 1.14 (1.08, 1.22) | 1.10 (1.03, 1.17) | 0.43 |
| **Alcohol per 10g/day** | 0.94 (0.92, 0.96) | 0.98 (0.96, 1.00) | **0.01** |
| **Wholegrains per 20g/day** | 0.89 (0.82, 0.95) | 0.85 (0.78, 0.92) | 0.42 |
| **Selenium per 10mg/day** | 0.85 (0.80, 0.91) | 0.94 (0.88, 1.00) | **0.03** |
| **Yoghurt per 50g/day** | 0.95 (0.91, 0.99) | 0.90 (0.86, 0.94) | 0.08 |
| **Vitamin B6 per 1mg/day** | 0.70 (0.60, 0.82) | 0.83 (0.70, 0.98) | 0.15 |
| **Rice per 10g/day** | 0.93 (0.90, 0.97) | 0.94 (0.90, 0.98) | 0.71 |
| **Magnesium per100mg/day** | 0.81 (0.74, 0.89) | 0.90 (0.82, 1.00) | 0.13 |
| **Apple ≥1 per week** | 0.94 (0.90, 0.98) | 0.94 (0.90, 0.98) | 1.00 |
| **Banana ≥1 per week** | 0.94 (0.90, 0.98) | 0.92 (0.88, 0.97) | 0.51 |
| **Courgette ≥1 per week** | 0.92 (0.88, 0.96) | 0.97 (0.92, 1.01) | 0.10 |
| **Carrot ≥1 per week** | 0.94 (0.88, 0.99) | 0.91 (0.86, 0.96) | 0.43 |
| **Brussel sprouts ≥1 per week** | 1.05 (1.01, 1.08) | 1.06 (1.02, 1.10) | 0.71 |
| **Nuts per 10g/day** | 0.90 (0.84, 0.97) | 0.90 (0.84, 0.98) | 1.00 |
| **Squash drinks per 50g/day** | 1.06 (1.02, 1.11) | 1.06 (1.01, 1.11) | 1.00 |
| **Crisps per 10g/day** | 1.30 (1.16, 1.47) | 1.07 (0.95, 1.22) | **0.03** |
| **Free sugars per 30g/day** | 1.13 (1.06, 1.21) | 1.05 (0.97, 1.13) | 0.15 |
| **Oily fish per 10g/day** | 0.92 (0.88, 0.96) | 0.98 (0.93, 1.03) | 0.06 |
| **Stewed fruit ≥1 per week** | 1.05 (1.01, 1.08) | 1.05 (1.01, 1.09) | 1.00 |
| **White bread per 10g/day** | 1.10 (1.04, 1.16) | 1.05 (0.99, 1.12) | 0.27 |
| **Potassium per 1000mg/day** | 0.86 (0.79, 0.93) | 0.95 (0.87, 1.04) | 0.11 |
| **SFA per 10g/day** | 1.17 (1.09, 1.25) | 1.01 (0.94, 1.09) | **0.004** |
| **Soup per 40g/day** | 1.06 (1.01, 1.12) | 1.06 (1.01, 1.12) | 1.00 |
| **Thiamin per 1mg/day** | 0.78 (0.64, 0.94) | 0.81 (0.66, 0.99) | 0.79 |
| **Avocado ≥1 per week** | 0.92 (0.86, 0.97) | 0.95 (0.89, 1.01) | 0.47 |

| | | | |
|---|---|---|---|
| **Lettuce ≥1 per week** | 0.93 (0.89, 0.97) | 0.99 (0.95, 1.03) | **0.04** |
| **Vitamin C per 100mg/day** | 0.91 (0.84, 0.99) | 0.91 (0.83, 0.99) | 1.00 |
| **Vitamin $B_{12}$ per 2 µg/day** | 0.89 (0.83, 0.95) | 0.97 (0.90, 1.04) | 0.09 |
| **Stone fruit ≥1 per week** | 0.97 (0.93, 1.00) | 0.96 (0.92, 0.99) | 0.69 |
| **Processed meat per 10g/day** | 1.06 (1.00, 1.12) | 1.05 (0.99, 1.12) | 0.82 |
| **Cauliflower ≥1 per week** | 1.04 (1.01, 1.08) | 1.02 (0.99, 1.06) | 0.43 |
| **Riboflavin per 1mg/day** | 0.87 (0.78, 0.98) | 0.92 (0.82, 1.03) | 0.50 |
| **Eggs per 10g/day** | 1.01 (0.97, 1.05) | 1.07 (1.03, 1.12) | **0.05** |
| **Cucumber ≥1 per week** | 0.97 (0.93, 1.00) | 0.97 (0.94, 1.01) | 1.00 |
| **Green peas ≥1 per week** | 1.03 (0.99, 1.07) | 1.05 (1.00, 1.09) | 0.52 |
| **Juice per 100g/day** | 1.06 (1.02, 1.11) | 1.02 (0.97, 1.07) | 0.24 |
| **Baked beans ≥1 per week** | 1.03 (1.00, 1.07) | 1.03 (1.00, 1.07) | 1.00 |
| **Mushrooms ≥1 per week** | 0.95 (0.92, 0.98) | 0.99 (0.96, 1.03) | 0.09 |
| **Calcium per 300mg/day** | 0.96 (0.90, 1.02) | 0.92 (0.86, 0.99) | 0.38 |
| **Brown bread per 10g/day** | 0.94 (0.88, 1.00) | 0.94 (0.88, 1.01) | 1.00 |
| **Cereal per 40g/day** | 0.97 (0.93, 1.01) | 0.96 (0.91, 1.00) | 0.75 |
| **Dried fruit per 10g/day** | 0.97 (0.93, 1.02) | 0.94 (0.89, 0.99) | 0.38 |
| **Folate per 100 µg/day** | 0.92 (0.85, 0.99) | 0.96 (0.88, 1.04) | 0.46 |
| **Sweet Biscuits per 10g/day** | 1.10 (1.04, 1.16) | 1.00 (0.94, 1.06) | **0.02** |

[1] Person-years/ number of incident cases

[2] *p*-heterogeneity between subgroups and was calculated with the inverse variance-weighted average of stratum-specific log hazard ratios.

Abbreviation: SFA saturated fatty acids.

All models were stratified by birth year (≤1921,five-year intervals between 1922 and 1946, >1946), year of completing the baseline dietary questionnaire (per year from 1999 to 2004, and 2005 or later), and 10 regions (9 in England and 1 in Scotland) and adjusted for ethnicity (white/ non-white, missing), Townsend deprivation index (quintiles and missing), education (tertiary, secondary, technical, no qualification, missing), self-rated health (excellent, good, fair, poor, missing), energy intake (500 to <1,000, 1,000 to <1,250, 1,250 to <1,500, 1,500 to <1,750, 1,750 to <2,000, 2,000 to < 2,250, 2,250 to ≤3,500 kcal/day), smoking (never smoker, past smoker, current smoker <15 cigarettes/day, current smoker ≥15 cigarettes/day, missing), alcohol consumption (<1, 1 to <10, 10 to <20, and ≥20g/day), strenuous exercise frequencies (none, <1, 1, 2-3, 4-6, 7 times/week, missing), MHT use (never, past, current, missing), parity (nulliparous, 1-2 children, ≥3 children, missing), age at menopause (<40, 40-44, 45-49, 50-54, ≥55 years, missing), height (quintiles and missing), family history of hip fractures (yes, no, missing), and BMI (<18.5, 18.5 to <20, 20 to <22.5, 22.5 to <25, 25 to <27.5, 27.5 to < 30, 30 to <32.5, ≥32.5 kg/m$^2$, missing). For analyses of total energy and alcohol, covariates of energy intake and alcohol consumption were taken out of the model respectively.

**STable 6. Risks of hip fracture with 60 significant dietary factors in subgroups of alcohol consumers**

| Dietary factors | Hazard Ratios (95% CI) **1 to <10g/day** (4,665,740/11,271)[1] | **≥10g/d** (2,379,049/5,248)[1] | *p*-het[2] |
|---|---|---|---|
| **Chicken per 20g/day** | 0.82 (0.77, 0.88) | 0.88 (0.80, 0.98) | 0.25 |
| **Broccoli ≥1 per week** | 0.89 (0.85, 0.93) | 0.90 (0.84, 0.96) | 0.79 |
| **Vegetables per 100g/day** | 0.86 (0.82, 0.91) | 0.89 (0.83, 0.96) | 0.45 |
| **Protein per 15g/day** | 0.80 (0.73, 0.87) | 0.74 (0.64, 0.84) | 0.34 |
| **Chips per 25g/day** | 1.19 (1.12, 1.25) | 1.11 (1.02, 1.20) | 0.16 |
| **Zinc per 2mg/day** | 0.82 (0.75, 0.90) | 0.73 (0.64, 0.83) | 0.15 |
| **Garlic ≥1 per week** | 0.92 (0.88, 0.95) | 0.89 (0.84, 0.94) | 0.34 |
| **Fizzy drinks per 50g/day** | 1.19 (1.10, 1.29) | 1.24 (1.11, 1.39) | 0.56 |
| **Pasta per 20g/day** | 0.87 (0.81, 0.93) | 0.83 (0.75, 0.91) | 0.44 |
| **Carotene per 1000 µg/day** | 0.92 (0.88, 0.96) | 0.93 (0.87, 0.99) | 0.79 |
| **Fiber per 5g/day** | 0.83 (0.79, 0.89) | 0.84 (0.77, 0.91) | 0.82 |
| **Niacin per 10mg/day** | 0.76 (0.69, 0.85) | 0.79 (0.68, 0.93) | 0.69 |
| **Fruit (excluding juice) per 200g/day** | 0.81 (0.76, 0.87) | 0.89 (0.81, 0.99) | 0.13 |
| **Phosphorus per 300mg/day** | 0.87 (0.80, 0.96) | 0.75 (0.66, 0.86) | 0.07 |
| **Onion ≥1 per week** | 0.91 (0.87, 0.95) | 0.91 (0.84, 0.98) | 1.00 |
| **Peppers ≥1 per week** | 0.95 (0.91, 0.98) | 0.93 (0.88, 0.98) | 0.52 |
| **Retinol per 250 µg/day** | 1.19 (1.11, 1.28) | 1.16 (1.05, 1.28) | 0.68 |
| **Alcohol per 10g/day** | 1.00 (0.95, 1.05) | 1.10 (1.05, 1.15) | **0.01** |
| **Wholegrains per 20g/day** | 0.83 (0.76, 0.90) | 0.75 (0.66, 0.85) | 0.19 |
| **Selenium per 10mg/day** | 0.88 (0.82, 0.94) | 0.95 (0.86, 1.05) | 0.21 |
| **Yoghurt per 50g/day** | 0.94 (0.89, 0.98) | 0.89 (0.82, 0.96) | 0.25 |
| **Vitamin B6 per 1mg/day** | 0.65 (0.54, 0.78) | 0.79 (0.60, 1.03) | 0.24 |
| **Rice per 10g/day** | 0.95 (0.91, 0.99) | 0.95 (0.90, 1.01) | 1.00 |
| **Magnesium per100mg/day** | 0.81 (0.73, 0.90) | 0.73 (0.63, 0.86) | 0.28 |
| **Apple ≥1 per week** | 0.91 (0.87, 0.95) | 0.90 (0.85, 0.96) | 0.77 |
| **Banana ≥1 per week** | 0.92 (0.88, 0.97) | 0.94 (0.88, 1.01) | 0.62 |
| **Courgette ≥1 per week** | 0.92 (0.88, 0.96) | 0.95 (0.90, 1.01) | 0.38 |
| **Carrot ≥1 per week** | 0.92 (0.86, 0.98) | 0.91 (0.84, 0.99) | 0.84 |
| **Brussel sprouts ≥1 per week** | 1.02 (0.98, 1.06) | 1.07 (1.01, 1.13) | 0.17 |
| **Nuts per 10g/day** | 0.83 (0.77, 0.90) | 0.86 (0.77, 0.96) | 0.61 |
| **Squash drinks per 50g/day** | 1.04 (0.99, 1.10) | 1.15 (1.06, 1.24) | **0.04** |
| **Crisps per 10g/day** | 1.22 (1.06, 1.39) | 1.01 (0.83, 1.23) | 0.12 |
| **Free sugars per 30g/day** | 1.13 (1.05, 1.22) | 1.01 (0.90, 1.14) | 0.12 |
| **Oily fish per 10g/day** | 0.92 (0.88, 0.97) | 1.02 (0.95, 1.10) | **0.02** |
| **Stewed fruit ≥1 per week** | 1.04 (1.00, 1.08) | 1.02 (0.96, 1.08) | 0.59 |
| **White bread per 10g/day** | 1.13 (1.06, 1.21) | 1.13 (1.03, 1.25) | 1.00 |
| **Potassium per 1000mg/day** | 0.86 (0.78, 0.95) | 0.87 (0.76, 1.01) | 0.90 |
| **SFA per 10g/day** | 1.18 (1.09, 1.28) | 1.08 (0.96, 1.21) | 0.22 |
| **Soup per 40g/day** | 1.10 (1.04, 1.16) | 1.02 (0.94, 1.10) | 0.12 |
| **Thiamin per 1mg/day** | 0.78 (0.63, 0.97) | 0.73 (0.53, 1.00) | 0.74 |
| **Avocado ≥1 per week** | 0.91 (0.85, 0.97) | 0.96 (0.89, 1.04) | 0.30 |

| | | | |
|---|---|---|---|
| **Lettuce ≥1 per week** | 0.94 (0.90, 0.98) | 0.95 (0.88, 1.01) | 0.80 |
| **Vitamin C per 100mg/day** | 0.86 (0.79, 0.95) | 0.97 (0.85, 1.11) | 0.15 |
| **Vitamin $B_{12}$ per 2 µg/day** | 0.91 (0.85, 0.98) | 0.95 (0.85, 1.06) | 0.52 |
| **Stone fruit ≥1 per week** | 0.95 (0.91, 0.98) | 0.99 (0.94, 1.05) | 0.22 |
| **Processed meat per 10g/day** | 1.08 (1.02, 1.15) | 1.13 (1.03, 1.23) | 0.41 |
| **Cauliflower ≥1 per week** | 1.04 (1.00, 1.09) | 1.04 (0.98, 1.10) | 1.00 |
| **Riboflavin per 1mg/day** | 0.92 (0.81, 1.05) | 0.75 (0.62, 0.90) | 0.08 |
| **Eggs per 10g/day** | 1.04 (0.99, 1.08) | 1.08 (1.02, 1.15) | 0.32 |
| **Cucumber ≥1 per week** | 0.95 (0.91, 0.98) | 1.01 (0.95, 1.07) | 0.09 |
| **Green peas ≥1 per week** | 1.07 (1.02, 1.11) | 1.05 (0.99, 1.12) | 0.62 |
| **Juice per 100g/day** | 1.05 (1.00, 1.10) | 1.05 (0.97, 1.13) | 1.00 |
| **Baked beans ≥1 per week** | 1.05 (1.01, 1.09) | 1.03 (0.98, 1.09) | 0.56 |
| **Mushrooms ≥1 per week** | 0.96 (0.92, 0.99) | 0.95 (0.89, 1.01) | 0.78 |
| **Calcium per 300mg/day** | 0.98 (0.91, 1.05) | 0.89 (0.80, 0.99) | 0.14 |
| **Brown bread per 10g/day** | 0.91 (0.84, 0.98) | 0.89 (0.79, 1.00) | 0.76 |
| **Cereal per 40g/day** | 0.94 (0.89, 0.99) | 0.91 (0.84, 0.98) | 0.50 |
| **Dried fruit per 10g/day** | 0.95 (0.91, 1.01) | 0.89 (0.82, 0.97) | 0.20 |
| **Folate per 100 µg/day** | 0.89 (0.82, 0.97) | 0.92 (0.81, 1.05) | 0.67 |
| **Sweet Biscuits per 10g/day** | 1.05 (0.98, 1.12) | 1.08 (0.97, 1.19) | 0.65 |

[1] Person-years/ number of incident cases

[2] *p*-heterogeneity between subgroups and was calculated with the inverse variance-weighted average of stratum-specific log hazard ratios.

Abbreviation: SFA saturated fatty acids.

All models were stratified by birth year (≤1921,five-year intervals between 1922 and 1946, >1946), year of completing the baseline dietary questionnaire (per year from 1999 to 2004, and 2005 or later), and 10 regions (9 in England and 1 in Scotland) and adjusted for ethnicity (white/ non-white, missing), Townsend deprivation index (quintiles and missing), education (tertiary, secondary, technical, no qualification, missing), self-rated health (excellent, good, fair, poor, missing), energy intake (500 to <1,000, 1,000 to <1,250, 1,250 to <1,500, 1,500 to <1,750, 1,750 to <2,000, 2,000 to < 2,250, 2,250 to ≤3,500 kcal/day), smoking (never smoker, past smoker, current smoker <15 cigarettes/day, current smoker ≥15 cigarettes/day, missing), alcohol consumption (<1, 1 to <10, 10 to <20, and ≥20g/day), strenuous exercise frequencies (none, <1, 1, 2-3, 4-6, 7 times/week, missing), MHT use (never, past, current, missing), parity (nulliparous, 1-2 children, ≥3 children, missing), age at menopause (<40, 40-44, 45-49, 50-54, ≥55 years, missing), height (quintiles and missing), family history of hip fractures (yes, no, missing), and BMI (<18.5, 18.5 to <20, 20 to <22.5, 22.5 to <25, 25 to <27.5, 27.5 to < 30, 30 to <32.5, ≥32.5 kg/$m^2$, missing). For analyses of total energy and alcohol, covariates of energy intake and alcohol consumption were taken out of the model respectively.

**STable 7. Risks of hip fracture with 60 significant dietary factors in subgroups of baseline health status**

| Dietary factors | Hazard Ratios (95% CI) | | |
|---|---|---|---|
| | **excellent/good health without comorbidities** (6,350,288/13,994)[1] | **fair/poor health or with comorbidities** (3,950,744/12,372)[1] | *p*-het[2] |
| **Chicken per 20g/day** | 0.85 (0.80, 0.90) | 0.88 (0.84, 0.92) | 1.00 |
| **Broccoli ≥1 per week** | 0.92 (0.89, 0.96) | 0.83 (0.78, 0.88) | 0.58 |
| **Vegetables per 100g/day** | 0.88 (0.85, 0.92) | 0.97 (0.94, 0.99) | 0.54 |
| **Protein per 15g/day** | 0.80 (0.73, 0.86) | 1.13 (1.07, 1.19) | 0.49 |
| **Chips per 25g/day** | 1.16 (1.10, 1.22) | 0.91 (0.85, 0.97) | 0.31 |
| **Zinc per 2mg/day** | 0.81 (0.74, 0.87) | 1.19 (1.11, 1.27) | 0.50 |
| **Garlic ≥1 per week** | 0.91 (0.88, 0.94) | 0.90 (0.84, 0.96) | 0.08 |
| **Fizzy drinks per 50g/day** | 1.15 (1.07, 1.24) | 0.95 (0.92, 0.99) | 0.11 |
| **Pasta per 20g/day** | 0.83 (0.78, 0.88) | 0.91 (0.84, 0.99) | 0.11 |
| **Carotene per 1000 µg/day** | 0.93 (0.89, 0.97) | 1.06 (1.00, 1.12) | 0.25 |
| **Fiber per 5g/day** | 0.88 (0.84, 0.93) | 0.95 (0.88, 1.03) | 0.06 |
| **Niacin per 10mg/day** | 0.80 (0.73, 0.88) | 0.94 (0.90, 0.99) | 0.33 |
| **Fruit (excluding juice) per 200g/day** | 0.87 (0.82, 0.92) | 1.10 (1.05, 1.15) | **0.02** |
| **Phosphorus per 300mg/day** | 0.84 (0.78, 0.91) | 0.97 (0.92, 1.02) | 0.56 |
| **Onion ≥1 per week** | 0.93 (0.90, 0.97) | 0.94 (0.89, 0.98) | 0.79 |
| **Peppers ≥1 per week** | 0.93 (0.90, 0.97) | 1.03 (0.97, 1.10) | 0.18 |
| **Retinol per 250 µg/day** | 1.16 (1.09, 1.24) | 0.99 (0.92, 1.06) | 0.05 |
| **Alcohol per 10g/day** | 0.96 (0.94, 0.98) | 1.09 (1.02, 1.16) | 0.09 |
| **Wholegrains per 20g/day** | 0.83 (0.77, 0.90) | 1.18 (1.03, 1.35) | 1.00 |
| **Selenium per 10mg/day** | 0.89 (0.83, 0.94) | 1.05 (1.00, 1.11) | 0.43 |
| **Yoghurt per 50g/day** | 0.91 (0.87, 0.95) | 0.97 (0.93, 1.02) | 0.35 |
| **Vitamin B6 per 1mg/day** | 0.77 (0.65, 0.90) | 1.00 (0.96, 1.04) | **0.02** |
| **Rice per 10g/day** | 0.91 (0.88, 0.95) | 1.06 (1.01, 1.11) | 0.38 |
| **Magnesium per100mg/day** | 0.81 (0.74, 0.89) | 0.78 (0.71, 0.85) | 0.68 |
| **Apple ≥1 per week** | 0.93 (0.89, 0.97) | 0.82 (0.74, 0.90) | 0.72 |
| **Banana ≥1 per week** | 0.92 (0.88, 0.97) | 0.80 (0.74, 0.87) | 0.83 |
| **Courgette ≥1 per week** | 0.93 (0.90, 0.97) | 0.89 (0.84, 0.94) | 0.77 |
| **Carrot ≥1 per week** | 0.90 (0.85, 0.95) | 0.89 (0.85, 0.93) | 0.17 |
| **Brussel sprouts ≥1 per week** | 1.04 (1.00, 1.07) | 0.89 (0.81, 0.99) | 0.18 |
| **Nuts per 10g/day** | 0.86 (0.80, 0.92) | 0.85 (0.78, 0.93) | 0.84 |
| **Squash drinks per 50g/day** | 1.02 (0.97, 1.06) | 0.70 (0.59, 0.83) | 0.43 |
| **Crisps per 10g/day** | 1.18 (1.05, 1.33) | 0.91 (0.83, 1.00) | 0.60 |
| **Free sugars per 30g/day** | 1.08 (1.00, 1.15) | 0.92 (0.84, 1.00) | 0.72 |
| **Oily fish per 10g/day** | 0.93 (0.89, 0.97) | 0.90 (0.84, 0.96) | 0.81 |
| **Stewed fruit ≥1 per week** | 1.05 (1.02, 1.09) | 1.10 (1.02, 1.18) | 0.72 |
| **White bread per 10g/day** | 1.11 (1.05, 1.17) | 1.09 (1.02, 1.17) | 0.20 |
| **Potassium per 1000mg/day** | 0.88 (0.80, 0.95) | 0.74 (0.60, 0.91) | 0.53 |
| **SFA per 10g/day** | 1.15 (1.07, 1.23) | 0.95 (0.87, 1.03) | 0.59 |
| **Soup per 40g/day** | 1.08 (1.03, 1.13) | 0.92 (0.85, 0.98) | 1.00 |
| **Thiamin per 1mg/day** | 0.81 (0.67, 0.99) | 0.91 (0.81, 1.03) | 0.59 |
| **Avocado ≥1 per week** | 0.93 (0.88, 0.99) | 1.05 (0.98, 1.14) | 0.08 |

| | | | |
|---|---|---|---|
| **Lettuce ≥1 per week** | 0.95 (0.92, 0.99) | 0.96 (0.90, 1.03) | 0.51 |
| **Vitamin C per 100mg/day** | 0.90 (0.83, 0.98) | 0.90 (0.87, 0.94) | 0.67 |
| **Vitamin $B_{12}$ per 2 µg/day** | 0.92 (0.86, 0.99) | 0.86 (0.83, 0.90) | **0.02** |
| **Stone fruit ≥1 per week** | 0.96 (0.93, 1.00) | 0.93 (0.89, 0.98) | 0.76 |
| **Processed meat per 10g/day** | 1.09 (1.03, 1.15) | 0.93 (0.89, 0.97) | 1.00 |
| **Cauliflower ≥1 per week** | 1.03 (1.00, 1.07) | 1.05 (1.01, 1.09) | 1.00 |
| **Riboflavin per 1mg/day** | 0.87 (0.78, 0.98) | 0.93 (0.87, 1.00) | 1.00 |
| **Eggs per 10g/day** | 1.07 (1.03, 1.11) | 0.94 (0.89, 1.00) | 0.29 |
| **Cucumber ≥1 per week** | 0.97 (0.94, 1.00) | 0.94 (0.91, 0.98) | 0.71 |
| **Green peas ≥1 per week** | 1.03 (0.99, 1.07) | 1.07 (1.03, 1.10) | 0.24 |
| **Juice per 100g/day** | 1.03 (0.98, 1.07) | 0.93 (0.90, 0.97) | 1.00 |
| **Baked beans ≥1 per week** | 1.02 (0.99, 1.05) | 1.04 (1.00, 1.08) | 0.71 |
| **Mushrooms ≥1 per week** | 0.96 (0.93, 1.00) | 0.94 (0.90, 0.98) | 0.71 |
| **Calcium per 300mg/day** | 0.93 (0.87, 0.99) | 0.96 (0.92, 1.00) | 0.71 |
| **Brown bread per 10g/day** | 0.90 (0.84, 0.96) | 0.97 (0.94, 1.01) | 1.00 |
| **Cereal per 40g/day** | 0.94 (0.90, 0.99) | 0.95 (0.92, 0.99) | 0.69 |
| **Dried fruit per 10g/day** | 0.95 (0.91, 1.00) | 0.97 (0.93, 1.00) | 0.69 |
| **Folate per 100 µg/day** | 0.92 (0.85, 1.00) | 1.05 (1.01, 1.08) | 0.20 |
| **Sweet Biscuits per 10g/day** | 1.01 (0.95, 1.07) | 1.04 (0.99, 1.08) | 0.75 |

[1] Person-years/ number of incident cases

[2] *p*-heterogeneity between subgroups and was calculated with the inverse variance-weighted average of stratum-specific log hazard ratios.

Abbreviation: SFA saturated fatty acids.

All models were stratified by birth year (≤1921,five-year intervals between 1922 and 1946, >1946), year of completing the baseline dietary questionnaire (per year from 1999 to 2004, and 2005 or later), and 10 regions (9 in England and 1 in Scotland) and adjusted for ethnicity (white/ non-white, missing), Townsend deprivation index (quintiles and missing), education (tertiary, secondary, technical, no qualification, missing), self-rated health (excellent, good, fair, poor, missing), energy intake (500 to <1,000, 1,000 to <1,250, 1,250 to <1,500, 1,500 to <1,750, 1,750 to <2,000, 2,000 to < 2,250, 2,250 to ≤3,500 kcal/day), smoking (never smoker, past smoker, current smoker <15 cigarettes/day, current smoker ≥15 cigarettes/day, missing), alcohol consumption (<1, 1 to <10, 10 to <20, and ≥20g/day), strenuous exercise frequencies (none, <1, 1, 2-3, 4-6, 7 times/week, missing), MHT use (never, past, current, missing), parity (nulliparous, 1-2 children, ≥3 children, missing), age at menopause (<40, 40-44, 45-49, 50-54, ≥55 years, missing), height (quintiles and missing), family history of hip fractures (yes, no, missing), and BMI (<18.5, 18.5 to <20, 20 to <22.5, 22.5 to <25, 25 to <27.5, 27.5 to < 30, 30 to <32.5, ≥32.5 kg/$m^2$, missing). For analyses of total energy and alcohol, covariates of energy intake and alcohol consumption were taken out of the model respectively.

**STable 8. Risks of hip fracture with 60 significant dietary factors in subgroups of frequency of strenuous exercise**

| Dietary factors | Hazard Ratios (95% CI) | | | |
|---|---|---|---|---|
| | **Rare/<1 per wk** (5,782,632/15,772)[1] | **1-6 times per wk** (4,361,583/9,948)[1] | **Daily** (294,246/873)[1] | *p*-het[2] |
| **Chicken per 20g/day** | 0.83 (0.79, 0.88) | 0.87 (0.81, 0.93) | 0.76 (0.60, 0.95) | 0.39 |
| **Broccoli ≥1 per week** | 0.88 (0.85, 0.91) | 0.92 (0.87, 0.96) | 0.80 (0.68, 0.94) | 0.15 |
| **Vegetables per 100g/day** | 0.88 (0.84, 0.91) | 0.90 (0.85, 0.95) | 0.73 (0.61, 0.87) | 0.09 |
| **Protein per 15g/day** | 0.77 (0.72, 0.84) | 0.84 (0.76, 0.92) | 0.68 (0.49, 0.94) | 0.25 |
| **Chips per 25g/day** | 1.13 (1.08, 1.18) | 1.18 (1.11, 1.25) | 1.08 (0.89, 1.32) | 0.43 |
| **Zinc per 2mg/day** | 0.82 (0.76, 0.88) | 0.82 (0.74, 0.90) | 0.70 (0.51, 0.98) | 0.65 |
| **Garlic ≥1 per week** | 0.93 (0.90, 0.97) | 0.88 (0.85, 0.92) | 0.84 (0.72, 0.97) | 0.09 |
| **Fizzy drinks per 50g/day** | 1.17 (1.10, 1.24) | 1.19 (1.10, 1.30) | 1.26 (0.96, 1.65) | 0.84 |
| **Pasta per 20g/day** | 0.89 (0.84, 0.94) | 0.84 (0.78, 0.90) | 0.77 (0.61, 0.98) | 0.28 |
| **Carotene per 1000 µg/day** | 0.92 (0.88, 0.95) | 0.92 (0.87, 0.96) | 0.82 (0.69, 0.96) | 0.41 |
| **Fiber per 5g/day** | 0.90 (0.86, 0.95) | 0.87 (0.82, 0.93) | 0.83 (0.67, 1.02) | 0.58 |
| **Niacin per 10mg/day** | 0.81 (0.74, 0.88) | 0.86 (0.77, 0.96) | 0.62 (0.43, 0.90) | 0.22 |
| **Fruit (excluding juice) per 200g/day** | 0.88 (0.83, 0.93) | 0.89 (0.83, 0.96) | 0.74 (0.58, 0.95) | 0.37 |
| **Phosphorus per 300mg/day** | 0.85 (0.79, 0.92) | 0.86 (0.78, 0.95) | 0.86 (0.62, 1.19) | 0.98 |
| **Onion ≥1 per week** | 0.95 (0.92, 0.99) | 0.89 (0.84, 0.93) | 0.77 (0.65, 0.91) | **0.01** |
| **Peppers ≥1 per week** | 0.94 (0.91, 0.97) | 0.95 (0.91, 0.99) | 0.83 (0.72, 0.97) | 0.23 |
| **Retinol per 250 µg/day** | 1.09 (1.03, 1.16) | 1.16 (1.08, 1.25) | 1.15 (0.90, 1.47) | 0.42 |
| **Alcohol per 10g/day** | 0.97 (0.95, 0.99) | 0.96 (0.93, 0.98) | 0.98 (0.90, 1.07) | 0.79 |
| **Wholegrains per 20g/day** | 0.88 (0.82, 0.95) | 0.84 (0.77, 0.92) | 0.86 (0.64, 1.17) | 0.73 |
| **Selenium per 10mg/day** | 0.87 (0.82, 0.93) | 0.91 (0.85, 0.99) | 0.83 (0.65, 1.08) | 0.59 |
| **Yoghurt per 50g/day** | 0.92 (0.88, 0.96) | 0.94 (0.89, 0.99) | 0.94 (0.79, 1.11) | 0.82 |
| **Vitamin B6 per 1mg/day** | 0.74 (0.64, 0.86) | 0.78 (0.64, 0.94) | 1.01 (0.52, 1.94) | 0.63 |
| **Rice per 10g/day** | 0.93 (0.90, 0.97) | 0.95 (0.91, 1.00) | 0.79 (0.68, 0.92) | 0.07 |
| **Magnesium per100mg/day** | 0.84 (0.77, 0.92) | 0.87 (0.78, 0.98) | 0.79 (0.54, 1.16) | 0.83 |
| **Apple ≥1 per week** | 0.93 (0.90, 0.97) | 0.95 (0.91, 1.00) | 0.85 (0.72, 1.00) | 0.40 |
| **Banana ≥1 per week** | 0.91 (0.88, 0.95) | 0.96 (0.91, 1.02) | 0.90 (0.75, 1.08) | 0.30 |
| **Courgette ≥1 per week** | 0.95 (0.91, 0.98) | 0.93 (0.89, 0.97) | 0.94 (0.80, 1.11) | 0.76 |
| **Carrot ≥1 per week** | 0.93 (0.89, 0.98) | 0.90 (0.84, 0.97) | 0.82 (0.66, 1.01) | 0.44 |
| **Brussel sprouts ≥1 per week** | 1.05 (1.01, 1.08) | 1.04 (1.00, 1.09) | 1.01 (0.88, 1.16) | 0.84 |
| **Nuts per 10g/day** | 0.93 (0.87, 1.00) | 0.88 (0.81, 0.95) | 0.77 (0.58, 1.03) | 0.32 |
| **Squash drinks per 50g/day** | 1.08 (1.04, 1.13) | 1.03 (0.97, 1.08) | 1.14 (0.97, 1.35) | 0.28 |
| **Crisps per 10g/day** | 1.26 (1.13, 1.40) | 1.02 (0.87, 1.19) | 1.60 (1.07, 2.39) | **0.03** |
| **Free sugars per 30g/day** | 1.12 (1.05, 1.19) | 1.04 (0.96, 1.13) | 1.20 (0.91, 1.58) | 0.29 |
| **Oily fish per 10g/day** | 0.91 (0.88, 0.96) | 0.98 (0.93, 1.03) | 0.85 (0.71, 1.02) | 0.05 |
| **Stewed fruit ≥1 per week** | 1.05 (1.02, 1.09) | 1.05 (1.01, 1.09) | 0.93 (0.80, 1.08) | 0.29 |
| **White bread per 10g/day** | 1.07 (1.02, 1.13) | 1.09 (1.02, 1.17) | 1.10 (0.87, 1.38) | 0.90 |
| **Potassium per 1000mg/day** | 0.88 (0.82, 0.96) | 0.94 (0.85, 1.04) | 0.74 (0.53, 1.05) | 0.32 |
| **SFA per 10g/day** | 1.10 (1.03, 1.18) | 1.09 (1.00, 1.19) | 1.27 (0.94, 1.70) | 0.62 |
| **Soup per 40g/day** | 1.06 (1.01, 1.11) | 1.08 (1.02, 1.14) | 0.95 (0.76, 1.17) | 0.51 |
| **Thiamin per 1mg/day** | 0.76 (0.64, 0.92) | 0.86 (0.68, 1.09) | 0.79 (0.36, 1.75) | 0.72 |
| **Avocado ≥1 per week** | 0.92 (0.86, 0.98) | 0.95 (0.90, 1.02) | 1.00 (0.80, 1.25) | 0.67 |

| | | | | |
|---|---|---|---|---|
| **Lettuce ≥1 per week** | 0.93 (0.90, 0.97) | 1.00 (0.95, 1.05) | 0.79 (0.68, 0.93) | **0.01** |
| **Vitamin C per 100mg/day** | 0.91 (0.85, 0.98) | 0.91 (0.82, 1.00) | 0.93 (0.67, 1.28) | 0.99 |
| **Vitamin $B_{12}$ per 2 μg/day** | 0.90 (0.85, 0.96) | 0.95 (0.88, 1.03) | 0.90 (0.69, 1.16) | 0.56 |
| **Stone fruit ≥1 per week** | 0.97 (0.94, 1.00) | 0.95 (0.92, 1.00) | 0.92 (0.79, 1.06) | 0.61 |
| **Processed meat per 10g/day** | 1.08 (1.03, 1.14) | 1.03 (0.97, 1.10) | 0.97 (0.77, 1.22) | 0.39 |
| **Cauliflower ≥1 per week** | 1.04 (1.01, 1.07) | 1.03 (0.99, 1.08) | 0.97 (0.84, 1.12) | 0.63 |
| **Riboflavin per 1mg/day** | 0.89 (0.80, 0.99) | 0.90 (0.78, 1.03) | 0.92 (0.59, 1.45) | 0.98 |
| **Eggs per 10g/day** | 1.04 (1.00, 1.07) | 1.04 (0.99, 1.09) | 1.03 (0.88, 1.20) | 0.99 |
| **Cucumber ≥1 per week** | 0.95 (0.92, 0.98) | 0.99 (0.95, 1.03) | 0.95 (0.82, 1.10) | 0.28 |
| **Green peas ≥1 per week** | 1.04 (1.01, 1.08) | 1.02 (0.98, 1.07) | 1.00 (0.86, 1.17) | 0.73 |
| **Juice per 100g/day** | 1.06 (1.01, 1.10) | 1.02 (0.97, 1.08) | 1.11 (0.93, 1.33) | 0.44 |
| **Baked beans ≥1 per week** | 1.04 (1.00, 1.07) | 1.02 (0.98, 1.07) | 0.98 (0.85, 1.13) | 0.62 |
| **Mushrooms ≥1 per week** | 0.97 (0.94, 1.01) | 0.98 (0.94, 1.02) | 0.80 (0.70, 0.93) | **0.03** |
| **Calcium per 300mg/day** | 0.91 (0.85, 0.97) | 0.98 (0.90, 1.06) | 1.08 (0.83, 1.42) | 0.23 |
| **Brown bread per 10g/day** | 0.93 (0.88, 0.99) | 0.95 (0.88, 1.03) | 0.94 (0.72, 1.23) | 0.91 |
| **Cereal per 40g/day** | 0.99 (0.95, 1.03) | 0.92 (0.87, 0.97) | 0.97 (0.81, 1.17) | 0.11 |
| **Dried fruit per 10g/day** | 0.97 (0.92, 1.01) | 0.96 (0.91, 1.01) | 0.86 (0.71, 1.05) | 0.50 |
| **Folate per 100 μg/day** | 0.93 (0.87, 1.00) | 0.95 (0.86, 1.04) | 0.99 (0.72, 1.35) | 0.89 |
| **Sweet Biscuits per 10g/day** | 1.05 (0.99, 1.11) | 1.05 (0.98, 1.13) | 1.10 (0.87, 1.39) | 0.93 |

[1] Person-years/ number of incident cases

[2] *p*-heterogeneity among all subgroups and was calculated with the inverse variance-weighted average of stratum-specific log hazard ratios.

Abbreviation: SFA saturated fatty acids.

All models were stratified by birth year (≤1921,five-year intervals between 1922 and 1946, >1946), year of completing the baseline dietary questionnaire (per year from 1999 to 2004, and 2005 or later), and 10 regions (9 in England and 1 in Scotland) and adjusted for ethnicity (white/ non-white, missing), Townsend deprivation index (quintiles and missing), education (tertiary, secondary, technical, no qualification, missing), self-rated health (excellent, good, fair, poor, missing), energy intake (500 to <1,000, 1,000 to <1,250, 1,250 to <1,500, 1,500 to <1,750, 1,750 to <2,000, 2,000 to < 2,250, 2,250 to ≤3,500 kcal/day), smoking (never smoker, past smoker, current smoker <15 cigarettes/day, current smoker ≥15 cigarettes/day, missing), alcohol consumption (<1, 1 to <10, 10 to <20, and ≥20g/day), strenuous exercise frequencies (none, <1, 1, 2-3, 4-6, 7 times/week, missing), MHT use (never, past, current, missing), parity (nulliparous, 1-2 children, ≥3 children, missing), age at menopause (<40, 40-44, 45-49, 50-54, ≥55 years, missing), height (quintiles and missing), family history of hip fractures (yes, no, missing), and BMI (<18.5, 18.5 to <20, 20 to <22.5, 22.5 to <25, 25 to <27.5, 27.5 to < 30, 30 to <32.5, ≥32.5 kg/$m^2$, missing). For analyses of total energy and alcohol, covariates of energy intake and alcohol consumption were taken out of the model respectively.

**STable 9. Risks of hip fracture with 60 significant dietary factors in subgroups of menopausal hormone therapy use**

| Dietary factors | Hazard Ratios (95% CI) Never (4,838,384/13,690)[1] | Hazard Ratios (95% CI) Ever (5,616,314/12,917)[1] | *p*-het[2] |
|---|---|---|---|
| **Chicken per 20g/day** | 0.87 (0.82, 0.92) | 0.81 (0.76, 0.86) | 0.10 |
| **Broccoli ≥1 per week** | 0.88 (0.85, 0.92) | 0.90 (0.87, 0.94) | 0.43 |
| **Vegetables per 100g/day** | 0.88 (0.84, 0.92) | 0.88 (0.84, 0.92) | 1.00 |
| **Protein per 15g/day** | 0.82 (0.76, 0.89) | 0.75 (0.69, 0.82) | 0.13 |
| **Chips per 25g/day** | 1.12 (1.07, 1.18) | 1.15 (1.10, 1.21) | 0.45 |
| **Zinc per 2mg/day** | 0.83 (0.76, 0.90) | 0.79 (0.73, 0.86) | 0.41 |
| **Garlic ≥1 per week** | 0.91 (0.87, 0.94) | 0.90 (0.87, 0.94) | 0.69 |
| **Fizzy drinks per 50g/day** | 1.16 (1.08, 1.24) | 1.20 (1.12, 1.29) | 0.50 |
| **Pasta per 20g/day** | 0.87 (0.82, 0.93) | 0.86 (0.81, 0.92) | 0.80 |
| **Carotene per 1000 μg/day** | 0.92 (0.88, 0.95) | 0.90 (0.87, 0.94) | 0.43 |
| **Fiber per 5g/day** | 0.89 (0.85, 0.94) | 0.89 (0.84, 0.93) | 1.00 |
| **Niacin per 10mg/day** | 0.86 (0.78, 0.94) | 0.77 (0.70, 0.85) | 0.11 |
| **Fruit (excluding juice) per 200g/day** | 0.90 (0.85, 0.96) | 0.86 (0.81, 0.92) | 0.31 |
| **Phosphorus per 300mg/day** | 0.85 (0.79, 0.92) | 0.85 (0.78, 0.93) | 1.00 |
| **Onion ≥1 per week** | 0.94 (0.90, 0.98) | 0.90 (0.87, 0.94) | 0.14 |
| **Peppers ≥1 per week** | 0.94 (0.91, 0.98) | 0.93 (0.90, 0.96) | 0.67 |
| **Retinol per 250 μg/day** | 1.09 (1.02, 1.16) | 1.15 (1.08, 1.23) | 0.25 |
| **Alcohol per 10g/day** | 0.95 (0.93, 0.97) | 0.97 (0.95, 1.00) | 0.22 |
| **Wholegrains per 20g/day** | 0.84 (0.78, 0.91) | 0.89 (0.82, 0.96) | 0.30 |
| **Selenium per 10mg/day** | 0.87 (0.82, 0.93) | 0.90 (0.84, 0.96) | 0.47 |
| **Yoghurt per 50g/day** | 0.94 (0.90, 0.99) | 0.91 (0.87, 0.95) | 0.33 |
| **Vitamin B6 per 1mg/day** | 0.89 (0.76, 1.05) | 0.63 (0.53, 0.74) | **0.004** |
| **Rice per 10g/day** | 0.93 (0.90, 0.97) | 0.94 (0.91, 0.98) | 0.69 |
| **Magnesium per100mg/day** | 0.85 (0.78, 0.94) | 0.86 (0.77, 0.94) | 0.87 |
| **Apple ≥1 per week** | 0.95 (0.92, 0.99) | 0.92 (0.88, 0.96) | 0.27 |
| **Banana ≥1 per week** | 0.92 (0.88, 0.96) | 0.93 (0.89, 0.98) | 0.74 |
| **Courgette ≥1 per week** | 0.93 (0.89, 0.97) | 0.95 (0.91, 0.99) | 0.49 |
| **Carrot ≥1 per week** | 0.92 (0.87, 0.97) | 0.93 (0.88, 0.98) | 0.78 |
| **Brussel sprouts ≥1 per week** | 1.05 (1.01, 1.09) | 1.05 (1.01, 1.09) | 1.00 |
| **Nuts per 10g/day** | 0.92 (0.85, 0.99) | 0.88 (0.82, 0.95) | 0.41 |
| **Squash drinks per 50g/day** | 1.05 (1.01, 1.10) | 1.07 (1.02, 1.12) | 0.56 |
| **Crisps per 10g/day** | 1.32 (1.17, 1.48) | 1.05 (0.92, 1.19) | **0.01** |
| **Free sugars per 30g/day** | 1.08 (1.01, 1.16) | 1.13 (1.05, 1.21) | 0.37 |
| **Oily fish per 10g/day** | 0.91 (0.87, 0.96) | 0.96 (0.92, 1.01) | 0.12 |
| **Stewed fruit ≥1 per week** | 1.07 (1.03, 1.11) | 1.02 (0.99, 1.06) | 0.06 |
| **White bread per 10g/day** | 1.05 (0.99, 1.11) | 1.12 (1.05, 1.19) | 0.14 |
| **Potassium per 1000mg/day** | 0.92 (0.84, 1.00) | 0.89 (0.81, 0.97) | 0.60 |
| **SFA per 10g/day** | 1.07 (1.00, 1.15) | 1.11 (1.03, 1.19) | 0.47 |
| **Soup per 40g/day** | 1.09 (1.04, 1.15) | 1.04 (0.99, 1.10) | 0.21 |
| **Thiamin per 1mg/day** | 0.87 (0.72, 1.06) | 0.72 (0.58, 0.88) | 0.19 |
| **Avocado ≥1 per week** | 0.88 (0.82, 0.94) | 0.98 (0.92, 1.04) | **0.02** |
| **Lettuce ≥1 per week** | 0.96 (0.92, 1.00) | 0.94 (0.90, 0.98) | 0.49 |

| | | | |
|---|---|---|---|
| **Vitamin C per 100mg/day** | 0.92 (0.85, 1.00) | 0.89 (0.82, 0.97) | 0.58 |
| **Vitamin $B_{12}$ per 2 µg/day** | 0.94 (0.88, 1.01) | 0.90 (0.84, 0.96) | 0.37 |
| **Stone fruit ≥1 per week** | 0.98 (0.94, 1.01) | 0.95 (0.91, 0.98) | 0.24 |
| **Processed meat per 10g/day** | 1.06 (1.00, 1.12) | 1.06 (1.00, 1.12) | 1.00 |
| **Cauliflower ≥1 per week** | 1.03 (0.99, 1.07) | 1.03 (1.00, 1.07) | 1.00 |
| **Riboflavin per 1mg/day** | 0.94 (0.84, 1.05) | 0.87 (0.77, 0.98) | 0.36 |
| **Eggs per 10g/day** | 1.02 (0.98, 1.06) | 1.06 (1.01, 1.10) | 0.19 |
| **Cucumber ≥1 per week** | 0.97 (0.93, 1.00) | 0.96 (0.93, 1.00) | 0.69 |
| **Green peas ≥1 per week** | 1.05 (1.01, 1.09) | 1.02 (0.98, 1.06) | 0.30 |
| **Juice per 100g/day** | 1.03 (0.98, 1.08) | 1.06 (1.01, 1.11) | 0.41 |
| **Baked beans ≥1 per week** | 1.00 (0.97, 1.03) | 1.06 (1.03, 1.10) | **0.01** |
| **Mushrooms ≥1 per week** | 0.95 (0.92, 0.99) | 0.99 (0.95, 1.03) | 0.14 |
| **Calcium per 300mg/day** | 0.94 (0.88, 1.01) | 0.94 (0.88, 1.01) | 1.00 |
| **Brown bread per 10g/day** | 0.92 (0.86, 0.99) | 0.95 (0.89, 1.02) | 0.52 |
| **Cereal per 40g/day** | 1.00 (0.95, 1.04) | 0.94 (0.90, 0.99) | 0.06 |
| **Dried fruit per 10g/day** | 0.98 (0.94, 1.03) | 0.94 (0.89, 0.98) | 0.22 |
| **Folate per 100 µg/day** | 0.96 (0.89, 1.04) | 0.91 (0.84, 0.99) | 0.35 |
| **Sweet Biscuits per 10g/day** | 1.06 (1.00, 1.12) | 1.06 (0.99, 1.12) | 1.00 |

[1] Person-years/ number of incident cases

[2] *p*-heterogeneity between subgroups and was calculated with the inverse variance-weighted average of stratum-specific log hazard ratios.

Abbreviation: SFA saturated fatty acids.

All models were stratified by birth year (≤1921,five-year intervals between 1922 and 1946, >1946), year of completing the baseline dietary questionnaire (per year from 1999 to 2004, and 2005 or later), and 10 regions (9 in England and 1 in Scotland) and adjusted for ethnicity (white/ non-white, missing), Townsend deprivation index (quintiles and missing), education (tertiary, secondary, technical, no qualification, missing), self-rated health (excellent, good, fair, poor, missing), energy intake (500 to <1,000, 1,000 to <1,250, 1,250 to <1,500, 1,500 to <1,750, 1,750 to <2,000, 2,000 to < 2,250, 2,250 to ≤3,500 kcal/day), smoking (never smoker, past smoker, current smoker <15 cigarettes/day, current smoker ≥15 cigarettes/day, missing), alcohol consumption (<1, 1 to <10, 10 to <20, and ≥20g/day), strenuous exercise frequencies (none, <1, 1, 2-3, 4-6, 7 times/week, missing), MHT use (never, past, current, missing), parity (nulliparous, 1-2 children, ≥3 children, missing), age at menopause (<40, 40-44, 45-49, 50-54, ≥55 years, missing), height (quintiles and missing), family history of hip fractures (yes, no, missing), and BMI (<18.5, 18.5 to <20, 20 to <22.5, 22.5 to <25, 25 to <27.5, 27.5 to < 30, 30 to <32.5, ≥32.5 kg/$m^2$, missing). For analyses of total energy and alcohol, covariates of energy intake and alcohol consumption were taken out of the model respectively.

**Stable10. Risks of hip fracture with 60 significant dietary factors in subgroups of deprivation status**

| Dietary factors | Hazard Ratios (95% CI) Least deprived (2,511,975/6,081)[1] | Most deprived (1,491,101/4,216)[1] | *p*-het[2] |
|---|---|---|---|
| **Chicken per 20g/day** | 0.86 (0.79, 0.94) | 0.83 (0.75, 0.92) | 0.60 |
| **Broccoli ≥1 per week** | 0.88 (0.83, 0.94) | 0.86 (0.81, 0.92) | 0.61 |
| **Vegetables per 100g/day** | 0.86 (0.81, 0.92) | 0.80 (0.74, 0.87) | 0.17 |
| **Protein per 15g/day** | 0.77 (0.68, 0.87) | 0.86 (0.74, 0.99) | 0.26 |
| **Chips per 25g/day** | 1.17 (1.08, 1.26) | 1.18 (1.09, 1.29) | 0.88 |
| **Zinc per 2mg/day** | 0.79 (0.70, 0.89) | 0.89 (0.77, 1.02) | 0.21 |
| **Garlic ≥1 per week** | 0.92 (0.87, 0.97) | 0.93 (0.87, 1.00) | 0.81 |
| **Fizzy drinks per 50g/day** | 1.35 (1.21, 1.50) | 1.15 (1.03, 1.29) | **0.04** |
| **Pasta per 20g/day** | 0.83 (0.76, 0.91) | 0.85 (0.77, 0.95) | 0.74 |
| **Carotene per 1000 µg/day** | 0.89 (0.84, 0.95) | 0.87 (0.81, 0.94) | 0.64 |
| **Fiber per 5g/day** | 0.84 (0.78, 0.91) | 0.89 (0.81, 0.98) | 0.36 |
| **Niacin per 10mg/day** | 0.78 (0.68, 0.90) | 0.85 (0.72, 1.00) | 0.44 |
| **Fruit (excluding juice) per 200g/day** | 0.93 (0.84, 1.02) | 0.86 (0.77, 0.97) | 0.31 |
| **Phosphorus per 300mg/day** | 0.81 (0.72, 0.91) | 0.92 (0.80, 1.06) | 0.17 |
| **Onion ≥1 per week** | 0.96 (0.90, 1.02) | 0.91 (0.85, 0.97) | 0.25 |
| **Peppers ≥1 per week** | 0.93 (0.88, 0.98) | 0.92 (0.86, 0.98) | 0.80 |
| **Retinol per 250 µg/day** | 1.15 (1.05, 1.27) | 1.06 (0.94, 1.19) | 0.29 |
| **Alcohol per 10g/day** | 0.96 (0.93, 1.00) | 0.98 (0.94, 1.02) | 0.46 |
| **Wholegrains per 20g/day** | 0.82 (0.73, 0.92) | 0.86 (0.75, 0.98) | 0.60 |
| **Selenium per 10mg/day** | 0.96 (0.87, 1.06) | 0.79 (0.71, 0.89) | **0.01** |
| **Yoghurt per 50g/day** | 0.91 (0.85, 0.97) | 0.91 (0.84, 0.99) | 1.00 |
| **Vitamin B6 per 1mg/day** | 0.77 (0.60, 0.98) | 0.79 (0.59, 1.05) | 0.89 |
| **Rice per 10g/day** | 0.95 (0.90, 1.01) | 0.96 (0.90, 1.03) | 0.82 |
| **Magnesium per100mg/day** | 0.85 (0.74, 0.98) | 0.84 (0.71, 1.00) | 0.92 |
| **Apple ≥1 per week** | 0.91 (0.86, 0.97) | 0.99 (0.93, 1.06) | 0.06 |
| **Banana ≥1 per week** | 0.99 (0.92, 1.06) | 0.88 (0.81, 0.94) | **0.02** |
| **Courgette ≥1 per week** | 0.94 (0.88, 0.99) | 0.88 (0.80, 0.96) | 0.23 |
| **Carrot ≥1 per week** | 0.93 (0.85, 1.02) | 1.00 (0.92, 1.10) | 0.27 |
| **Brussel sprouts ≥1 per week** | 1.03 (0.98, 1.09) | 1.06 (1.00, 1.13) | 0.49 |
| **Nuts per 10g/day** | 0.88 (0.79, 0.98) | 0.93 (0.81, 1.07) | 0.54 |
| **Squash drinks per 50g/day** | 0.99 (0.93, 1.07) | 1.03 (0.95, 1.11) | 0.46 |
| **Crisps per 10g/day** | 1.13 (0.94, 1.37) | 1.14 (0.92, 1.41) | 0.95 |
| **Free sugars per 30g/day** | 1.12 (1.00, 1.24) | 1.03 (0.91, 1.16) | 0.31 |
| **Oily fish per 10g/day** | 0.95 (0.89, 1.01) | 0.97 (0.89, 1.05) | 0.69 |
| **Stewed fruit ≥1 per week** | 1.02 (0.96, 1.07) | 1.07 (1.00, 1.14) | 0.27 |
| **White bread per 10g/day** | 1.13 (1.03, 1.24) | 1.03 (0.93, 1.14) | 0.19 |
| **Potassium per 1000mg/day** | 0.96 (0.84, 1.09) | 0.92 (0.79, 1.07) | 0.68 |
| **SFA per 10g/day** | 1.12 (1.01, 1.25) | 1.16 (1.02, 1.32) | 0.68 |
| **Soup per 40g/day** | 1.06 (0.99, 1.15) | 1.12 (1.03, 1.23) | 0.35 |
| **Thiamin per 1mg/day** | 0.82 (0.61, 1.10) | 0.81 (0.57, 1.16) | 0.96 |
| **Avocado ≥1 per week** | 0.91 (0.83, 0.99) | 0.95 (0.83, 1.10) | 0.61 |
| **Lettuce ≥1 per week** | 0.99 (0.93, 1.06) | 0.96 (0.90, 1.03) | 0.52 |

| | | | |
|---|---|---|---|
| **Vitamin C per 100mg/day** | 0.92 (0.81, 1.04) | 0.90 (0.78, 1.05) | 0.82 |
| **Vitamin $B_{12}$ per 2 µg/day** | 0.91 (0.82, 1.01) | 0.98 (0.87, 1.11) | 0.36 |
| **Stone fruit ≥1 per week** | 0.98 (0.93, 1.03) | 1.00 (0.94, 1.07) | 0.63 |
| **Processed meat per 10g/day** | 1.09 (1.00, 1.18) | 1.04 (0.94, 1.16) | 0.49 |
| **Cauliflower ≥1 per week** | 1.04 (0.99, 1.10) | 1.03 (0.96, 1.10) | 0.83 |
| **Riboflavin per 1mg/day** | 0.85 (0.71, 1.01) | 0.95 (0.77, 1.16) | 0.42 |
| **Eggs per 10g/day** | 1.07 (1.01, 1.14) | 1.02 (0.95, 1.10) | 0.32 |
| **Cucumber ≥1 per week** | 1.01 (0.96, 1.07) | 0.94 (0.88, 1.00) | 0.09 |
| **Green peas ≥1 per week** | 1.06 (1.00, 1.12) | 1.07 (1.00, 1.14) | 0.83 |
| **Juice per 100g/day** | 1.03 (0.96, 1.10) | 1.06 (0.97, 1.15) | 0.61 |
| **Baked beans ≥1 per week** | 1.01 (0.96, 1.07) | 1.08 (1.01, 1.15) | 0.12 |
| **Mushrooms ≥1 per week** | 1.02 (0.96, 1.07) | 0.94 (0.88, 1.00) | 0.06 |
| **Calcium per 300mg/day** | 0.96 (0.87, 1.07) | 0.97 (0.86, 1.10) | 0.90 |
| **Brown bread per 10g/day** | 0.91 (0.82, 1.01) | 0.89 (0.78, 1.00) | 0.79 |
| **Cereal per 40g/day** | 0.89 (0.83, 0.95) | 1.01 (0.93, 1.10) | **0.02** |
| **Dried fruit per 10g/day** | 0.97 (0.91, 1.04) | 0.95 (0.86, 1.04) | 0.73 |
| **Folate per 100 µg/day** | 0.87 (0.77, 0.98) | 1.00 (0.87, 1.16) | 0.15 |
| **Sweet Biscuits per 10g/day** | 1.04 (0.95, 1.14) | 1.04 (0.94, 1.15) | 1.00 |

[1] Person-years/ number of incident cases

[2] *p*-heterogeneity between subgroups and was calculated with the inverse variance-weighted average of stratum-specific log hazard ratios.

Abbreviation: SFA saturated fatty acids.

All models were stratified by birth year (≤1921,five-year intervals between 1922 and 1946, >1946), year of completing the baseline dietary questionnaire (per year from 1999 to 2004, and 2005 or later), and 10 regions (9 in England and 1 in Scotland) and adjusted for ethnicity (white/ non-white, missing), Townsend deprivation index (quintiles and missing), education (tertiary, secondary, technical, no qualification, missing), self-rated health (excellent, good, fair, poor, missing), energy intake (500 to <1,000, 1,000 to <1,250, 1,250 to <1,500, 1,500 to <1,750, 1,750 to <2,000, 2,000 to < 2,250, 2,250 to ≤3,500 kcal/day), smoking (never smoker, past smoker, current smoker <15 cigarettes/day, current smoker ≥15 cigarettes/day, missing), alcohol consumption (<1, 1 to <10, 10 to <20, and ≥20g/day), strenuous exercise frequencies (none, <1, 1, 2-3, 4-6, 7 times/week, missing), MHT use (never, past, current, missing), parity (nulliparous, 1-2 children, ≥3 children, missing), age at menopause (<40, 40-44, 45-49, 50-54, ≥55 years, missing), height (quintiles and missing), family history of hip fractures (yes, no, missing), and BMI (<18.5, 18.5 to <20, 20 to <22.5, 22.5 to <25, 25 to <27.5, 27.5 to < 30, 30 to <32.5, ≥32.5 kg/$m^2$, missing). For analyses of total energy and alcohol, covariates of energy intake and alcohol consumption were taken out of the model respectively.

**STable 11. Risk of hip fracture for top ten foods and nutrients in the "healthiest" subgroup** (Person-years = 633,195, 1,771 incident cases)

| Foods | HR (95% CI) | *p*-value | Nutrients | HR (95% CI) | *p*-value |
|---|---|---|---|---|---|
| **Chicken per 20g** | 0.86 (0.73, 1.02) | 0.09 | **Protein per 15g** | 0.83 (0.66, 1.06) | 0.14 |
| **Vegetables per 100g** | 0.85 (0.75, 0.96) | **0.01** | **Zinc per 2mg** | 0.93 (0.74, 1.18) | 0.56 |
| **Chips per 25g** | 1.05 (0.91, 1.22) | 0.49 | **Carotene per 1000µg** | 0.93 (0.83, 1.05) | 0.24 |
| **Fizzy drinks per 50g** | 1.10 (0.86, 1.40) | 0.43 | **Fiber per 5g** | 0.81 (0.69, 0.94) | **0.01** |
| **Pasta intake per 20g** | 0.83 (0.69, 0.98) | **0.03** | **Niacin intake per 10mg** | 0.88 (0.67, 1.16) | 0.36 |

Note: The "healthiest" subgroup was defined as people who had BMI in 20-24.9 kg/m$^2$, never smoker, mild/moderate drinker with alcohol consumption 1 to <10g/day, and MHT never user. The sequence of the dietary factors was based on *p*-values in the main analyses.

The results were based on the fully adjusted models, which were stratified by birth year (≤1921,five-year intervals between 1922 and 1946, >1946), year of completing the baseline dietary questionnaire (per year from 1999 to 2004, and 2005 or later), and 10 regions (9 in England and 1 in Scotland) and adjusted for ethnicity (white/ non-white, missing), Townsend deprivation index (quintiles and missing), education (tertiary, secondary, technical, no qualification, missing), self-rated health (excellent, good, fair, poor, missing), energy intake (500 to <1,000, 1,000 to <1,250, 1,250 to <1,500, 1,500 to <1,750, 1,750 to <2,000, 2,000 to < 2,250, 2,250 to ≤3,500 kcal/day), smoking (never smoker, past smoker, current smoker <15 cigarettes/day, current smoker ≥15 cigarettes/day, missing), alcohol consumption (<1, 1 to <10, 10 to <20, and ≥20g/day), strenuous exercise frequencies (none, <1, 1, 2-3, 4-6, 7 times/week, missing), MHT use (never, past, current, missing), parity (nulliparous, 1-2 children, ≥3 children, missing), age at menopause (<40, 40-44, 45-49, 50-54, ≥55 years, missing), height (quintiles and missing), family history of hip fractures (yes, no, missing), and BMI (<18.5, 18.5 to <20, 20 to <22.5, 22.5 to <25, 25 to <27.5, 27.5 to < 30, 30 to <32.5, ≥32.5 kg/m$^2$, missing).

**STable 12. Risks of hip fracture with 99 dietary factors after excluding first five years of follow-up** (Person-years = 7,969,339, 25,940 incident cases)

| Dietary factors | Model 1 | Model 2 | Model 3 | |
|---|---|---|---|---|
| | HR (95% CI) | HR (95% CI) | HR (95% CI) | pvalue[1] |
| **Chicken per 20g/day** | 0.77 (0.74, 0.80) | 0.81 (0.78, 0.85) | 0.84 (0.80, 0.87) | 4.37E-16 |
| **Protein per 15g/day** | 0.85 (0.82, 0.88) | 0.74 (0.69, 0.78) | 0.79 (0.74, 0.83) | 2.54E-15 |
| **Broccoli ≥1 per week** | 0.85 (0.82, 0.87) | 0.90 (0.87, 0.93) | 0.90 (0.87, 0.92) | 3.74E-14 |
| **Vegetables per 100g/day** | 0.84 (0.81, 0.86) | 0.87 (0.85, 0.90) | 0.89 (0.86, 0.91) | 1.01E-13 |
| **Zinc per 2mg/day** | 0.84 (0.81, 0.88) | 0.77 (0.73, 0.82) | 0.81 (0.76, 0.85) | 2.91E-13 |
| **Chips per 25g/day** | 1.19 (1.15, 1.23) | 1.13 (1.09, 1.17) | 1.14 (1.10, 1.18) | 1.06E-12 |
| **Garlic ≥1 per week** | 0.87 (0.85, 0.90) | 0.91 (0.88, 0.93) | 0.91 (0.88, 0.93) | 5.17E-12 |
| **Fizzy drinks per 50g/day** | 1.14 (1.08, 1.19) | 1.10 (1.05, 1.16) | 1.19 (1.13, 1.25) | 2.33E-11 |
| **Pasta per 20g/day** | 0.81 (0.77, 0.84) | 0.85 (0.82, 0.89) | 0.87 (0.83, 0.91) | 2.92E-10 |
| **Carotene per 1000 µg/day** | 0.88 (0.86, 0.90) | 0.91 (0.89, 0.94) | 0.91 (0.88, 0.94) | 6.66E-10 |
| **Fiber per 5g/day** | 0.85 (0.83, 0.88) | 0.90 (0.87, 0.93) | 0.89 (0.85, 0.92) | 1.10E-09 |
| **Niacin per 10mg/day** | 0.77 (0.73, 0.82) | 0.76 (0.71, 0.81) | 0.82 (0.76, 0.87) | 3.80E-09 |
| **Fruit (excluding juice) per 200g/day** | 0.78 (0.75, 0.81) | 0.89 (0.85, 0.93) | 0.88 (0.84, 0.92) | 5.70E-08 |
| **Phosphorus per 300mg/day** | 0.89 (0.86, 0.93) | 0.83 (0.78, 0.88) | 0.85 (0.80, 0.90) | 6.47E-08 |
| **Peppers ≥1 per week** | 0.88 (0.86, 0.91) | 0.93 (0.91, 0.96) | 0.94 (0.91, 0.96) | 4.01E-07 |
| **Vitamin B6 per 1mg/day** | 0.68 (0.62, 0.74) | 0.69 (0.61, 0.77) | 0.75 (0.66, 0.84) | 1.09E-06 |
| **Onion ≥1 per week** | 0.87 (0.85, 0.90) | 0.91 (0.89, 0.94) | 0.93 (0.90, 0.96) | 2.04E-06 |
| **Retinol per 250 µg/day** | 1.15 (1.11, 1.19) | 1.13 (1.08, 1.18) | 1.12 (1.07, 1.17) | 3.16E-06 |
| **Magnesium per100mg/day** | 0.86 (0.82, 0.91) | 0.84 (0.78, 0.90) | 0.85 (0.79, 0.91) | 5.29E-06 |
| **Wholegrains per 20g/day** | 0.80 (0.76, 0.85) | 0.91 (0.86, 0.96) | 0.88 (0.83, 0.93) | 6.34E-06 |
| **Rice per 10g/day** | 0.90 (0.87, 0.92) | 0.93 (0.91, 0.96) | 0.94 (0.91, 0.96) | 6.54E-06 |
| **Apple ≥1 per week** | 0.85 (0.83, 0.88) | 0.93 (0.90, 0.96) | 0.94 (0.91, 0.96) | 6.55E-06 |
| **Selenium per 10mg/day** | 0.90 (0.86, 0.93) | 0.87 (0.83, 0.91) | 0.90 (0.86, 0.94) | 7.32E-06 |
| **Yoghurt per 50g/day** | 0.87 (0.84, 0.89) | 0.91 (0.88, 0.94) | 0.93 (0.90, 0.96) | 1.02E-05 |
| **Alcohol per 10g/day** | 0.98 (0.96, 0.99) | 0.98 (0.96, 0.99) | 0.97 (0.95, 0.98) | 1.28E-05 |
| **Courgette ≥1 per week** | 0.90 (0.88, 0.93) | 0.95 (0.92, 0.97) | 0.94 (0.91, 0.96) | 1.42E-05 |
| **Banana ≥1 per week** | 0.87 (0.85, 0.90) | 0.92 (0.89, 0.95) | 0.93 (0.90, 0.96) | 1.75E-05 |
| **Nuts per 10g/day** | 0.89 (0.84, 0.93) | 0.93 (0.88, 0.98) | 0.89 (0.85, 0.94) | 2.78E-05 |
| **Crisps per 10 g/day** | 1.22 (1.12, 1.33) | 1.16 (1.06, 1.26) | 1.19 (1.09, 1.29) | 1.23E-04 |
| **Free sugars per 30g/day** | 1.10 (1.05, 1.14) | 1.11 (1.06, 1.17) | 1.10 (1.05, 1.16) | 1.70E-04 |
| **Brussel sprouts ≥1 per week** | 1.05 (1.02, 1.07) | 1.04 (1.01, 1.07) | 1.05 (1.02, 1.08) | 2.36E-04 |
| **Stewed fruit ≥1 per week** | 1.02 (1.00, 1.05) | 1.05 (1.02, 1.08) | 1.05 (1.02, 1.08) | 2.63E-04 |
| **White bread per 10g/day** | 1.13 (1.08, 1.17) | 1.05 (1.01, 1.09) | 1.08 (1.04, 1.13) | 2.63E-04 |
| **Squash drinks per 50g/day** | 1.04 (1.01, 1.08) | 1.05 (1.01, 1.08) | 1.06 (1.03, 1.09) | 4.67E-04 |
| **Carrot ≥1 per week** | 0.86 (0.82, 0.89) | 0.93 (0.89, 0.96) | 0.93 (0.89, 0.97) | 5.40E-04 |
| **Lettuce ≥1 per week** | 0.90 (0.88, 0.93) | 0.94 (0.92, 0.97) | 0.95 (0.92, 0.98) | 7.63E-04 |
| **Potassium per1000mg/day** | 0.89 (0.85, 0.93) | 0.87 (0.81, 0.92) | 0.90 (0.85, 0.96) | 9.47E-04 |
| **Oily fish per 10g/day** | 0.90 (0.87, 0.93) | 0.95 (0.92, 0.98) | 0.95 (0.92, 0.98) | 9.81E-04 |
| **Avocado ≥1 per week** | 0.93 (0.89, 0.97) | 0.95 (0.91, 0.99) | 0.93 (0.89, 0.97) | 1.18E-03 |
| **Soup per 40g/day** | 1.02 (0.98, 1.05) | 1.04 (1.00, 1.08) | 1.06 (1.02, 1.10) | 1.44E-03 |
| **Processed meat per 10g/day** | 1.03 (0.99, 1.07) | 1.02 (0.98, 1.06) | 1.07 (1.02, 1.11) | 1.69E-03 |
| **SFA per 10g/day** | 1.12 (1.08, 1.16) | 1.12 (1.06, 1.18) | 1.09 (1.03, 1.15) | 1.75E-03 |

| | | | | |
|---|---|---|---|---|
| **Thiamin per 1mg/day** | 0.74 (0.67, 0.82) | 0.73 (0.64, 0.85) | 0.80 (0.69, 0.92) | 2.26E-03 |
| **Mushrooms ≥1 per week** | 0.91 (0.89, 0.93) | 0.94 (0.92, 0.97) | 0.96 (0.94, 0.99) | 2.86E-03 |
| **Vitamin C per 100mg/day** | 0.77 (0.73, 0.82) | 0.90 (0.85, 0.95) | 0.91 (0.86, 0.97) | 3.03E-03 |
| **Stone fruit ≥1 per week** | 0.91 (0.89, 0.93) | 0.96 (0.93, 0.98) | 0.96 (0.94, 0.99) | 3.09E-03 |
| **Vitamin B12 per 2 µg/day** | 0.89 (0.86, 0.93) | 0.89 (0.85, 0.94) | 0.93 (0.88, 0.98) | 3.29E-03 |
| **Cauliflower ≥1 per week** | 1.01 (0.98, 1.03) | 1.01 (0.99, 1.04) | 1.04 (1.01, 1.06) | 6.27E-03 |
| **Juice per 100g/day** | 0.99 (0.96, 1.03) | 1.04 (1.01, 1.08) | 1.05 (1.01, 1.08) | 6.47E-03 |
| **Baked beans ≥1 per week** | 1.02 (0.99, 1.04) | 1.02 (1.00, 1.05) | 1.03 (1.01, 1.06) | 7.43E-03 |
| **Green peas ≥1 per week** | 1.03 (1.00, 1.05) | 1.03 (1.00, 1.06) | 1.04 (1.01, 1.07) | 9.72E-03 |
| **Cucumber ≥1 per week** | 0.91 (0.89, 0.94) | 0.95 (0.93, 0.98) | 0.97 (0.94, 0.99) | 1.02E-02 |
| **Eggs per 10g/day** | 1.02 (1.00, 1.05) | 1.01 (0.98, 1.04) | 1.04 (1.01, 1.07) | 1.25E-02 |
| **Dried fruit per 10g/day** | 0.92 (0.89, 0.95) | 0.99 (0.95, 1.02) | 0.96 (0.92, 0.99) | 1.53E-02 |
| **Riboflavin per 1mg/day** | 0.86 (0.80, 0.92) | 0.86 (0.80, 0.94) | 0.90 (0.83, 0.98) | 1.69E-02 |
| **Sweet Biscuits per 10g/day** | 1.08 (1.04, 1.13) | 1.06 (1.02, 1.11) | 1.05 (1.01, 1.10) | 1.94E-02 |
| **Cereal per 40g/day** | 0.95 (0.92, 0.98) | 0.99 (0.95, 1.02) | 0.96 (0.93, 1.00) | 2.86E-02 |
| **Tea per 500g/day** | 1.05 (1.02, 1.08) | 1.04 (1.01, 1.07) | 1.03 (1.00, 1.06) | 2.89E-02 |
| FDR-CORRECTED NON-SIGNIFICANT BELOW | | | | |
| Calcium per 300mg/day | 0.95 (0.92, 0.99) | 0.94 (0.90, 0.99) | 0.95 (0.90, 1.00) | 3.19E-02 |
| Folate per 100 µg/day | 0.89 (0.85, 0.93) | 0.93 (0.88, 0.98) | 0.94 (0.89, 1.00) | 3.77E-02 |
| Sauce per 10g/day | 0.93 (0.87, 0.98) | 0.92 (0.87, 0.98) | 0.94 (0.89, 1.00) | 5.06E-02 |
| Brown bread per 10g/day | 0.92 (0.87, 0.96) | 0.98 (0.93, 1.03) | 0.95 (0.91, 1.00) | 5.56E-02 |
| Leeks ≥1 per week | 0.94 (0.92, 0.97) | 0.97 (0.94, 0.99) | 0.97 (0.95, 1.00) | 5.84E-02 |
| Parsnip ≥1 per week | 0.95 (0.93, 0.98) | 0.97 (0.94, 1.00) | 0.98 (0.95, 1.00) | 7.67E-02 |
| Red meat per 20g/day | 0.94 (0.92, 0.97) | 0.94 (0.92, 0.97) | 0.98 (0.95, 1.00) | 8.58E-02 |
| Soya milk ≥1 per week | 0.94 (0.88, 1.02) | 0.97 (0.91, 1.05) | 0.94 (0.87, 1.01) | 8.73E-02 |
| Jam per 5g/day | 1.10 (0.99, 1.21) | 1.11 (1.00, 1.23) | 1.09 (0.99, 1.21) | 8.76E-02 |
| Vitamin D per 1 µg/day | 0.92 (0.89, 0.95) | 0.96 (0.92, 0.99) | 0.97 (0.94, 1.01) | 9.49E-02 |
| Tomato ≥1 per week | 0.90 (0.87, 0.94) | 0.96 (0.92, 1.00) | 0.97 (0.93, 1.01) | 1.08E-01 |
| Spinach ≥1 per week | 0.97 (0.94, 1.01) | 0.99 (0.95, 1.03) | 0.97 (0.93, 1.01) | 1.09E-01 |
| Celery ≥1 per week | 0.94 (0.92, 0.97) | 0.97 (0.95, 1.00) | 0.98 (0.95, 1.01) | 1.41E-01 |
| Potatoes per 40g/day | 1.02 (0.98, 1.06) | 1.04 (1.00, 1.08) | 1.03 (0.99, 1.07) | 1.46E-01 |
| Pear ≥1 per week | 0.94 (0.91, 0.96) | 0.97 (0.95, 0.99) | 0.98 (0.96, 1.01) | 1.82E-01 |
| Milk per 200g/day | 1.00 (0.96, 1.05) | 0.97 (0.92, 1.01) | 0.97 (0.93, 1.02) | 1.91E-01 |
| Ice cream ≥1 per week | 1.00 (0.97, 1.03) | 1.01 (0.98, 1.04) | 1.02 (0.99, 1.05) | 2.13E-01 |
| Cheese per 10g/day | 1.03 (1.00, 1.06) | 1.04 (1.01, 1.07) | 1.02 (0.99, 1.05) | 2.68E-01 |
| Red & processed meat per 30g/day | 0.94 (0.92, 0.97) | 0.94 (0.91, 0.97) | 0.98 (0.95, 1.01) | 2.74E-01 |
| Coffee per 500g/day | 1.06 (1.02, 1.10) | 0.99 (0.95, 1.03) | 1.02 (0.98, 1.06) | 2.77E-01 |
| Grapefruit ≥1 per week | 0.97 (0.94, 1.00) | 1.01 (0.98, 1.04) | 1.02 (0.99, 1.05) | 3.17E-01 |
| Soya/ tofu ≥1 per week | 1.05 (0.98, 1.12) | 1.07 (1.00, 1.15) | 1.04 (0.97, 1.11) | 3.21E-01 |
| MUFA per 5g/day | 1.00 (0.97, 1.02) | 0.99 (0.95, 1.03) | 0.98 (0.94, 1.02) | 3.26E-01 |
| Energy per 1000kj/day | 1.00 (0.97, 1.02) | 0.99 (0.97, 1.01) | 0.99 (0.97, 1.01) | 3.27E-01 |
| Cake per 10g/day | 1.01 (0.97, 1.05) | 1.00 (0.96, 1.05) | 0.98 (0.93, 1.02) | 3.52E-01 |
| Boiled sweets per 5g/day | 1.08 (1.00, 1.17) | 1.02 (0.95, 1.10) | 1.04 (0.96, 1.12) | 3.53E-01 |
| Beetroot ≥1 per week | 0.98 (0.96, 1.01) | 1.00 (0.98, 1.03) | 1.01 (0.99, 1.04) | 3.74E-01 |
| Non-oily fish per 10g/day | 0.93 (0.89, 0.98) | 0.95 (0.91, 1.00) | 0.98 (0.93, 1.03) | 3.92E-01 |
| Chick peas/ lentils ≥1 per week | 0.97 (0.93, 1.01) | 1.00 (0.96, 1.04) | 0.98 (0.94, 1.03) | 4.41E-01 |
| Carbohydrate per 50g/day | 0.99 (0.97, 1.02) | 1.03 (0.98, 1.08) | 1.02 (0.97, 1.07) | 4.52E-01 |

| | | | | |
|---|---|---|---|---|
| PUFA per 5g/day | 0.94 (0.88, 1.01) | 0.93 (0.84, 1.01) | 0.97 (0.88, 1.06) | 4.96E-01 |
| Total fat per 20g/day | 1.04 (1.00, 1.07) | 1.04 (0.97, 1.11) | 1.02 (0.96, 1.09) | 5.22E-01 |
| Orange ≥1 per week | 0.94 (0.92, 0.97) | 1.00 (0.97, 1.02) | 1.01 (0.98, 1.03) | 5.93E-01 |
| Cabbage ≥1 per week | 0.98 (0.96, 1.01) | 0.98 (0.96, 1.00) | 0.99 (0.97, 1.02) | 6.21E-01 |
| Total sugars per 50g/day | 0.99 (0.95, 1.03) | 1.03 (0.97, 1.09) | 1.01 (0.96, 1.07) | 6.29E-01 |
| Chocolate per 10g/day | 1.02 (0.98, 1.07) | 1.01 (0.96, 1.06) | 0.99 (0.94, 1.04) | 6.83E-01 |
| Swede ≥1 per week | 0.98 (0.95, 1.01) | 0.99 (0.96, 1.01) | 1.00 (0.98, 1.03) | 8.47E-01 |
| Sweetcorn ≥1 per week | 0.97 (0.94, 1.00) | 0.99 (0.96, 1.02) | 1.00 (0.97, 1.03) | 8.63E-01 |
| Crispbread per 10g/day | 0.97 (0.94, 1.01) | 1.00 (0.96, 1.03) | 1.00 (0.97, 1.04) | 8.63E-01 |
| Vitamin E per 2mg/day | 0.96 (0.92, 1.01) | 0.96 (0.91, 1.02) | 1.00 (0.95, 1.05) | 9.49E-01 |
| Green beans ≥1 per week | 0.97 (0.95, 1.00) | 0.99 (0.97, 1.02) | 1.00 (0.97, 1.03) | 9.76E-01 |

[1] pvalue of model 3 and was the basis of ranking from the smallest to the largest.

Abbreviation: SFA saturated fatty acids, MUFA mono-unsaturated fatty acids, PUFA poly-unsaturated fatty acids.

All models were stratified by birth year (≤1921, five-year intervals between 1922 and 1946, >1946), year of completing the baseline dietary questionnaire (per year from 1999 to 2004, and 2005 or later), and 10 regions (9 in England and 1 in Scotland). Model 1 was adjusted for ethnicity (white/ non-white, missing), Townsend deprivation index (quintiles and missing), education (tertiary, secondary, technical, no qualification, missing). Model 2 was additionally adjusted for self-rated health (excellent, good, fair, poor, missing), energy intake (500 to <1,000, 1,000 to <1,250, 1,250 to <1,500, 1,500 to <1,750, 1,750 to <2,000, 2,000 to < 2,250, 2,250 to ≤3,500 kcal/day), smoking (never smoker, past smoker, current smoker <15 cigarettes/day, current smoker ≥15 cigarettes/day, missing), alcohol consumption (<1, 1 to <10, 10 to <20, and ≥20g/day), strenuous exercise frequencies (none, <1, 1, 2-3, 4-6, 7 times/week, missing), MHT use (never, past, current, missing), parity (nulliparous, 1-2 children, ≥3 children, missing), age at menopause (<40, 40-44, 45-49, 50-54, ≥55 years, missing), height (quintiles and missing), family history of hip fractures (yes, no, missing). Model 3 was additionally adjusted for BMI (<18.5, 18.5 to <20, 20 to <22.5, 22.5 to <25, 25 to <27.5, 27.5 to < 30, 30 to <32.5, ≥32.5 kg/m$^2$, missing). For analyses of total energy and alcohol, covariates of energy intake and alcohol consumption were taken out of the model respectively.
